# Why do we get sick? Genetic evidence for evolutionary trade-offs between fertility, longevity, and disease

**DOI:** 10.1101/2025.06.18.25329754

**Authors:** Eva Brigos-Barril, Claudia Vasallo, Xavier Farré, Carlos Morcillo-Suárez, Sara Polo-Alonso, Blanca Rodríguez-Fernández, Alejandro Valenzuela, Elena Bosch, Hafid Laayouni, Natàlia Vilor-Tejedor, Arcadi Navarro, Gerard Muntané

**Affiliations:** Department of Medicine and Life Sciences, Universitat Pompeu Fabra, Barcelona, Spain; IBE, Institut de Biologia Evolutiva (CSIC-Universitat Pompeu Fabra); Centre for Genomic Regulation (CRG), The Barcelona Institute of Science and Technology, Barcelona, Spain; Genomes for Life-GCAT Lab, CORE Program. Germans Trias i Pujol Research Institute (IGTP), Badalona, Spain; Barcelonaβeta Brain Research Center (BBRC) - Pasqual Maragall Foundation. Barcelona, Spain; Universitat Autònoma de Barcelona, 08193, Cerdanyola del Vallès, Barcelona, Spain; Department of Human Genetics, Radboud University Medical Center, Nijmegen, Netherlands; Hospital del Mar Medical Research Institute, Barcelona, Spain; Institució Catalana de Recerca i Estudis Avançats (ICREA), Barcelona, Spain; Hospital Universitari Institut Pere Mata (HUIPM), Institut d’Investigació Sanitària Pere Virgili (IISPV-CERCA), Universitat Rovira i Virgili (URV), Reus, Catalonia, Spain; Centro de Investigación Biomédica en Red de Salud Mental (CIBERSAM), Instituto de Salud Carlos III, Madrid, Spain

## Abstract

The persistence of genetic variants that increase susceptibility to complex diseases poses an evolutionary paradox: despite their detrimental health effects, these variants are not eliminated by natural selection. Life-history theory proposes that trade-offs and pleiotropic effects across fitness components may explain the evolutionary maintenance of disease-associated alleles. We hypothesize that certain disease-risk alleles might segregate in the population because they confer reproductive advantages, even at the expense of late-life costs on health and longevity. Leveraging genome-wide association studies, we investigated genetic correlations and pleiotropic relationships between 62 complex diseases, longevity, and fertility. In our study, we estimated fertility by meta-analyzing complementary measures of offspring number and used parental lifespan as a proxy for longevity. We found that 85% of diseases showed negative genetic correlations with longevity, whereas 87% of diseases with significant correlations showed positive associations with fertility. Moreover, most disease-risk variants were associated with reduced longevity, even after accounting for socioeconomic confounders. Fertility-increasing alleles exhibited evolutionary signals consistent with adaptive selection despite having pleiotropic effects on disease-risk. Finally, we compared the number of offspring among individuals with high genetic risk of disease and found that, for most diseases, affected individuals had more offspring than disease-free individuals. However, for early-onset conditions, non-affected individuals exhibited higher fertility, highlighting the reproductive cost of early-onset diseases. These findings support the Antagonistic Pleiotropy theory, showing that alleles that enhance early-life reproductive success can persist despite late-life health costs. By uncovering these evolutionary trade-offs between reproduction, longevity, and disease risk, our study shows how Darwinian selection continues to shape contemporary patterns of human disease susceptibility. Understanding these evolutionary trade-offs can inform public health approaches and help anticipate unintended consequences of targeting disease-related genetic pathways.

## Introduction

The human genome carries numerous alleles associated with complex diseases (Welter et al. 2014; Abdellaoui et al. 2023). Despite their detrimental effects on health and survival, these alleles are not purged by natural selection, a longstanding paradox in evolutionary biology (Nesse and Williams 1996). One plausible explanation lies in pleiotropy, the phenomenon where a single genetic variant affects more than one trait, which plays a key role in linking biological systems and evolutionary pressures (Watanabe et al. 2019). Since traits interact rather than evolve in isolation, pleiotropy provides a mechanistic basis for evolutionary trade-offs and may help explain the persistence of disease-risk alleles over generations (Benton et al. 2021).

Life-history theory suggests that organisms allocate limited resources among three primary fitness domains: growth, reproduction and maintenance (Charlesworth 1994; Giaimo and Traulsen 2019) which inherently create trade-offs, particularly between reproduction and longevity, that shape evolutionary outcomes. For instance, experiments in *Drosophila* demonstrated that mutant sterile fruit flies live longer than wild types under controlled conditions (Smith 1958), with similar patterns observed in other taxa (Harshman and Zera 2007). In humans, the evidence is more nuanced. While some studies have failed to identify a clear cost of reproduction on lifespan (Kuningas et al. 2011), demographic analyses across multiple populations have consistently shown an inverse relationship between offspring number and post-reproductive longevity (Westendorp and Kirkwood 1998; Hsu et al. 2021; Long and Zhang 2023) even if socio-economic factors (SEF) can confound this relationship (Lycett, Dunbar, and Voland 2000).

The time of disease onset is a critical factor to understand this evolutionary dynamic, as alleles that manifest after the reproductive years face weaker selective pressures. The Mutation Accumulation (MA) and Antagonistic Pleiotropy (AP) theories offer complementary, and non– mutually exclusive, frameworks for the evolution of genetic influences on longevity and age-related diseases (Medawar 1952; Williams 1957; Kirkwood and Austad 2000). The MA theory proposes that deleterious mutations with late-life effects can accumulate in the population through genetic drift (Medawar 1952); while AP posits that alleles conferring early-life fitness benefits are favored by selection despite late-life costs to health (Williams 1957). Indeed, previous studies have identified hundreds of antagonistic pleiotropy variants and higher allele frequencies and effect sizes for late-onset disease–risk alleles consistent with both theories (Rodríguez et al. 2017; Austad and Hoffman 2018).

A growing body of evidence links genetic susceptibility to common diseases with fundamental fitness traits, particularly longevity and reproduction. For instance, alleles that enhance reproductive success simultaneously increase disease risk or reduce longevity (Mathieson et al. 2023; Long and Zhang 2023), suggesting an explanation for their persistence in human populations. Likewise, cardiovascular disease risk is strongly associated with shortened lifespan (Melzer, Pilling, and Ferrucci 2020; Joshi et al. 2017), while alleles associated with reproductive-related traits have been linked to various adult diseases (Burgess et al. 2017). Despite growing evidence for these trade-offs, a systematic analysis of the shared genetic architecture linking human longevity, fertility, and complex disease is still lacking. Moreover, it remains unclear whether the observed associations reflect deep evolutionary constraints, recent selective pressures, or socioeconomic confounding.

Our study aims to address this gap through a comprehensive analysis of pleiotropic relationships across longevity, fertility (see Glossary) and a wide spectrum of 62 human complex diseases. We defined fertility as reproductive success by meta-analyzing genome-wide association studies (GWAS) of offspring count, and we used a published GWAS of parental lifespan as a proxy for longevity. We integrated cross-trait genetic correlations (see Glossary), pleiotropy detection (see Glossary), signatures of natural selection and Mendelian randomization to test the hypothesis that disease-risk alleles persist in human populations due to concurring pleiotropic effects that increase reproductive fitness. We also leveraged demographic data from the UK Biobank to assess how disease manifestation influences reproductive success. Our findings uncovered shared genetic architecture linking longevity, fertility and disease risk, alongside consistent signatures of recent evolutionary adaptation. These results provide novel insights into evolutionary trade-offs that shape disease susceptibility and life-history traits, with potential implications for personalized medicine and public health strategies.

## Results

### A meta-analysis of fertility GWAS

We conducted a GWAS meta-analysis to quantify the genetic architecture of reproductive success, defined as realized fertility, using three complementary measures: number of children ever born (Barban et al. 2016), number of live births in females (Watanabe et al. 2019), and number of children fathered in males (Watanabe et al. 2019; **Supplementary Fig. 1**). This analysis identified five genome-wide significant loci, corresponding to ten independent SNPs (**Supplementary Table 1)**, all of which overlap with a recent large-scale GWAS on reproductive traits (Mathieson et al. 2023). The strong genetic correlation between our analysis and this previous study (*r_g_*=0.99, SD=7.4e-03) supports the consistency and reliability of our findings. SNP-based heritability of reproductive success was modest but significant (h^2^=0.03, SE=1.4e-03), and MAGMA tissue gene expression analysis revealed significant enrichment in the brain (P=2.3e-04) and ovarian tissues (P=1.4e-03; **Supplementary Figs. 2–3**). To validate the genetic variance in reproductive success identified by our GWAS, we constructed a polygenic score (PGS) and tested its predictive performance on pregnancy-related outcomes in an independent European-ancestry cohort (n=1,005). The PGS significantly predicted the number of live births (NLB, P=0.04) and the number of completed pregnancies (NCP, P=0.04) after adjusting for relevant covariates (**Supplementary Table 2**). Despite small effect sizes, these results show that common variants linked to reproductive success can nonetheless predict individual fertility outcomes, reinforcing the biological validity of the fertility GWAS.

### Disease-risk alleles are genetically linked to reduced longevity but enhanced fertility

We conducted genome-wide genetic correlation analyses to investigate the relationships between 62 complex diseases and two key life-history traits: longevity and fertility. This analysis aimed to test whether disease-associated alleles systematically correlate with reduced lifespan and/or enhanced reproductive success. Among the diseases analyzed, 41 exhibited significant genetic correlations with longevity (P < 0.05). Notably, 85% of these (35 out of 41) demonstrated negative correlations, indicating that individuals with greater genetic susceptibility to disease tend to have shorter lifespans (**Fig. 1a**, **Supplementary Table 3**). The strongest negative correlations were observed for circulatory system diseases, including heart failure (*r_g_*=-0.74, P=5.32e-67), hypertension (*r_g_*=-0.69, P=1.39e-145), myocardial infarction (*r_g_*=-0.66, P=1.38e-81) and coronary artery disease (*r_g_*=-0.65, P=6.52e-59). Only six diseases showed positive genetic correlations with longevity (P<0.05), including age-related hearing impairment (*r_g_*=0.20, P=2.4e-03) and melanoma (*r_g_*=0.17, P=3.9e-03), among others. The domains most strongly associated with reduced longevity were the circulatory, endocrine and metabolic, and musculoskeletal systems (**Supplementary Fig. 4**). Notably, diseases with later onset exhibited stronger negative genetic correlations with longevity (r=–0.43, P=5e-03, **Supplementary Fig. 5** and **Supplementary Table 3**), consistent with the evolutionary prediction that genetic variants with post-reproductive effects are more likely to persist across generations despite their health-related costs.

**Fig. 1.**
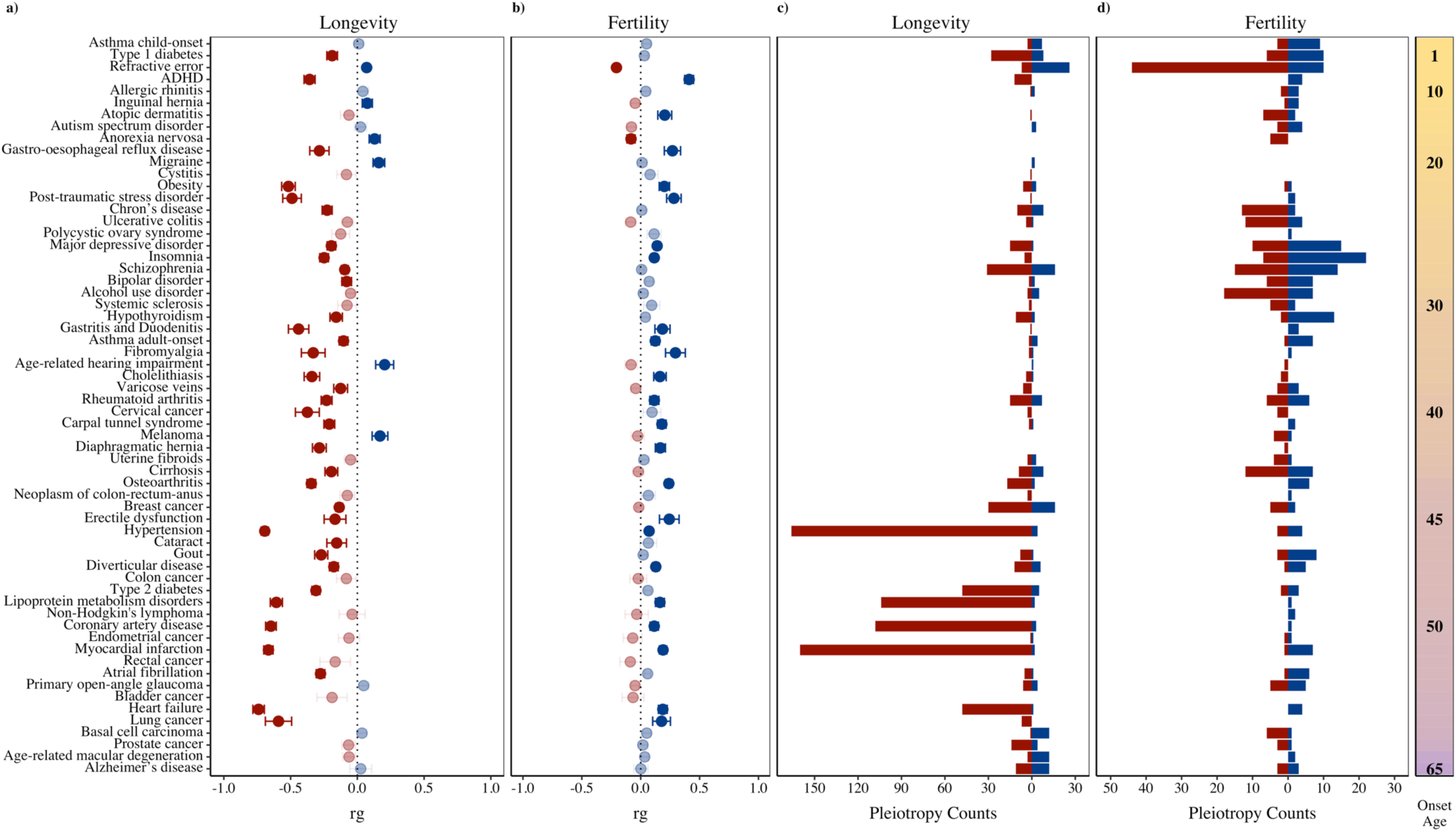
Genetic correlations and pleiotropies between complex diseases and life-history traits. **a-b**, global genetic correlations (*rg*) for the 62 complex diseases with longevity and fertility, respectively, with error bars indicating standard errors. **c-d**, number of pleiotropic SNPs for the 62 complex diseases with longevity and fertility, respectively. Diseases are sorted in ascending order of age at disease onset as represented in the last panel, starting with the most early-onset disease. Positive pleiotropies effects are depicted in blue, while negative pleiotropic effects are shown in red (see Glossary). **a-b**, color intensity corresponds to statistical significance, with darker hues indicating P-value < 0.05.

In sharp contrast, fertility exhibited predominantly positive genetic correlations with disease risk. Out of the 30 diseases with a significant correlation (P<0.05), 26 (87%) were positive, suggesting that alleles promoting reproductive success may simultaneously increase disease risk. (**Fig. 1b, Supplementary Table 3**). The strongest associations were observed for ADHD (*r_g_*=0.41, P=1.1e-27), fibromyalgia (*r_g_*=0.29, P=4e-04) and post-traumatic stress disorder (*r_g_*=0.28, P=4.59e-06). In this case, no association was found between correlation strength and the age at disease onset (r=-0.01, P=9.5e-01, **Supplementary Fig. 6**). Domain-specific analyses showed stronger associations with increased fertility in the genitourinary, musculoskeletal and digestive systems (**Supplementary Fig. 7**). These patterns remained statistically robust after multiple-test correction (FDR<0.05; **Supplementary Fig. 8-10**).

### Pleiotropic loci shared between fitness components and complex diseases

To further dissect the shared genetic basis underlying the observed trade-offs, we identified pleiotropic loci jointly associated with both complex diseases and life-history traits. We identified 1,142 independent shared SNPs (conjFDR<0.05, **Fig. 1c**, **Supplementary Table 4**) jointly associated with our set of complex diseases and longevity. In line with the observed genetic correlations, 82% (n=942) of these loci showed negative pleiotropy (see Glossary), which indicates that alleles increasing disease risk were also associated with reduced lifespan, consistent with the concept of evolutionary cost. These pleiotropic relationships were especially prevalent in diseases of the circulatory system (**Supplementary Fig. 11**): hypertension (n=170), myocardial infarction (n=162), and coronary artery disease (n=111). Notably, 25% (n=191) of SNPs displayed pleiotropic associations across multiple diseases pinpointing potential genetic hotspots (**Supplementary Table 4**). For example, rs17696736 was pleiotropic across 12 diseases. Functional annotation of the 750 unique pleiotropic SNPs showed enrichment in intronic regions (40.3%) a pattern previously observed in studies of complex trait genetics (Watanabe et al. 2019), **Supplementary Figures 12-14**). In addition, negative pleiotropies were up-regulated in the colon, and down-regulated in the cervical spinal cord and substantia nigra (**Supplementary Figures 15-16**). Over-representation analysis revealed that negative pleiotropy is mediated by genes that elevate disease risk and shorten lifespan, with significant enrichment in lipid metabolism pathways (e.g., cholesterol, FDR=4.17e-04). Conversely, positive pleiotropies were enriched for histone genes involved in chromatin structure (FDR=2.22e-04, **Supplementary Table 5**).

On the other hand, we identified 460 independent SNPs (conjFDR<0.05) jointly associated with complex disease and fertility (**Fig. 1d**, **Supplementary Table 6**). Of these, 229 loci (49.8%) exhibited positive pleiotropy (see Glossary), whereas for 231 loci (50.2%) the direction of the effects was negative. This nearly even split highlights the complexity of evolutionary forces shaping fertility-related disease alleles. Mental, behavioral, and neurodevelopmental domains showed the highest genetic overlap with fertility (23.7%, n=109), followed by the digestive system (14.8%, n=68; **Supplementary Fig. 17**). A substantial subset of these SNPs (26%, n=74) exhibited pleiotropy between multiple diseases and fertility (**Supplementary Table 6**). Functional characterization of the 285 unique pleiotropic SNPs between complex diseases and fertility revealed enrichment in intronic regions (40.5%), as well as upstream (24.7%) and downstream (15.2%) of genes (**Supplementary Fig. 18**). These SNPs, predominantly mapped to protein-coding genes (73.6%) or long intergenic non-coding RNAs (lincRNAs, 8.5%; **Supplementary Fig. 19**).

### Antagonistic pleiotropy links higher fertility to reduced longevity

Building on the genetic correlation and pleiotropy findings, we investigated the shared genetic architecture between longevity and fertility. First, we described a modest and significant negative genetic correlation between both key life-history traits (r_g_=-0.10, P=2.4e-03). This inverse relationship suggests a trade-off whereby alleles that increase reproductive output may compromise lifespan, which is consistent with theoretical predictions and empirical observations in many species (Charlesworth 1994). For the first time, eight shared genetic variants with effects on both phenotypes were identified with pleioFDR (**Supplementary Table 7**). Notably, seven of these variants displayed direction of effects concordant with the Antagonistic Pleiotropy theory of aging (see Glossary), wherein the allele that enhances fertility was associated with reduced longevity (Williams 1957). Most of these variants were intronic and none of them have previously been linked to fertility. These findings provide direct variant-level support for the theory that reproduction-enhancing alleles incur biological costs later in life. Mapped lincRNAs showed tissue-specific expression in the testes and brain, suggesting potential relevance to reproductive timing and neuroendocrine regulation (**Supplementary Fig. 20**). In contrast, only one variant showed the same direction of effect on both traits: an intronic SNP in *WNT3*, a gene essential for embryonic development and germ cell maturation (see Supplementary Material).

### Fertility-enhancing alleles show signatures of recent positive selection despite pleiotropic effects on disease

All pleiotropic variants were annotated based on the disease-risk allele and classified according to their effects on fitness: positive pleiotropy (blue in the figures) denoted SNPs where the risk allele was associated with increased longevity or fertility, while negative pleiotropy (red) referred to SNPs where the risk allele was linked to reduced longevity or fertility. We then investigated whether alleles that enhance fitness -either through increased longevity or fertility-have been favored by natural selection despite their detrimental effects on disease risk. To test this, we compared measures of recent positive selection, including the Singleton Density Score (SDS, (Field et al. 2016) and the integrated-Haplotype Score (iHS, (Voight et al. 2006), between pleiotropic variants exhibiting positive versus negative pleiotropy.

SDS infers positive selection by linking singleton depletion to rising allele frequencies, providing unprecedented resolution for recent human adaptation. In essence, positive SDS values indicate alleles that have recently increased in frequency (∼75–100 generations). Although most pleiotropic variants exhibited moderate absolute SDS scores, selection patterns varied depending on whether the risk allele enhanced or reduced fitness traits (**Supplementary Tables 4 and 6**). Disease-risk alleles associated with increased fertility showed significantly higher positive SDS values compared to those linked to reduced fertility, indicating a trend toward recent positive selection (**Fig. 2**). Stratification by age at disease onset revealed that this pattern was consistently for both early- and late-onset diseases (**Fig. 2a**). The significant disparity between fertility-increasing and fertility-decreasing risk alleles (Mann–Whitney P=2.7e-04) suggests that the reproductive advantages conferred by these variants may outweigh their disease-associated costs (**Fig. 2b**). This supports the idea that reproductive success, and not disease avoidance, is the primary target of recent natural selection. Permutation-based resampling confirmed the robustness of this finding, with the observed p-value falling within the lowest 5% of the empirical null distribution (**Supplementary Fig. 21**), providing robust evidence for differential selective pressures favoring fertility-increasing alleles. Consistently, fertility-increasing alleles exhibited significantly higher SDS values than disease-risk alleles (Mann–Whitney P = 2.5e-03; **Fig. 2c**). In contrast, there was no enrichment of positive iHS values for fertility-increasing alleles (P=2.4e-01), suggesting that selection pressures might be recent rather than ancient, as iHS captures older selection signals extending up to ∼30,000 years. Similarly, pleiotropic SNPs associated with longevity showed no significant differences in SDS (P=5.07e-02) or iHS (P=1.07e-01) between alleles that increased versus decreased lifespan, indicating a lack of recent selective pressures favoring longevity-related variants (see Supplementary Material). Notably, pleiotropic alleles that enhance fertility tended to exhibit antagonistic effects on longevity (one sample, one-tailed t-test P = 3e-10; **Fig. 2d**). This trade-off likely reflects the weaker selective pressures acting on late-life traits with limited impact on reproductive fitness. Taken together, these findings show that natural selection continues to shape human genomes by prioritizing reproductive advantages, even when they come at the cost of long-term health.

**Fig. 2.**
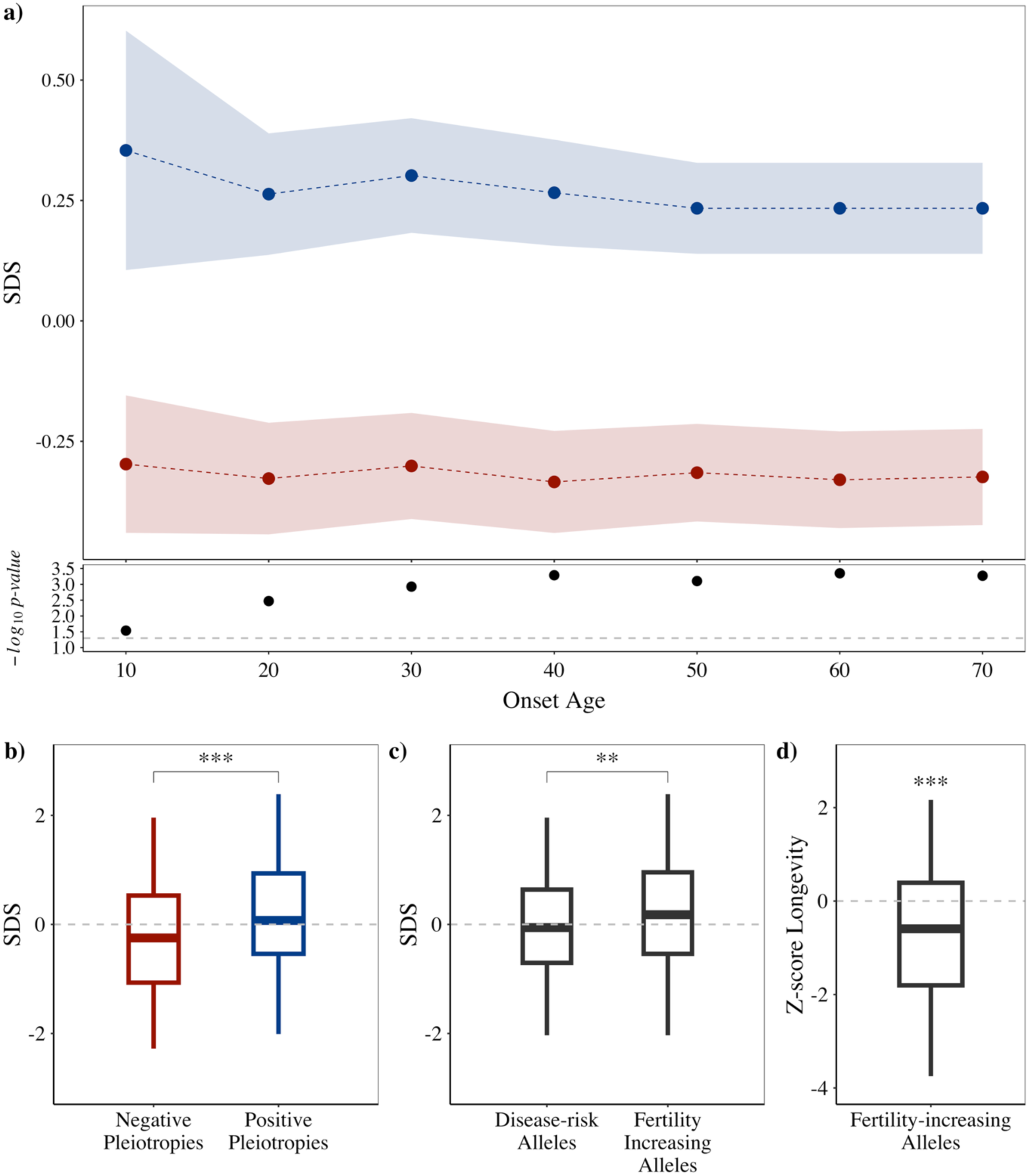
Evolutionary signals towards positive selection on fertility-increasing alleles. We classified pleiotropic variants by the effect of the disease-risk allele on fertility: positive pleiotropy (blue) when the risk allele associates with increased fertility, and negative pleiotropy (red) when it associates with decreased fertility. **a**, Cumulative Singleton Density Score (SDS) across age onset categories for positive and negative pleiotropies. Inset shows log₂-transformed p-values from Mann–Whitney U tests at each onset age. **b–d**, Boxplots with the interquartile range (IQR) and median; whiskers extend to 1.5×IQR. Significant Mann–Whitney U test p-values are indicated (*p < 0.05, **p < 0.01, ***p < 0.001). **b**, SDS values for disease-risk alleles grouped by pleiotropy type (Mann–Whitney p = 2.7 × 10⁻⁴, ***). **c**, SDS values for the disease-risk allele versus the fertility-increasing allele within the set of pleiotropic SNPs (Mann–Whitney p = 2.5 × 10⁻³, **). **d**, GWAS-estimated effect sizes for longevity (Z-scores) among the 285 unique fertility–disease pleiotropies. A one-sample, one-tailed Student’s t-test tested whether the mean longevity effect of fertility-increasing alleles is below zero (H₀: μ = 0; H₁: μ < 0; p = 3 × 10⁻¹⁰, ***).

### Mendelian randomization supports a directional link from disease risk to reduced longevity

We used two-sample Mendelian randomization to explore directional links between disease risk and life-history traits. First, we applied univariable MR (UVMR) to estimate total effects, then multivariable MR (MVMR) accounting for socioeconomic factors. Among the 62 diseases analyzed, 22 (35%) showed a statistically significant inverse association with longevity in UVMR (P<0.05), consistent with a causal effect of disease liability on shorter lifespan **(Supplementary Fig. 22** and **Table 8**). Cardiovascular diseases exhibited the strongest evidence of an adverse effect on lifespan, including hypertension (OR=0.62, 95% CI: 0.59-0.65), coronary artery disease (OR=0.63, CI: 0.55-0.71), myocardial infarction (OR=0.65, CI: 0.59-0.70), and heart failure (OR=0.72, CI: 0.60-0.83). Notably, 19 of the 22 associations remained significant after accounting for socioeconomic factors in MVMR, with most surviving FDR correction (**Supplementary Fig. 23** and **Table 9**). These results indicate that the observed longevity reduction is not simply attributable to confounding by socioeconomic status. Reverse analyses assessing the effect of longevity on disease risk were not performed, as such directionality is biologically implausible.

We then examined whether genetic predisposition to fertility might be linked to disease-risk. Eight traits showed nominal associations in UVMR, including a reduced risk for schizophrenia (OR=0.49, CI: 0.29-0.85) and breast cancer (OR=0.51, CI: 0.30-0.85), and an increased risk for atrial fibrillation (OR=1.91, CI: 1.19-3.05; P<0.05; **Supplementary Fig. 24a** and **Table 10**). However, these estimates did not survive multiple testing corrections (FDR>0.05) and may be affected by the small number of genome-wide significant variants available for fertility (n=5, **Supplementary Fig. 25a** and **Table 11**). Such limited statistical power cautions against strong causal interpretation of these results.

Finally, reverse MR analyses were conducted to assess whether genetic liability to complex diseases influences fertility. Modest associations were identified for nine conditions (P<0.05, **Supplementary Fig. 24b** and **Table 12**). Genetic risk for ADHD was associated with a slight increase in fertility (OR=1.08, CI: 1.04–1.12), whereas melanoma risk showed an inverse association (OR=0.97, CI: 0.96–0.98). Interestingly, melanoma risk was also positively associated with longevity (**Supplementary Fig. 22**), suggesting a genetic architecture consistent with the Antagonistic Pleiotropy theory. Most reverse associations remained significant after adjustment for SEF and correction for multiple testing (**Supplementary Fig. 25b** and **Table 13**).

Together, MR analyses indicate that genetic correlations between disease risk, longevity, and fertility reflect, at least in part, causal, directional relationships. While limitations persist— particularly weak instruments for fertility and potential residual confounding—our results support a model whereby disease-associated alleles influence life-history traits.

### Age of disease onset modulates the reproductive impact of genetic risk

To investigate how the timing of disease manifestation modulates the relationship between genetic risk and reproductive outcomes, we analyzed fertility patterns in UK Biobank participants stratified by both disease status and polygenic risk. For each of the 62 complex diseases, we compared the number of offspring among individuals in the top decile of genetic risk, distinguishing between those with a confirmed diagnosis and those who remained undiagnosed. This design allowed us to assess whether subclinical individuals (those carrying a high genetic liability but without disease manifestation), might experience a reproductive advantage compared to individuals who develop the disease.

Significant reproductive differences were observed for 52 of the 62 diseases examined. In 41 diseases (79%), individuals with a clinical diagnosis exhibited higher average fertility than their undiagnosed counterparts. Conversely, for the 11 diseases where diagnosed individuals had lower fertility, the majority (73%, n=8) had an onset before age 20. These early-onset conditions were associated with greater fertility among high disease-risk but unaffected individuals. In contrast, late-onset diseases, typically emerging after the reproductive years, were associated with increased fertility in diagnosed individuals, reinforcing the idea that natural selection acts most strongly on early-life traits (**Fig. 3**). Notably, after age 45— reflecting the end of the reproductive window—all diseases are linked to higher fertility among diagnosed individuals, aligning with established patterns of senescence (Rodríguez et al. 2017). This suggests that early-onset diseases impose direct reproductive costs, whereas late-onset diseases may allow individuals to benefit from the fertility-enhancing effects of risk alleles before symptoms emerge.

**Fig. 3.**
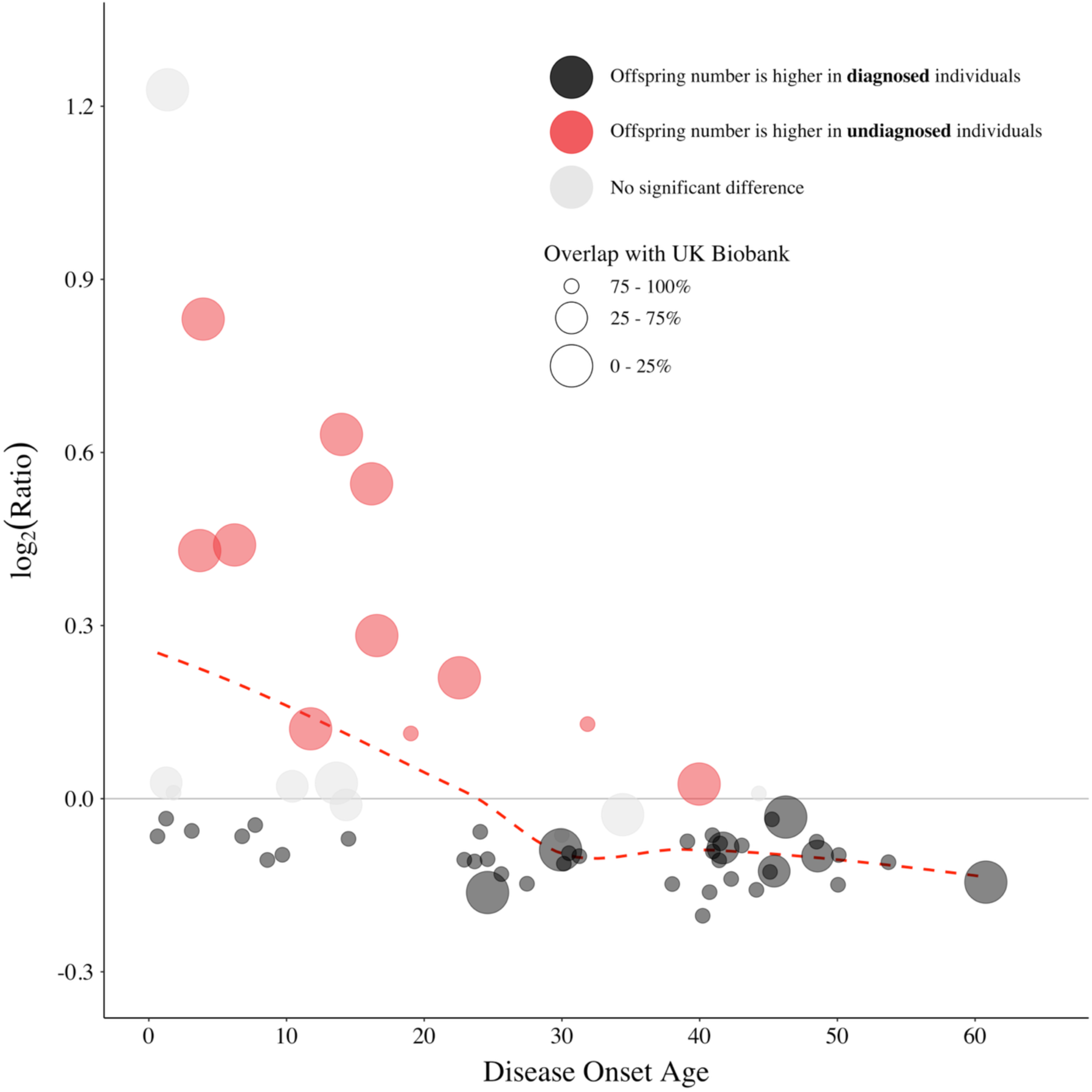
Age at disease onset shapes fertility outcomes. Log₂ ratio of mean offspring count (undiagnosed/diagnosed) among individuals in the top decile of genetic liability for 62 complex diseases. Diseases are ordered on the x-axis by their typical age at onset. Positive values (red) indicate higher fertility in undiagnosed individuals; negative values (black) indicate higher fertility in diagnosed individuals. Error bars—standard errors of the log₂ ratios estimated via the delta method—are omitted for clarity. Ten diseases whose confidence intervals include zero are shaded in gray, indicating no significant difference. Dot size is inversely proportional to sample overlap with the UK Biobank.

### Discussion: evolutionary trade-offs shape the genetic architecture of disease, fertility, and lifespan

By examining the pleiotropic architecture underlying key human life-history traits and complex diseases, we demonstrate a genetic trade-off between longevity and fertility, a relationship that also extends to their shared genetic links with disease. Indeed, increased disease risk is often linked to a shorter lifespan and, surprisingly, higher fertility. We show that these genetic variants have been favored by natural selection due to their reproductive benefits, despite their detrimental effects on health, particularly when those effects emerge after reproductive age. Demographic analysis corroborates this interpretation by showing that the effects of disease-risk alleles on fertility vary with age of disease onset. Together, these findings shed light on the fundamental evolutionary trade-offs between human longevity and reproduction, providing strong and multi-level support for the Antagonistic Pleiotropy theory of senescence in humans.

First, our meta-analysis of offspring count confirmed that fertility is a polygenic trait with modest yet significant heritability and can be reliably predicted using GWAS-derived polygenic scores. The polygenic architecture of fertility was biologically plausible, showing enrichment for gene expression in brain and ovarian tissues, which is consistent with known roles (Watanabe et al. 2017; Leeuw et al. 2015). Notably, one of the lead SNPs mapped to the ESR1 gene, which has previously been associated with endometriosis and infertility (Paskulin et al. 2013; Ge et al. 2014; Benonisdottir et al. 2024). Furthermore, our findings showed strong genetic correlations with prior GWASs of related reproductive traits (Mathieson et al. 2023), supporting the robustness and validity of our approach, and providing evidence for the shared genetic basis of reproductive traits.

Second, genetic correlation analyses revealed that increased risk of disease is generally associated with fertility. This suggests that genetic factors that promote reproductive success often increase disease susceptibility. Notably, these correlations were strongest for early-onset disorders, and most pleiotropic associations with fertility were concentrated among diseases that manifest early in life. This emphasizes the complex trade-offs between reproductive advantages and increased disease liability (Stearns 1989; Gagnon et al. 2009; Crespi 2010; Abrams and Miller 2011; Aktipis et al. 2013). The most pronounced positive correlations with fertility were observed within the psychiatric domain, which aligns with previous evidence linking traits such as creativity and sociability to increased reproductive success (Nettle and Clegg 2005). In contrast, most complex diseases were negatively correlated with longevity, consistent with earlier findings (Timmers et al. 2019; Deelen et al. 2019; Hu et al. 2022) and supporting the idea that disease risk undermines lifespan through shared biological pathways.

At the variant level, pleiotropic SNPs generally recapitulated these genome-wide correlations, particularly regarding longevity, where negative pleiotropic effects were more frequently observed among late-onset diseases. However, many loci exhibited effects that diverged from expectations based on simple genetic correlations, reflecting the nuanced and multifaceted nature of the relationship between genetic traits and disease liability (Williams 1957; Wu et al. 2022; Kuningas et al. 2011). We also identified a modest but significant negative genetic correlation between fertility and longevity. This finding is consistent with the long-standing trade-off between reproduction and lifespan (Charlesworth 1994), which supports the antagonistic pleiotropy theory of ageing (Pettay et al. 2005; Tabatabaie et al. 2011; Long and Zhang 2023).

Compared to disease-risk alleles with detrimental effects on fertility, disease-risk alleles with fertility-enhancing pleiotropic effects exhibit suggestive signatures of positive selection over the past ∼2,000 years. In contrast, disease-risk alleles associated with longevity did not show comparable evidence of positive selection. These patterns show that selection is driven mostly by fertility rather than by health, illustrating how early-life reproductive advantages offset health costs that may lead to shorter lifespans (Tropf et al. 2015; Rodríguez et al. 2017; Long and Zhang 2023; Gao 2024). These findings also illustrate a fundamental asymmetry in the evolution of life-history traits: genetic variants that influence traits early in life are more likely to be shaped by natural selection due to their stronger impact on reproductive fitness (Liu et al. 2024; Benonisdottir et al. 2024). Interestingly, the modern data we used demonstrates that these evolutionary dynamics are still evident after the demographic transition, when fertility rates declined and reproductive behavior changed (Frejka 2016). This highlights the lasting impact of natural selection on the human genome (Nesse and Williams 1996; Jensen et al. 2015; Hsu et al. 2021).

Hormonal pathways, particularly those involving insulin and insulin-like growth factor 1 (IGF-1), have long been associated with the trade-off between reproduction and longevity in model organisms (Dillin, Crawford, and Kenyon 2002). However, the underlying pathways in humans have not been exhaustively identified. For the first time, we have identified eight pleiotropic loci that affect both fertility and longevity. Notably, seven of these loci displayed antagonistic pleiotropy, whereby alleles that enhance fertility were associated with reduced longevity. Functional annotation of these variants revealed significant enrichment in regulatory elements, consistent with previous studies indicating that antagonistic pleiotropic variants often reside in non-coding regions and play a role in regulating multiple genes or tissues (Long and Zhang 2023). These regulatory features may enable small, coordinated changes in gene expression that mediate complex trade-offs across life-history traits.

Overall, MR analyses indicated that genetic liability to complex diseases is causally associated with reduced longevity, supporting the notion that disease-risk alleles have a causal influence on lifespan. While previous studies have suggested that socioeconomic inequality affects longevity (Ye et al. 2023), we found that the associations between diseases and longevity persisted even after adjusting for socioeconomic factors. This suggests that the observed effects are not solely attributable to environmental or cultural confounders and provides a key validation of the evolutionary interpretation from a causal inference framework. In contrast, the results obtained for the association between genetic liability to complex diseases and fertility were more variable, reflecting the complexity of reproductive traits and of their interplay with disease risk, as well as the potential limitations of the genetic instruments available for fertility-related outcomes. Future studies with stronger instruments for fertility and sex-stratified analyses may help clarify these relationships.

Finally, our findings reveal that the influence of genetic disease risk on fertility is modulated by the timing of disease onset as expected according to the Antagonistic Pleiotropy theory of senescence. Specifically, individuals with high genetic predisposition to late-onset diseases tend to have more children. This suggests that alleles that predispose individuals to diseases that manifest after reproductive age may confer reproductive advantages earlier in life. Moreover, individuals who carry disease-risk alleles for early-onset conditions but remain disease-free during their reproductive years, also exhibit higher fertility rates. This implies that the absence of disease manifestation during critical reproductive periods enables carriers to benefit from the fertility-enhancing effects of these alleles without incurring the associated health costs. This is consistent with previous studies in which the age at disease onset influences childlessness (Liu et al. 2024). These age-sensitive trade-offs may constitute a key mechanism by which natural selection maintains disease-risk variants in human populations.

Our study has several limitations that warrant consideration. First, while similar evolutionary patterns have been documented in historical cohorts across the epidemiological transition, our analyses rely on individuals of European ancestry, which may limit generalizability to other populations (Kaptijn et al. 2015; Morita 2018). Second, although aggregating data from both sexes increased statistical power, the current lack of sex-stratified data prevents us from uncovering sex-specific associations and gene–sex interactions (Jensen et al. 2015; Bernabeu et al. 2021; Gardner et al. 2022). Third, while MR strengthens causal inference, our findings remain vulnerable to limitations in instrument strength, especially for fertility-related traits, due to modest SNP-based heritability and to residual confounding by cohort-specific factors (Kaptijn et al. 2015; Hsu et al. 2021). Future work should integrate more diverse biobank resources and explicitly model gene–environment and gene–sex interactions to test the robustness and broader applicability of these evolutionary trade-offs.

Together, our findings reveal a nuanced interplay between genetic risk, disease onset, and reproductive timing, uncovering age-dependent evolutionary trade-offs that help explain the persistence of disease-associated variants in human populations (Nesse and Williams 1996). These results provide strong empirical support for the Antagonistic Pleiotropy theory of senescence (Williams 1957). Also, by demonstrating how the timing of trait expression modulates selection pressure, favoring variants that enhance early-life fertility despite late-life health costs, our study highlights a fundamental mechanism shaping human health. These insights not only deepen our understanding of the evolutionary forces shaping human health but also highlight the importance of incorporating evolutionary perspectives into biomedical research. Doing so may improve the interpretation of polygenic scores and inform interventions that aim to reduce disease burden without inadvertently compromising reproductive health or fitness-related traits.

## Methods

### Genome-wide association study (GWAS) meta-analysis on fertility

We conducted a GWAS meta-analysis on reproductive success using three complementary measures: number of children ever born (Barban et al. 2016), number of live births in females (Watanabe et al. 2019), and number of children fathered in males (Watanabe et al. 2019). The meta-analysis was performed using the software METAL (Willer, Li, and Abecasis 2010). The analysis employed a sample size weighting scheme to account for varying sample sizes across input studies (**Supplementary Table 14**) and applied overlap correction to address shared participants, primarily from the UK Biobank. To further reduce potential biases, genomic control was implemented to correct for population stratification. Additional methodological details are provided in Supplementary Section 1.

### GWAS summary statistics

We assembled a dataset of 62 GWAS summary statistics covering complex diseases across 12 major clinical domains, sourced from publicly available repositories (Watanabe et al. 2019; Sullivan 2010; Cerezo et al. 2025). To ensure quality and comparability, we selected datasets that met the following criteria: studies in individuals of European ancestry (>95%), with a total sample size ≥20,000, at least 2,000 cases, and SNP_h²_ ≥ 2%. To standardize phenotypes, we used the International Classification of Diseases, 10th revision (ICD-10, (World Health Assembly 1990), which provides a hierarchical framework and universally recognized system for categorizing medical conditions (**Supplementary Table 14**).

Longevity data was obtained from Timmers et al. 2019, which estimated genetic effects on lifespan using nearly one million parental lifespans from ∼450,000 UK Biobank participants of European ancestry. For fertility, we meta-analyzed three independent GWASs using the number of children as a proxy for reproductive success, given the challenges in directly quantifying reproductive potential (**Supplementary Material**).

GWAS summary statistics were standardized using a reference set of 9,546,816 SNPs from the 1000 Genomes Project (https://www.internationalgenome.org/). We removed non-biallelic variants, SNPs without SNP ID (rsIDs), duplicates, and strand-ambiguous alleles (A/T or G/C). When available, SNPs with imputation quality (INFO) scores < 0.9 were excluded. All datasets were aligned to hg19 genome build including only autosomal SNPs. Allele frequencies were taken from original studies or calculated using European samples in the 1000 Genomes reference panel. Finally, only SNPs with a minor allele frequency (MAF) > 0.01 were retained.

### Age at disease onset

Age at onset for 62 complex diseases was estimated using prevalence data from the Risteys FinRegistry Release 11 (Viippola et al. 2023), a population-based dataset representing individuals of European ancestry (**Supplementary Table 14; Supplementary Figs. 26-27**). Onset age was operationally defined as the maximum age within the youngest 5% of affected individuals, providing a conservative threshold for early manifestation (Jia et al. 2019). Further details are available in Supplementary Methods Section 2.2.

### Heritability and genetic correlations

Bivariate LD score regression (LDSC; Bulik-Sullivan et al. 2015) was used to estimate pairwise genome-wide genetic correlations (*r_g_*) between fertility and longevity, as well as the set of 62 complex diseases, as well as SNP heritability for all traits. We retained the original number of SNPs in all datasets. We evaluated potential GWAS-related confounding by calculating Pearson correlations between r_g_ and key study characteristics (e.g., heritability, sample size). To identify influential outliers, we conducted a disease-wise bootstrap analysis. Genetic correlation effect sizes correlated mainly with sample size (r=-0.32, P=0.012, **Supplementary Fig. 28**) and the number of genome-wide significant SNPs (r=-0.32, P=0.012, **Supplementary Fig. 29**) in complex disease GWASs, capturing the influence of statistical power on the results. See Supplementary Methods Section 2.3.

### Pleiotropy

We employed the genetic pleiotropy-informed conjunctional false discovery rate (conjFDR) method (Andreassen et al. 2013) to identify pleiotropic SNPs affecting both disease risk and fitness components. The analysis was performed on GWAS summary statistics for 62 complex diseases, longevity, and fertility, using a common set of 2,241,316 SNPs. Pleiotropic SNPs (conjFDR<0.05) were classified as positive or negative based on their effects on the fitness trait (See Glossary Box). Pleiotropy counts correlated with the number of disease cases (r=0.36, P=0.005, **Supplementary Fig. 30**), the total number of genome-wide significant SNPs (r=0.69, P=5.9e-10) and the SNP_h2_ (r=0.5, P=3.5e-5, **Supplementary Fig. 31**). See Supplementary Methods Section 2.4.

### Functional annotation

We mapped pleiotropic SNPs (conjFDR<0.05) by physical proximity (5000bp) and performed gene-based annotation of canonical transcripts to identify functional consequences of SNPs with Ensembl Variant Effect Predictor (VEP, http://grch37.ensembl.org/Tools/VEP). We performed enrichment analysis via FUMA’s GENE2FUNC module to elucidate the molecular pathways implicated by these loci (Watanabe et al. 2017). The Benjamini-Hochberg procedure controlled the false discovery rate, and we used the common SNPs gene set as the background reference (**Supplementary Fig. 32**). See Supplementary Methods Section 2.5.

### Evolutionary analysis of pleiotropic SNPs

To investigate whether positive and negative pleiotropies associated with complex diseases exhibit distinct patterns of positive selection, we analyzed evolutionary signatures in disease-risk alleles identified as independent pleiotropic SNPs. We hypothesized that alleles conferring fitness advantages—either by enhancing longevity or fertility—would display stronger signals of positive selection compared to those associated with reduced fitness, such as decreased longevity or fertility. To test this hypothesis, we used two established indices of positive selection: the integrated haplotype score (iHS; Voight et al. 2006), and the singleton density score (SDS; Field et al. 2016). To validate the results, we conducted a leave-one-out analysis for each disease and performed bootstrapping through resampling with replacement (**Supplementary Table 15**). See Supplementary Methods Section 2.6.

### Mendelian randomization

Using independent genome-wide significant SNPs as instrumental variables, we applied univariate bi-directional MR with the inverse-variance weighted (IVW) method via the MendelianRandomization R package (Burgess et al. 2024). For significant causal estimates, we further conducted multivariable MR analyses to incorporate socioeconomic factors as potential mediators (**Supplementary Fig. 33**). To ensure the robustness of our findings, we employed sensitivity analyses using Cochran’s Q statistic to assess heterogeneity among instrumental variables and the MR-Egger intercept and MR-PRESSO to evaluate potential pleiotropy (Verbanck et al. 2018). Our findings focused on effect estimates and directions rather than definitive causal conclusions. See Supplementary Methods Section 2.7.

### Demography in the UK Biobank

To investigate how disease status contributes to the positive genetic correlation between fertility and disease susceptibility, we computed disease polygenic scores and analyzed fertility outcomes in the UK Biobank (Sudlow et al. 2015). Linear regression models, adjusted for confounders and socioeconomic factors, indicated that education and income explained less than 1% of PGS variance. We stratified individuals by genetic risk, focusing on the top 10% (highest PGS category), and compared fertility outcomes between this group across diseases (**Supplementary Table 16**). PGS calculations used default settings with cross-validation and Bayesian model selection to include only independent, significant variants. See Supplementary Methods Section 2.8.

## Data Availability

All data produced in the present study are available upon reasonable request to the authors

## Glossary Box

- Reproduction is a fundamental component of Darwinian fitness, reflecting an individual’s potential to contribute offspring to future generations. In this study, we use the term **fertility** to denote this reproductive success, measured through realized offspring count. In our GWAS we meta-analyzed three complementary traits: the number of live birth pregnancies, the number of children fathered in males and the number of children ever born to females and males.
- **Genetic correlation** quantifies the proportion of additive genetic variance shared between two traits, offering a global measure of their genetic overlap. Its magnitude reflects the strength of this shared influence, and its sign indicates whether alleles that increase one trait tend to increase or decrease the other. LD score regression (LDSC) estimates these correlations by leveraging linkage disequilibrium patterns and using GWAS summary statistics. Although genetic correlation does not prove causation, it often signals underlying pleiotropy between traits, especially when supported by biological plausibility or functional annotation.
- **Pleiotropy** occurs when a single genetic variant influences multiple phenotypic traits, potentially mediating trade-offs between them. To identify independent pleiotropic single nucleotide polymorphisms (SNPs) between two traits, we utilized the pleioFDR framework. This approach enhances detection power by leveraging shared genetic signals from GWAS summary statistics and rigorously controls the false discovery rate through integrated conditional and conjunctional FDR methods. As a result, we can prioritize SNPs with validated multi-trait associations while minimizing spurious findings.
- **Positive pleiotropy** denotes SNPs with concordant allelic effects between two traits. For instance, an allele that increases the risk of a disease may also be associated with high risk for other diseases or with higher fertility. In the context of longevity, an allele increasing disease risk is associated with enhanced longevity. In this study, positive pleiotropy is consistently indicated in blue.
- **Negative pleiotropy** refers to SNPs with opposing allelic effects between two traits. For example, an allele that increases disease risk can simultaneously reduce fertility or be associated with shorter lifespan. In this study, negative pleiotropy is consistently depicted in red.
- **Antagonistic Pleiotropy** is a specific form of pleiotropy particularly relevant to longevity research, in which the same allele confers an advantage early in life but incurs a health cost later in life. First proposed by Williams in 1957 as part of his theory on the evolutionary origins of senescence, it describes that alleles conferring early-life benefits—when natural selection is strong—may be favored despite their detrimental effects later in life, when selection pressures wane.

## Acknowledgements

This research has been conducted using the UK Biobank Resource under application number 67292. We also utilized data from dbGaP study phs000289 under accession number 33199.

## References

Abdellaoui, Abdel, Loic Yengo, Karin J. H. Verweij, and Peter M. Visscher. 2023. “15 Years of GWAS Discovery: Realizing the Promise.” American Journal of Human Genetics 110 (2): 179–94. 10.1016/j.ajhg.2022.12.011.

Abrams, Elizabeth T., and Elizabeth M. Miller. 2011. “The Roles of the Immune System in Women’s Reproduction: Evolutionary Constraints and Life History Trade-Offs.” American Journal of Physical Anthropology 146 (S53): 134–54. 10.1002/ajpa.21621.

Aktipis, C. Athena, Amy M. Boddy, Robert A. Gatenby, Joel S. Brown, and Carlo C. Maley. 2013. “Life History Trade-Offs in Cancer Evolution.” Nature Reviews Cancer 13 (12): 883–92. 10.1038/nrc3606.

Andreassen, Ole A., Wesley K. Thompson, Andrew J. Schork, Stephan Ripke, Morten Mattingsdal, John R. Kelsoe, Kenneth S. Kendler, et al. 2013. “Improved Detection of Common Variants Associated with Schizophrenia and Bipolar Disorder Using Pleiotropy-Informed Conditional False Discovery Rate.” PLOS Genetics 9 (4): e1003455. 10.1371/journal.pgen.1003455.

Austad, Steven N, and Jessica M Hoffman. 2018. “Is Antagonistic Pleiotropy Ubiquitous in Aging Biology?” Evolution, Medicine, and Public Health 2018 (1): 287–94. 10.1093/emph/eoy033.

Barban, Nicola, Rick Jansen, Ronald de Vlaming, Ahmad Vaez, Jornt J. Mandemakers, Felix C. Tropf, Xia Shen, et al. 2016. “Genome-Wide Analysis Identifies 12 Loci Influencing Human Reproductive Behavior.” Nature Genetics 48 (12): 1462–72. 10.1038/ng.3698.

Benonisdottir, Stefania, Vincent J. Straub, Augustine Kong, and Melinda C. Mills. 2024. “Genetics of Female and Male Reproductive Traits and Their Relationship with Health, Longevity and Consequences for Offspring.” Nature Aging 4 (12): 1745–59. 10.1038/s43587-024-00733-w.

Benton, Mary Lauren, Abin Abraham, Abigail L. LaBella, Patrick Abbot, Antonis Rokas, and John A. Capra. 2021. “The Influence of Evolutionary History on Human Health and Disease.” Nature Reviews. Genetics 22 (5): 269–83. 10.1038/s41576-020-00305-9.

Bernabeu, Elena, Oriol Canela-Xandri, Konrad Rawlik, Andrea Talenti, James Prendergast, and Albert Tenesa. 2021. “Sex Differences in Genetic Architecture in the UK Biobank.” Nature Genetics 53 (9): 1283–89. 10.1038/s41588-021-00912-0.

Bulik-Sullivan, Brendan, Hilary K. Finucane, Verneri Anttila, Alexander Gusev, Felix R. Day, Po-Ru Loh, Laramie Duncan, et al. 2015. “An Atlas of Genetic Correlations across Human Diseases and Traits.” Nature Genetics 47 (11): 1236–41. 10.1038/ng.3406.

Burgess, Stephen, Deborah J Thompson, Jessica M B Rees, Felix R Day, John R Perry, and Ken K Ong. 2017. “Dissecting Causal Pathways Using Mendelian Randomization with Summarized Genetic Data: Application to Age at Menarche and Risk of Breast Cancer.” Genetics 207 (2): 481–87. 10.1534/genetics.117.300191.

Burgess, Stephen, Olena Yavorska, James Staley, Fernando Hartwig, Jim Broadbent, Christopher Foley, Andrew Grant, et al. 2024. “MendelianRandomization: Mendelian Randomization Package.” https://cran.r-project.org/web/packages/MendelianRandomization/index.html.

Cerezo, Maria, Elliot Sollis, Yue Ji, Elizabeth Lewis, Ala Abid, Karatuğ Ozan Bircan, Peggy Hall, et al. 2025. “The NHGRI-EBI GWAS Catalog: Standards for Reusability, Sustainability and Diversity.” Nucleic Acids Research 53 (D1): D998–1005. 10.1093/nar/gkae1070.

Charlesworth, Brian. 1994. Evolution in Age-Structured Populations. Cambridge University Press.

Crespi, Bernard J. 2010. “The Origins and Evolution of Genetic Disease Risk in Modern Humans.” Annals of the New York Academy of Sciences 1206 (1): 80–109. 10.1111/j.1749-6632.2010.05707.x.

Deelen, Joris, Daniel S. Evans, Dan E. Arking, Niccolò Tesi, Marianne Nygaard, Xiaomin Liu, Mary K. Wojczynski, et al. 2019. “A Meta-Analysis of Genome-Wide Association Studies Identifies Multiple Longevity Genes.” Nature Communications 10 (1): 3669. 10.1038/s41467-019-11558-2.

Dillin, Andrew, Douglas K. Crawford, and Cynthia Kenyon. 2002. “Timing Requirements for Insulin/IGF-1 Signaling in C. Elegans.” *Science (New York*, N.Y.) 298 (5594): 830–34. 10.1126/science.1074240.

Field, Yair, Evan A Boyle, Natalie Telis, Ziyue Gao, Kyle J. Gaulton, David Golan, Loic Yengo, et al. 2016. “Detection of Human Adaptation during the Past 2000 Years.” Science 354 (6313): 760–64. 10.1126/science.aag0776.

Frejka, Tomas. 2016. “The Demographic Transition Revisited: A Cohort Perspective.” WP-2016-012. 0 ed. Rostock: Max Planck Institute for Demographic Research. 10.4054/MPIDR-WP-2016-012.

Gagnon, Alain, Ken R. Smith, Marc Tremblay, Hélène Vézina, Paul-Philippe Paré, and Bertrand Desjardins. 2009. “Is There a Trade-off between Fertility and Longevity? A Comparative Study of Women from Three Large Historical Databases Accounting for Mortality Selection.” American Journal of Human Biology 21 (4): 533–40. 10.1002/ajhb.20893.

Gao, Ziyue. 2024. “Unveiling Recent and Ongoing Adaptive Selection in Human Populations.” PLOS Biology 22 (1): e3002469. 10.1371/journal.pbio.3002469.

Gardner, Eugene J., Matthew D. C. Neville, Kaitlin E. Samocha, Kieron Barclay, Martin Kolk, Mari E. K. Niemi, George Kirov, Hilary C. Martin, and Matthew E. Hurles. 2022. “Reduced Reproductive Success Is Associated with Selective Constraint on Human Genes.” Nature 603 (7903): 858–63. 10.1038/s41586-022-04549-9.

Ge, Yu-Zheng, Lu-Wei Xu, Rui-Peng Jia, Zheng Xu, Wen-Cheng Li, Ran Wu, Sheng Liao, et al. 2014. “Association of Polymorphisms in Estrogen Receptors (ESR1 and ESR2) with Male Infertility: A Meta-Analysis and Systematic Review.” Journal of Assisted Reproduction and Genetics 31 (5): 601–11. 10.1007/s10815-014-0212-5.

Giaimo, Stefano, and Arne Traulsen. 2019. “Generation Time Measures the Trade-Off between Survival and Reproduction in a Life Cycle.” The American Naturalist 194 (2): 285–90. 10.1086/704155.

Harshman, Lawrence G., and Anthony J. Zera. 2007. “The Cost of Reproduction: The Devil in the Details.” Trends in Ecology & Evolution 22 (2): 80–86. 10.1016/j.tree.2006.10.008.

Hsu, Chen-Hao, Oliver Posegga, Kai Fischbach, and Henriette Engelhardt. 2021. “Examining the Trade-Offs between Human Fertility and Longevity over Three Centuries Using Crowdsourced Genealogy Data.” PLOS ONE 16 (8): e0255528. 10.1371/journal.pone.0255528.

Hu, Dingxue, Yan Li, Detao Zhang, Jiahong Ding, Zijun Song, Junxia Min, Yi Zeng, and Chao Nie. 2022. “Genetic Trade-Offs between Complex Diseases and Longevity.” Aging Cell 21 (7): e13654. 10.1111/acel.13654.

Jensen, Martin Blomberg, Lærke Priskorn, Tina Kold Jensen, Anders Juul, and Niels Erik Skakkebaek. 2015. “Temporal Trends in Fertility Rates: A Nationwide Registry Based Study from 1901 to 2014.” PLOS ONE 10 (12): e0143722. 10.1371/journal.pone.0143722.

Jia, Gengjie, Yu Li, Hanxin Zhang, Ishanu Chattopadhyay, Anders Boeck Jensen, David R. Blair, Lea Davis, et al. 2019. “Estimating Heritability and Genetic Correlations from Large Health Datasets in the Absence of Genetic Data.” Nature Communications 10 (1): 5508. 10.1038/s41467-019-13455-0.

Joshi, Peter K., Nicola Pirastu, Katherine A. Kentistou, Krista Fischer, Edith Hofer, Katharina E. Schraut, David W. Clark, et al. 2017. “Genome-Wide Meta-Analysis Associates HLA-DQA1/DRB1 and LPA and Lifestyle Factors with Human Longevity.” Nature Communications 8 (1): 910. 10.1038/s41467-017-00934-5.

Kaptijn, Ralf, Fleur Thomese, Aart C. Liefbroer, Frans Van Poppel, David Van Bodegom, and Rudi G. J. Westendorp. 2015. “The Trade-Off between Female Fertility and Longevity during the Epidemiological Transition in the Netherlands.” PLOS ONE 10 (12): e0144353. 10.1371/journal.pone.0144353.

Kirkwood, Thomas B. L., and Steven N. Austad. 2000. “Why Do We Age?” Nature 408 (6809): 233–38. 10.1038/35041682.

Kuningas, Maris, Signe Altmäe, André G. Uitterlinden, Albert Hofman, Cornelia M. van Duijn, and Henning Tiemeier. 2011. “The Relationship between Fertility and Lifespan in Humans.” Age 33 (4): 615–22. 10.1007/s11357-010-9202-4.

Leeuw, Christiaan A. de, Joris M. Mooij, Tom Heskes, and Danielle Posthuma. 2015. “MAGMA: Generalized Gene-Set Analysis of GWAS Data.” PLOS Computational Biology 11 (4): e1004219. 10.1371/journal.pcbi.1004219.

Liu, Aoxing, Evelina T. Akimova, Xuejie Ding, Sakari Jukarainen, Pekka Vartiainen, Tuomo Kiiskinen, Sara Koskelainen, et al. 2024. “Evidence from Finland and Sweden on the Relationship between Early-Life Diseases and Lifetime Childlessness in Men and Women.” Nature Human Behaviour 8 (2): 276–87. 10.1038/s41562-023-01763-x.

Long, Erping, and Jianzhi Zhang. 2023. “Evidence for the Role of Selection for Reproductively Advantageous Alleles in Human Aging.” Science Advances 9 (49): eadh4990. 10.1126/sciadv.adh4990.

Lycett, J.e, R.i.m Dunbar, and E Voland. 2000. “Longevity and the Costs of Reproduction in a Historical Human Population.” Proceedings of the Royal Society of London. Series B: Biological Sciences 267 (1438): 31–35. 10.1098/rspb.2000.0962.

Mathieson, Iain, Felix R. Day, Nicola Barban, Felix C. Tropf, David M. Brazel, Diana van Heemst, Ahmad Vaez, et al. 2023. “Genome-Wide Analysis Identifies Genetic Effects on Reproductive Success and Ongoing Natural Selection at the FADS Locus.” Nature Human Behaviour 7 (5): 790–801. 10.1038/s41562-023-01528-6.

Medawar, P.B. 1952. *An Unsolved Problem of Biology: An Inaugural Lecture Delivered at University College, London, 6 December*, 1951. London: H.K. Lewis and Company.

Melzer, David, Luke C. Pilling, and Luigi Ferrucci. 2020. “The Genetics of Human Ageing.” Nature Reviews Genetics 21 (2): 88–101. 10.1038/s41576-019-0183-6.

Morita, Masahito. 2018. “Demographic Studies Enhance the Understanding of Evolutionarily (Mal)Adaptive Behaviors and Phenomena in Humans: A Review on Fertility Decline and an Integrated Model.” Population Ecology 60 (1): 143–54. 10.1007/s10144-017-0597-y.

Nesse, Randolph M., and George C. Williams. 1996. *Why We Get Sick: The New Science of Darwinian Medicine*. 1st ed. New York: Vintage Books. https://www.penguinrandomhouse.com/books/120768/why-we-get-sick-by-randolph-m-nesse/.

Nettle, Daniel, and Helen Clegg. 2005. “Schizotypy, Creativity and Mating Success in Humans.” Proceedings of the Royal Society B: Biological Sciences 273 (1586): 611–15. 10.1098/rspb.2005.3349.

Paskulin, Diego Davila, João Sabino Cunha-Filho, Livia Davila Paskulin, Carlos Augusto Bastos Souza, and Patricia Ashton-Prolla. 2013. “ESR1 Rs9340799 Is Associated with Endometriosis-Related Infertility and In Vitro Fertilization Failure.” Disease Markers 35 (6): 796290. 10.1155/2013/796290.

Pettay, Jenni E., Loeske E. B. Kruuk, Jukka Jokela, and Virpi Lummaa. 2005. “Heritability and Genetic Constraints of Life-History Trait Evolution in Preindustrial Humans.” Proceedings of the National Academy of Sciences of the United States of America 102 (8): 2838–43. 10.1073/pnas.0406709102.

Rodríguez, Juan Antonio, Urko M. Marigorta, David A. Hughes, Nino Spataro, Elena Bosch, and Arcadi Navarro. 2017. “Antagonistic Pleiotropy and Mutation Accumulation Influence Human Senescence and Disease.” Nature Ecology & Evolution 1 (3): 1–5. 10.1038/s41559-016-0055.

Smith, J. Maynard. 1958. “The Effects of Temperature and of Egg-Laying on the Longevity of Drosophila Subobscura.” Journal of Experimental Biology 35 (4): 832–42. 10.1242/jeb.35.4.832.

Stearns, S. C. 1989. “Trade-Offs in Life-History Evolution.” Functional Ecology 3 (3): 259–68. 10.2307/2389364.

Sudlow, Cathie, John Gallacher, Naomi Allen, Valerie Beral, Paul Burton, John Danesh, Paul Downey, et al. 2015. “UK Biobank: An Open Access Resource for Identifying the Causes of a Wide Range of Complex Diseases of Middle and Old Age.” PLOS Medicine 12 (3): e1001779. 10.1371/journal.pmed.1001779.

Sullivan, Patrick F. 2010. “The Psychiatric GWAS Consortium: Big Science Comes to Psychiatry.” Neuron 68 (2): 182–86. 10.1016/j.neuron.2010.10.003.

Tabatabaie, Vafa, Gil Atzmon, Swapnil N. Rajpathak, Ruth Freeman, Nir Barzilai, and Jill Crandall. 2011. “Exceptional Longevity Is Associated with Decreased Reproduction.” Aging 3 (12): 1202–5. 10.18632/aging.100415.

Timmers, Paul RHJ, Ninon Mounier, Kristi Lall, Krista Fischer, Zheng Ning, Xiao Feng, Andrew D Bretherick, et al. 2019. “Genomics of 1 Million Parent Lifespans Implicates Novel Pathways and Common Diseases and Distinguishes Survival Chances.” eLife 8 (January):e39856. 10.7554/eLife.39856.

Tropf, Felix C., Gert Stulp, Nicola Barban, Peter M. Visscher, Jian Yang, Harold Snieder, and Melinda C. Mills. 2015. “Human Fertility, Molecular Genetics, and Natural Selection in Modern Societies.” PLOS ONE 10 (6): e0126821. 10.1371/journal.pone.0126821.

Verbanck, Marie, Chia-Yen Chen, Benjamin Neale, and Ron Do. 2018. “Detection of Widespread Horizontal Pleiotropy in Causal Relationships Inferred from Mendelian Randomization between Complex Traits and Diseases.” Nature Genetics 50 (5): 693–98. 10.1038/s41588-018-0099-7.

Viippola, Essi, Sara Kuitunen, Rodosthenis S Rodosthenous, Andrius Vabalas, Tuomo Hartonen, Pekka Vartiainen, Joanne Demmler, et al. 2023. “Data Resource Profile: Nationwide Registry Data for High-Throughput Epidemiology and Machine Learning (FinRegistry).” International Journal of Epidemiology 52 (4): e195–200. 10.1093/ije/dyad091.

Voight, Benjamin F., Sridhar Kudaravalli, Xiaoquan Wen, and Jonathan K. Pritchard. 2006. “A Map of Recent Positive Selection in the Human Genome.” PLOS Biology 4 (3): e72. 10.1371/journal.pbio.0040072.

Watanabe, Kyoko, Sven Stringer, Oleksandr Frei, Maša Umićević Mirkov, Christiaan de Leeuw, Tinca J. C. Polderman, Sophie van der Sluis, Ole A. Andreassen, Benjamin M. Neale, and Danielle Posthuma. 2019. “A Global Overview of Pleiotropy and Genetic Architecture in Complex Traits.” Nature Genetics 51 (9): 1339–48. 10.1038/s41588-019-0481-0.

Watanabe, Kyoko, Erdogan Taskesen, Arjen van Bochoven, and Danielle Posthuma. 2017. “Functional Mapping and Annotation of Genetic Associations with FUMA.” Nature Communications 8 (1): 1826. 10.1038/s41467-017-01261-5.

Welter, Danielle, Jacqueline MacArthur, Joannella Morales, Tony Burdett, Peggy Hall, Heather Junkins, Alan Klemm, et al. 2014. “The NHGRI GWAS Catalog, a Curated Resource of SNP-Trait Associations.” Nucleic Acids Research 42 (D1): D1001–6. 10.1093/nar/gkt1229.

Westendorp, Rudi G. J., and Thomas B. L. Kirkwood. 1998. “Human Longevity at the Cost of Reproductive Success.” Nature 396 (6713): 743–46. 10.1038/25519.

Willer, Cristen J., Yun Li, and Gonçalo R. Abecasis. 2010. “METAL: Fast and Efficient Meta-Analysis of Genomewide Association Scans.” Bioinformatics 26 (17): 2190–91. 10.1093/bioinformatics/btq340.

Williams, GC. 1957. “Pleiotropy, Natural Selection, and the Evolution of Senescence.” Evolution (NY) 11:398–411. 10.2307/2406060.

World Health Assembly, 43. 1990. “Report of the International Conference for the Tenth Revision of the International Classification of Diseases.” https://iris.who.int/handle/10665/173188.

Wu, Dou, Zi Wang, Jingying Huang, Liang Huang, Songbo Zhang, Ruixue Zhao, Wei Li, Di Chen, and Guangshuo Ou. 2022. “An Antagonistic Pleiotropic Gene Regulates the Reproduction and Longevity Tradeoff.” Proceedings of the National Academy of Sciences of the United States of America 119 (18): e2120311119. 10.1073/pnas.2120311119.

Ye, Chao-Jie, Li-Jie Kong, Yi-Ying Wang, Chun Dou, Jie Zheng, Min Xu, Yu Xu, et al. 2023. “Mendelian Randomization Evidence for the Causal Effects of Socio-Economic Inequality on Human Longevity among Europeans.” Nature Human Behaviour 7 (8): 1357–70. 10.1038/s41562-023-01646-1.

