## Supplementary Figures and Note for "Why do we get sick? Genetic evidence for evolutionary trade-offs between fertility, longevity, and disease"

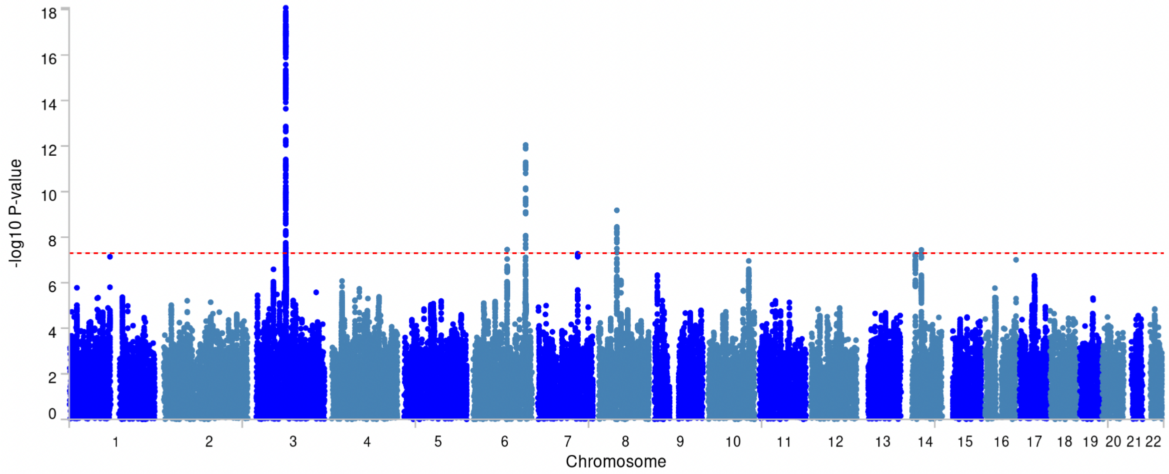

**Supplementary Figure 1. Manhattan plot of GWAS results from the fertility meta-analysis.** This Manhattan plot generated by FUMA illustrates the genome-wide association results for meta-analysis on related measures of offspring number. The x-axis represents genomic positions across chromosomes 1 to 22, ordered sequentially. The y-axis displays the *$-log_{10}$(p-value)* for each single nucleotide polymorphism (SNP). Each point represents an individual SNP, with its position corresponding to its chromosomal location and its height indicating the strength of association with the fertility phenotype. The red horizontal line denotes the genome-wide significance threshold ($p < 5\times10^{-8}$). SNPs above this line are considered statistically significant associations. Different colors alternate between chromosomes to aid visualization.

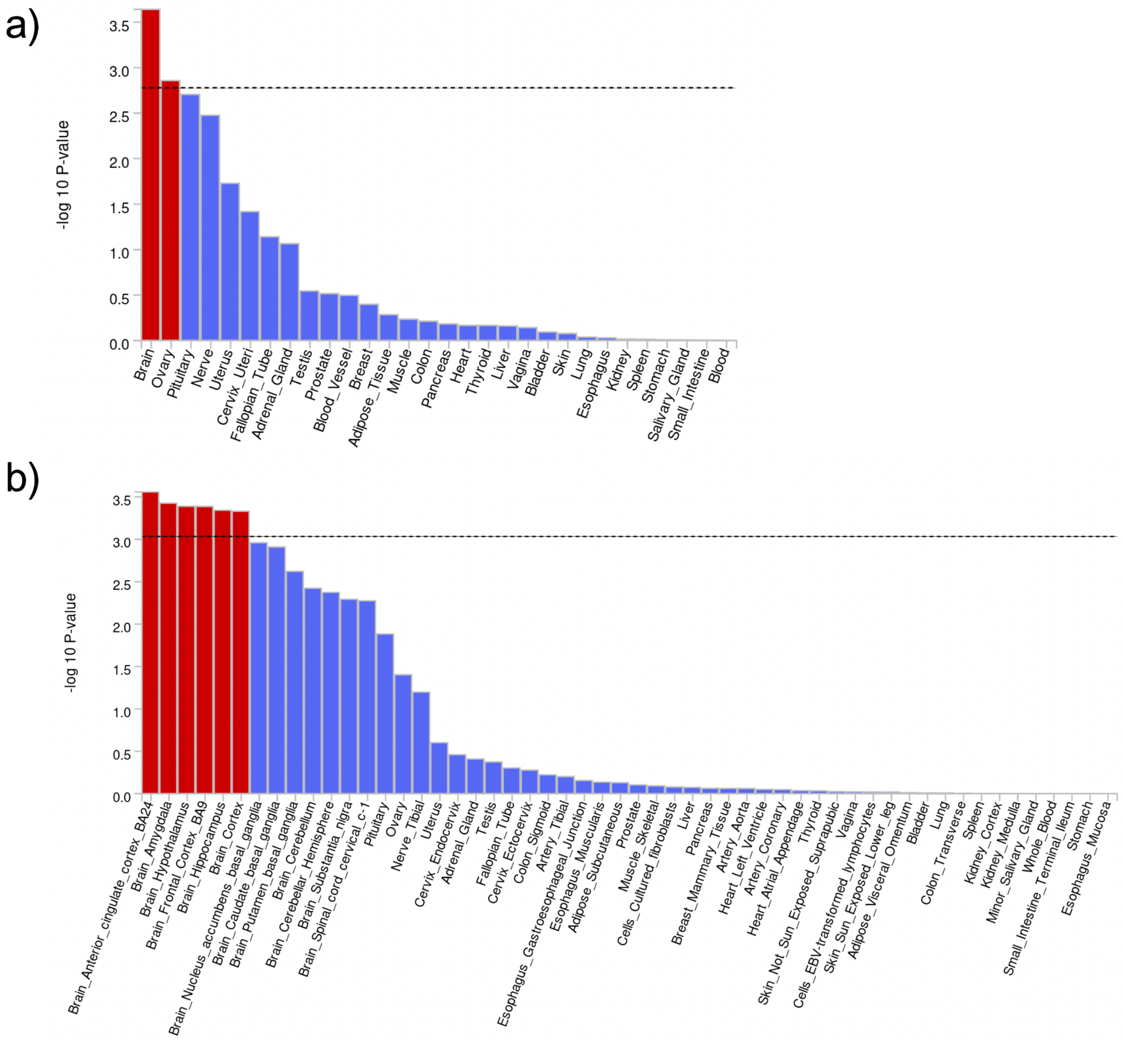

**Supplementary Figure 2. MAGMA tissue expression analysis for the fertility meta-analysis GWAS.** MAGMA gene-property analysis results for gene expression across GTEx v8 are shown for (a) 30 general tissue types and (b) 53 specific tissue types, as implemented in FUMA SNP2GENE. The x-axis represents tissue types, and the y-axis displays the $-log_{10}(p\text{-}value)$ for the association between tissue-specific gene expression and genetic associations with fertility. The black dashed line denotes the Bonferroni-corrected significance threshold. Bars exceeding this threshold indicate tissues with significant enrichment of fertility-associated genes. This analysis identifies tissues where fertility-associated genes are preferentially expressed, providing insights into relevant biological pathways.

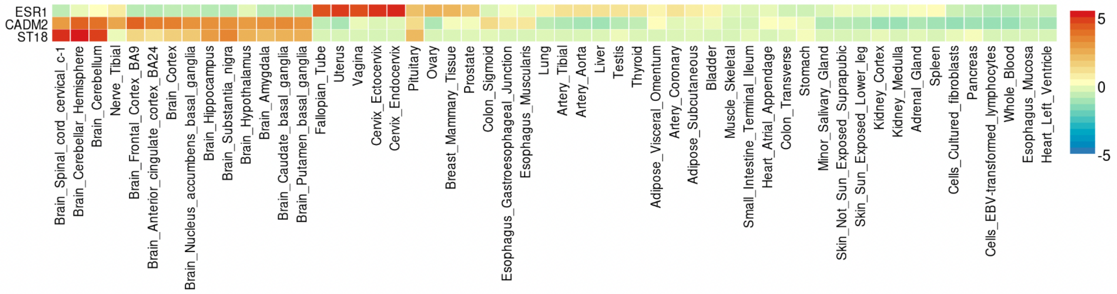

**Supplementary Figure 3**. **Gene expression heatmap for prioritized genes from the fertility meta-analysis GWAS**. Gene expression heatmap using GTEx v8 54 specific tissue types for the prioritized genes from the fertility meta-analysis, as implemented in the FUMA GENE2FUNC tool. The x-axis displays tissues ordered by clusters from general tissue types. The y-axis shows the prioritized genes. Color intensity represents the normalized expression values per tissue in log2 transformation, enabling comparison between tissues for each gene. Red colors indicate higher expression, blue colors indicate lower expression, and neutral colors indicate average expression levels.

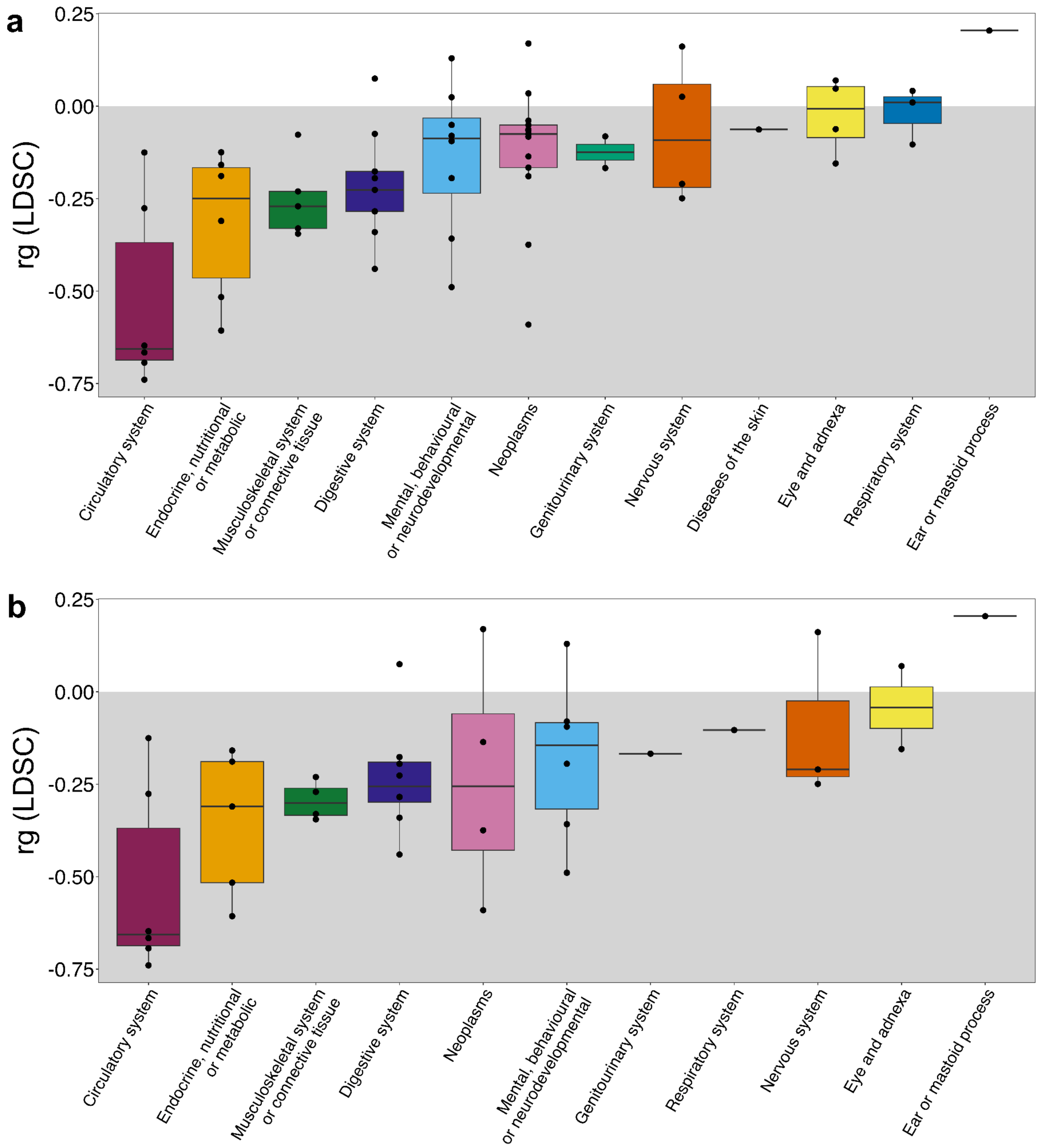

**Supplementary Figure 4. Genetic correlations between longevity and complex diseases across ICD10 domains.** (a) Comprehensive Analysis: Box plots representing genetic correlations for all 62 diseases studied, grouped by their respective ICD10 domains. (b) Significant Associations: Box plots depicting genetic correlations for the subset of 41 diseases that demonstrated statistically significant associations with longevity ($P < 0.05$). Each box plot displays the median, interquartile range, and individual diseases within each ICD-10 domain, allowing for comparison of the strength and direction of genetic associations across different disease domains. The gray shaded area highlights the negative genetic correlations.

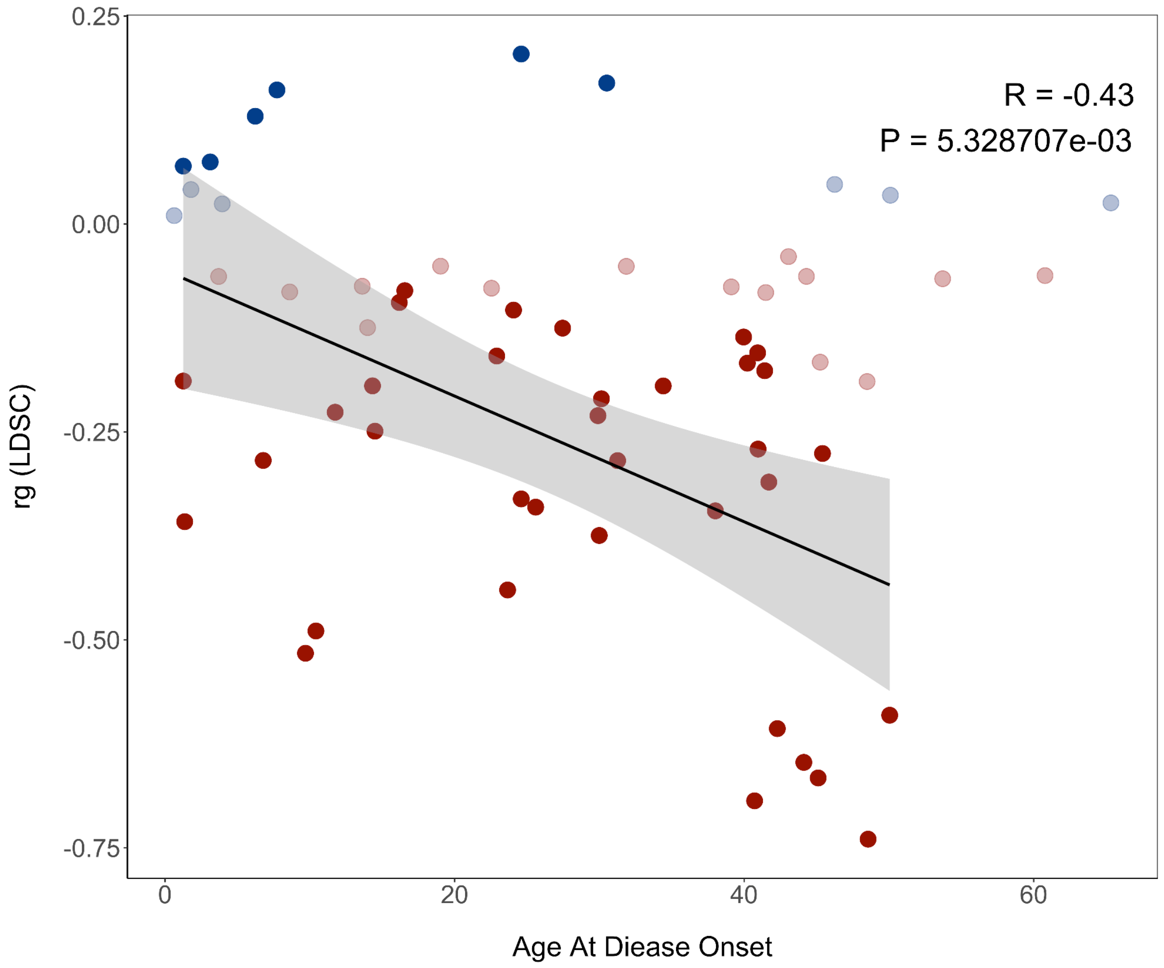

**Supplementary Figure 5**. **Global genetic correlations between longevity and complex diseases: influence of age at disease onset (P < 0.05)**. Spearman correlation plot illustrating the global genetic correlation between longevity and complex diseases, stratified by the age at disease onset. Blue dots represent positive genetic correlations, while red dots denote negative genetic correlations. Strongly colored dots indicate diseases with significant correlation p-value (P < 0.05). The correlation analysis includes only significant diseases, revealing a negative relationship between genetic correlation with longevity and age at disease onset (r=–0.43, P=5.3e-03).

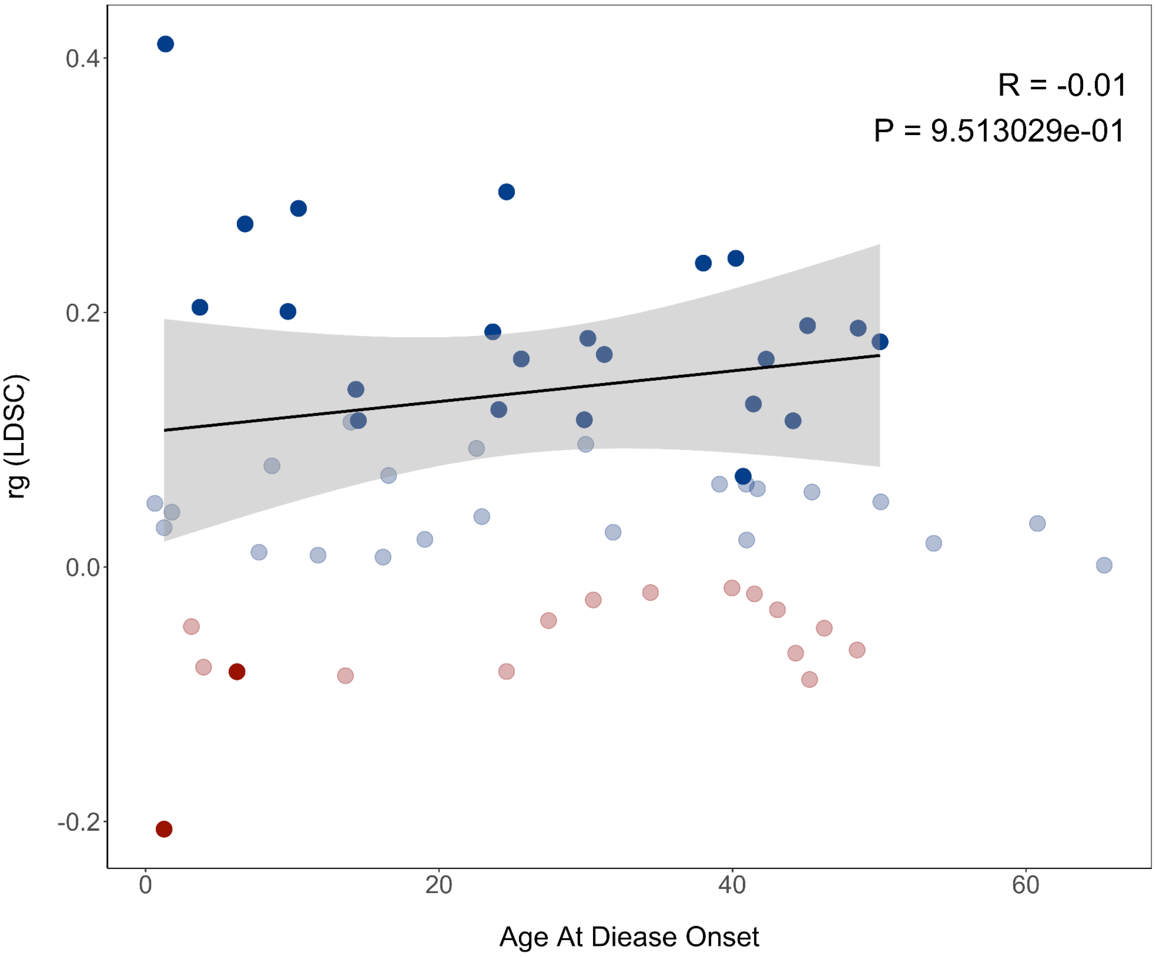

**Supplementary Figure 6**. **Global genetic correlations between fertility and complex diseases: influence of age at disease onset (P < 0.05)**. Spearman correlation plot illustrating the global genetic correlation between fertility and complex diseases, stratified by the age at disease onset. Blue dots represent positive genetic correlations, while red dots denote negative genetic correlations. Strongly colored dots indicate diseases with significant correlation p-value (P < 0.05). The correlation analysis includes only significant diseases, revealing no relationship between genetic correlation with fertility and age at disease onset (r=-0.01, P=9.5-e01).

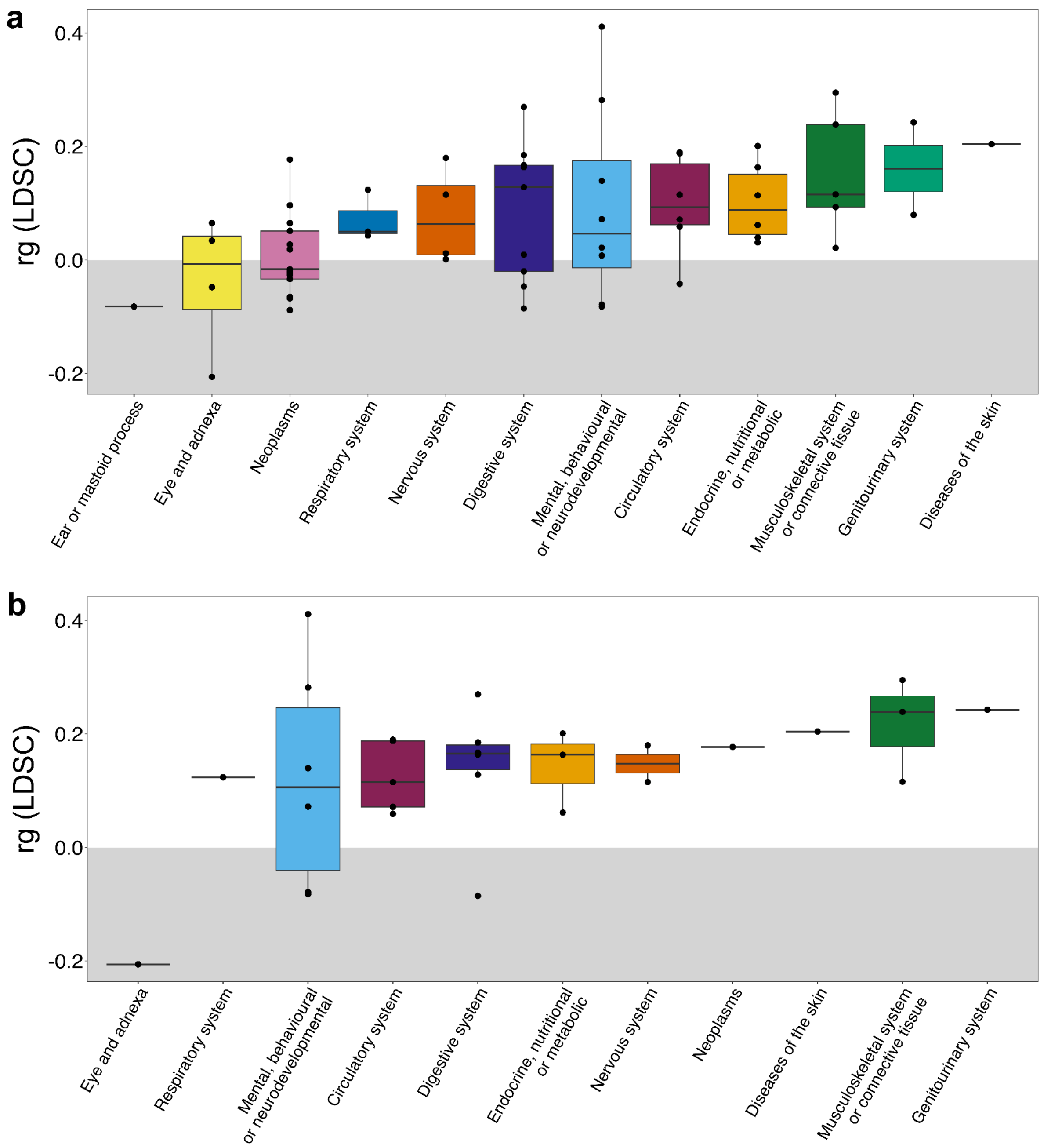

**Supplementary Figure 7. Genetic correlations between fertility and complex diseases across ICD10 domains.** (a) Comprehensive Analysis: Box plots representing genetic correlations for all 62 diseases studied, grouped by their respective ICD10 domains. (b) Significant Associations: Box plots depicting genetic correlations for the subset of 38 diseases that demonstrated statistically significant associations with longevity after applying multiple-test correction (FDR < 0.05). Each box plot displays the median, interquartile range, and individual diseases within each ICD-10 domain, allowing for comparison of the strength and direction of genetic associations across different disease domains. The gray shaded area highlights the negative genetic correlations.

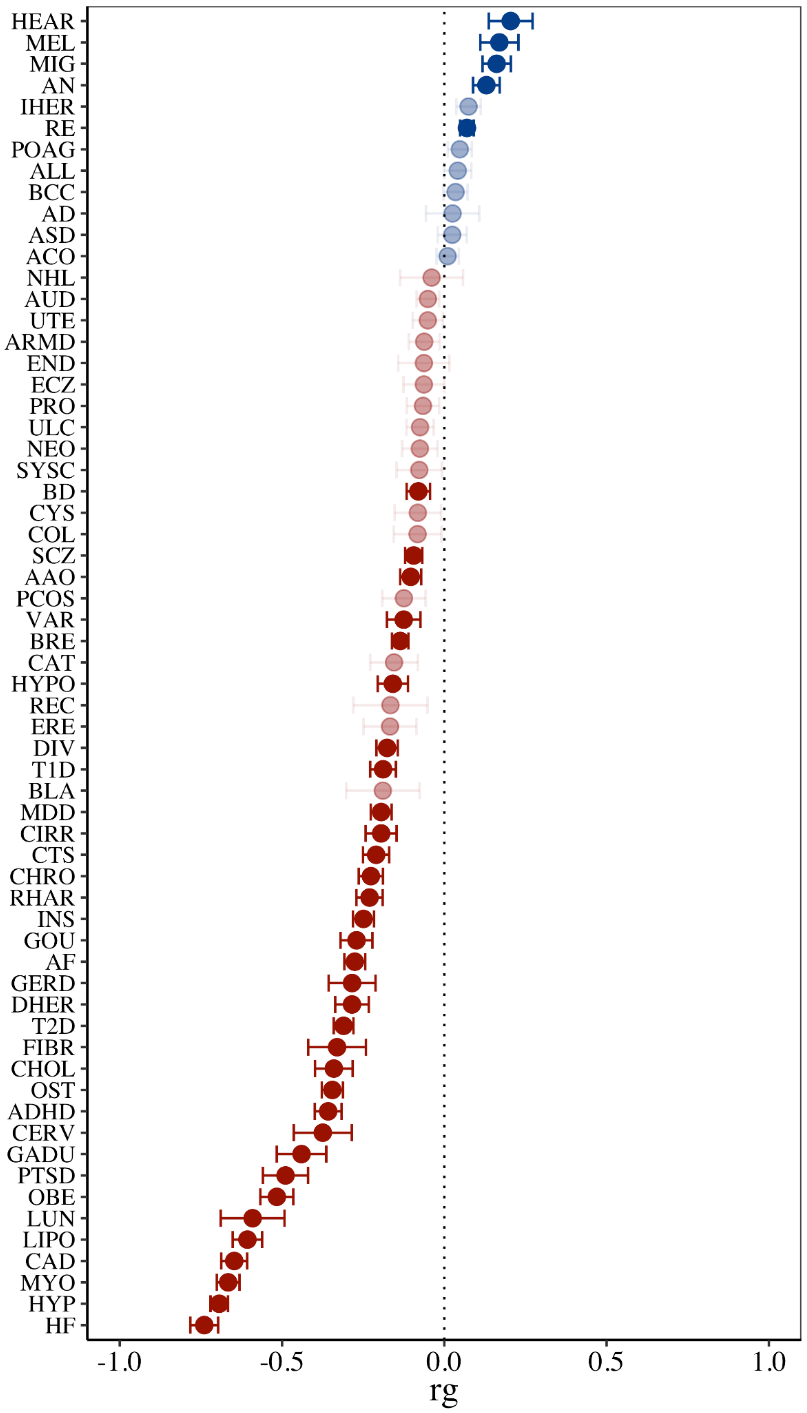

**Supplementary Figure 8. Global genetic correlations between longevity and complex diseases (FDR<0.05)**. Global genetic correlations, computed with LDSC, between longevity and 62 complex diseases. While Figure 1 of the main text used p-values ($P < 0.05$) to determine significance, here we confirm that the trends remain consistent when applying multiple-test correction using the Benjamini-Hochberg false discovery rate. In the figure, red indicates negative correlations and blue indicates positive correlations with longevity. Increased transparency denotes non-significant correlations (FDR>0.05), whereas brighter colors signify significant correlations (FDR<0.05).

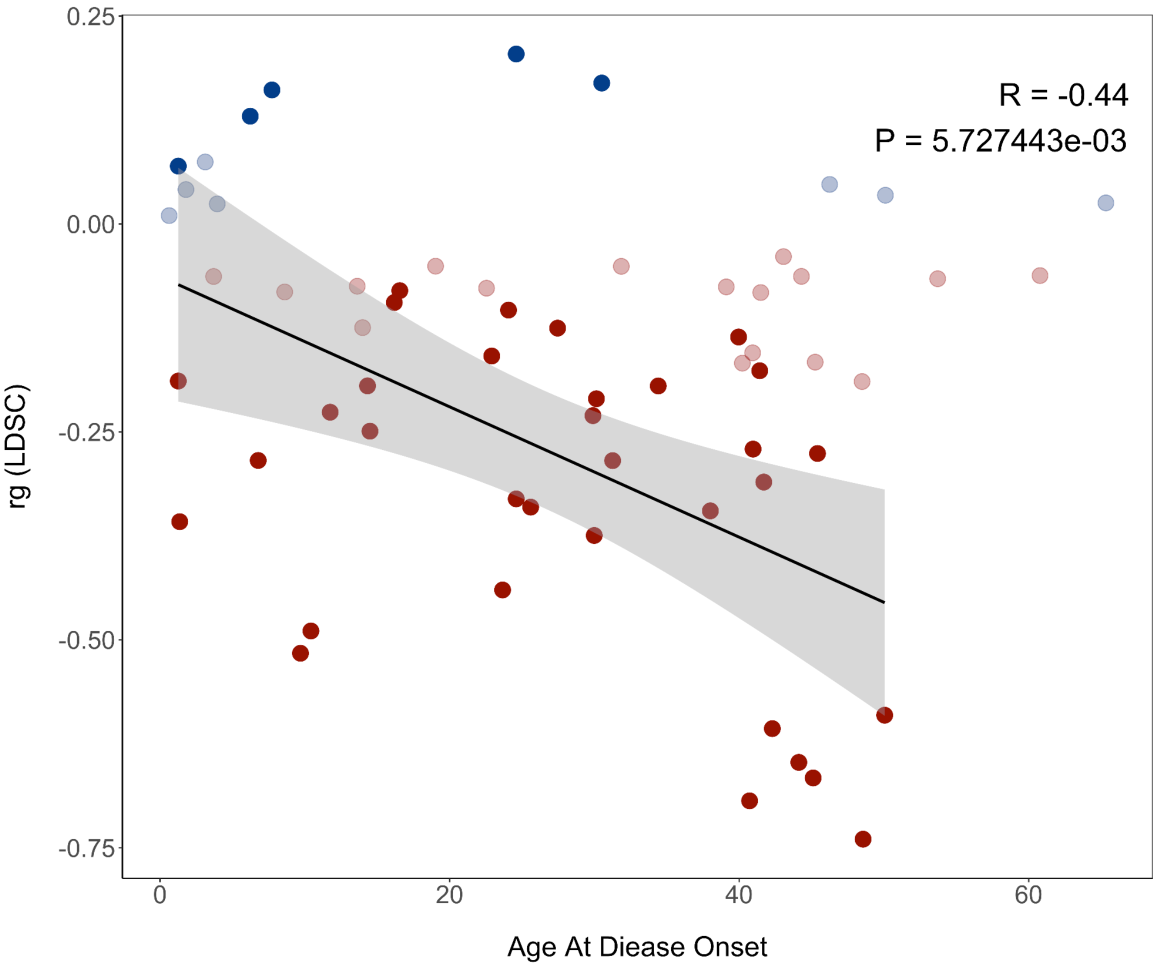

**Supplementary Figure 9. Global genetic correlations between longevity and complex diseases: influence of age at disease onset (FDR < 0.05)**. Correlation plot illustrating the global genetic correlation between longevity and complex diseases, stratified by the age at disease onset. Blue dots represent positive genetic correlations, while red dots denote negative genetic correlations. Strongly colored dots indicate diseases that remain significant after multiple testing corrections (FDR < 0.05). The correlation analysis includes only significant diseases, revealing a negative relationship between genetic correlation with longevity and age at disease onset (r = –0.44, p = 5.7e-03).

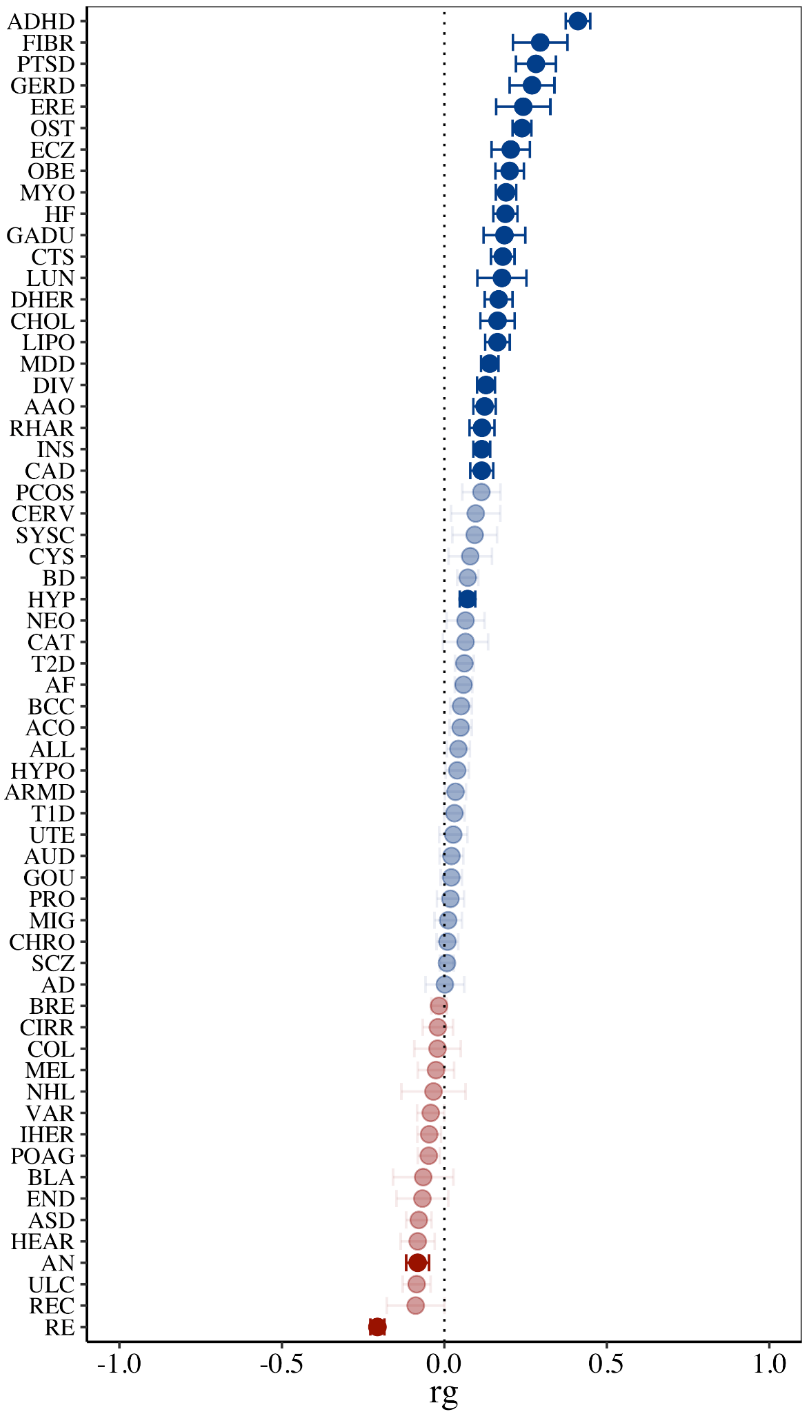

**Supplementary Figure 10. Global genetic correlations between fertility and complex diseases (FDR<0.05)**. Global genetic correlations, computed with LDSC, between fertility and 62 complex diseases. While Figure 1 of the main text used p-values (P < 0.05) to determine significance, here we confirm that the trends remain consistent when applying multiple-test correction using the Benjamini-Hochberg false discovery rate. In the figure, red indicates negative correlations and blue indicates positive correlations with fertility. Increased transparency denotes non-significant correlations (FDR>0.05), whereas brighter colors signify significant correlations (FDR<0.05).

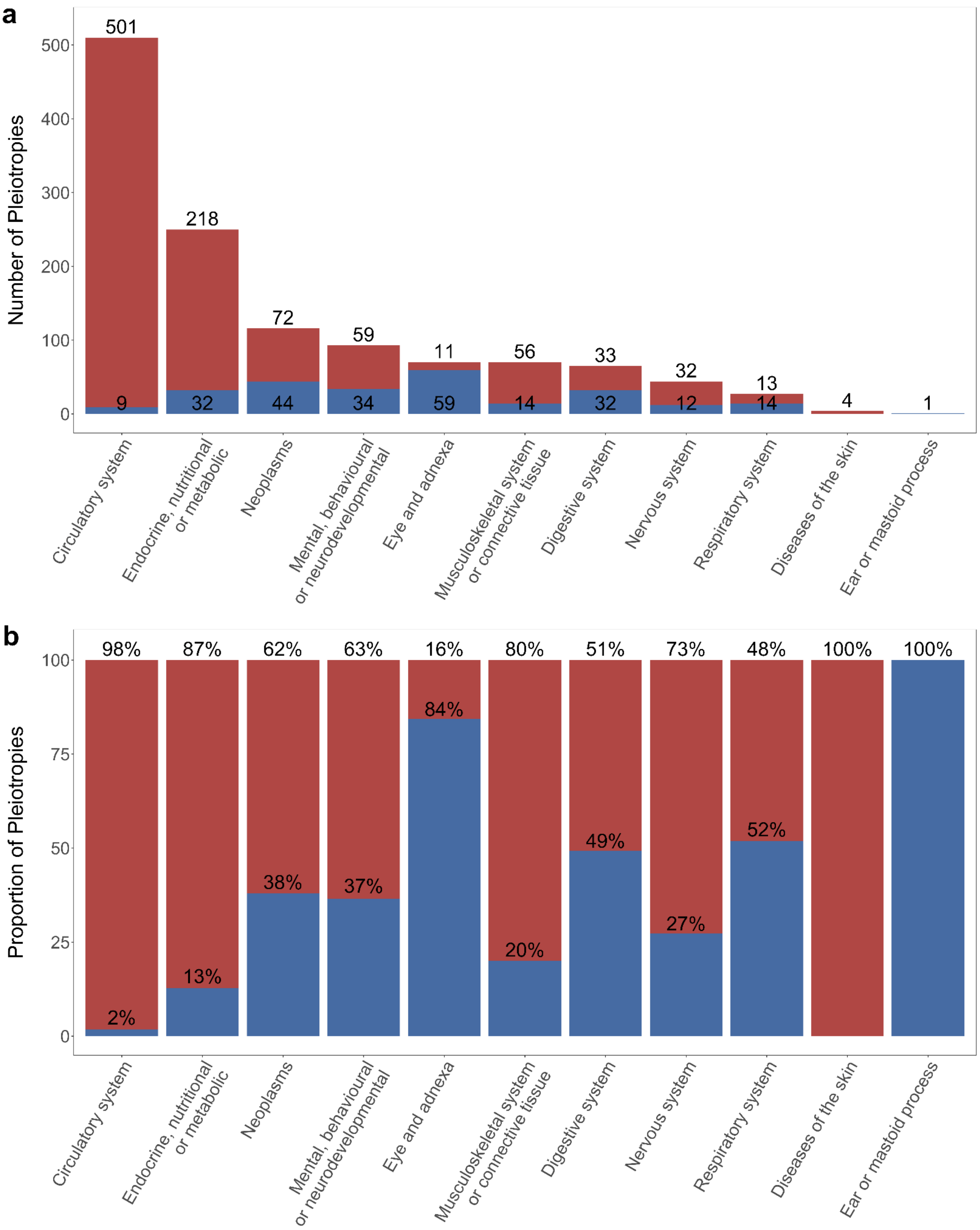

**Supplementary Figure 11**. **Distribution of longevity-associated pleiotropies across disease domains: counts and proportions.** Descriptive bar charts illustrating positive and negative pleiotropies with longevity by disease domain. Blue bars denote positive pleiotropies, whereas red bars indicate negative pleiotropies. a) The total counts of positive and negative pleiotropies within each domain. b) The relative proportions of positive and negative pleiotropies across domains.

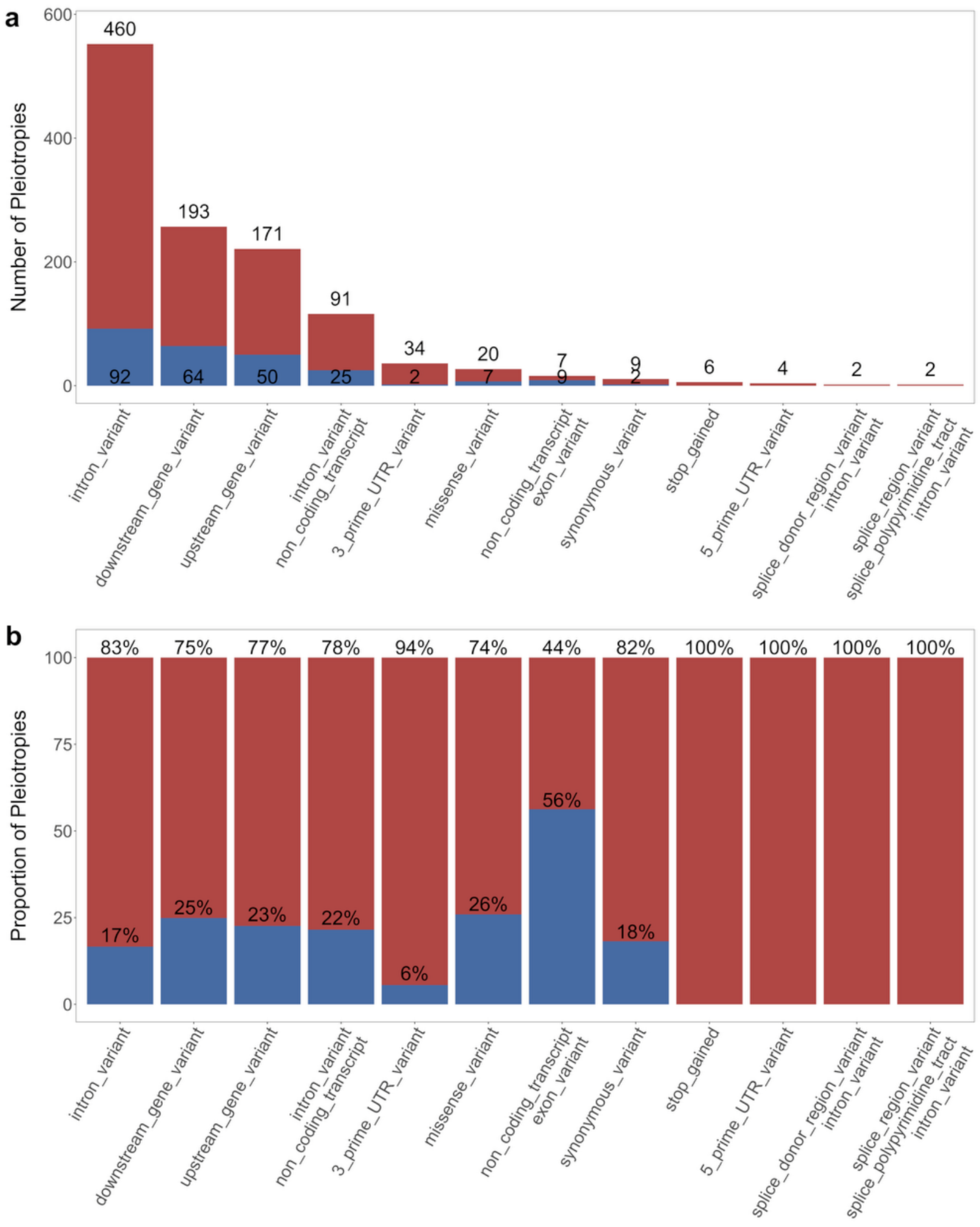

**Supplementary Figure 12**. **Distribution of longevity-associated pleiotropies across functional consequences: counts and proportions.** Descriptive bar charts illustrating positive and negative pleiotropies with longevity by functional consequence from Ensembl Variant Effect Predictor (VEP). Blue bars denote positive pleiotropies, whereas red bars indicate negative pleiotropies. a) The total counts of positive and negative pleiotropies within each consequence. b) The relative proportions of positive and negative pleiotropies across consequences.

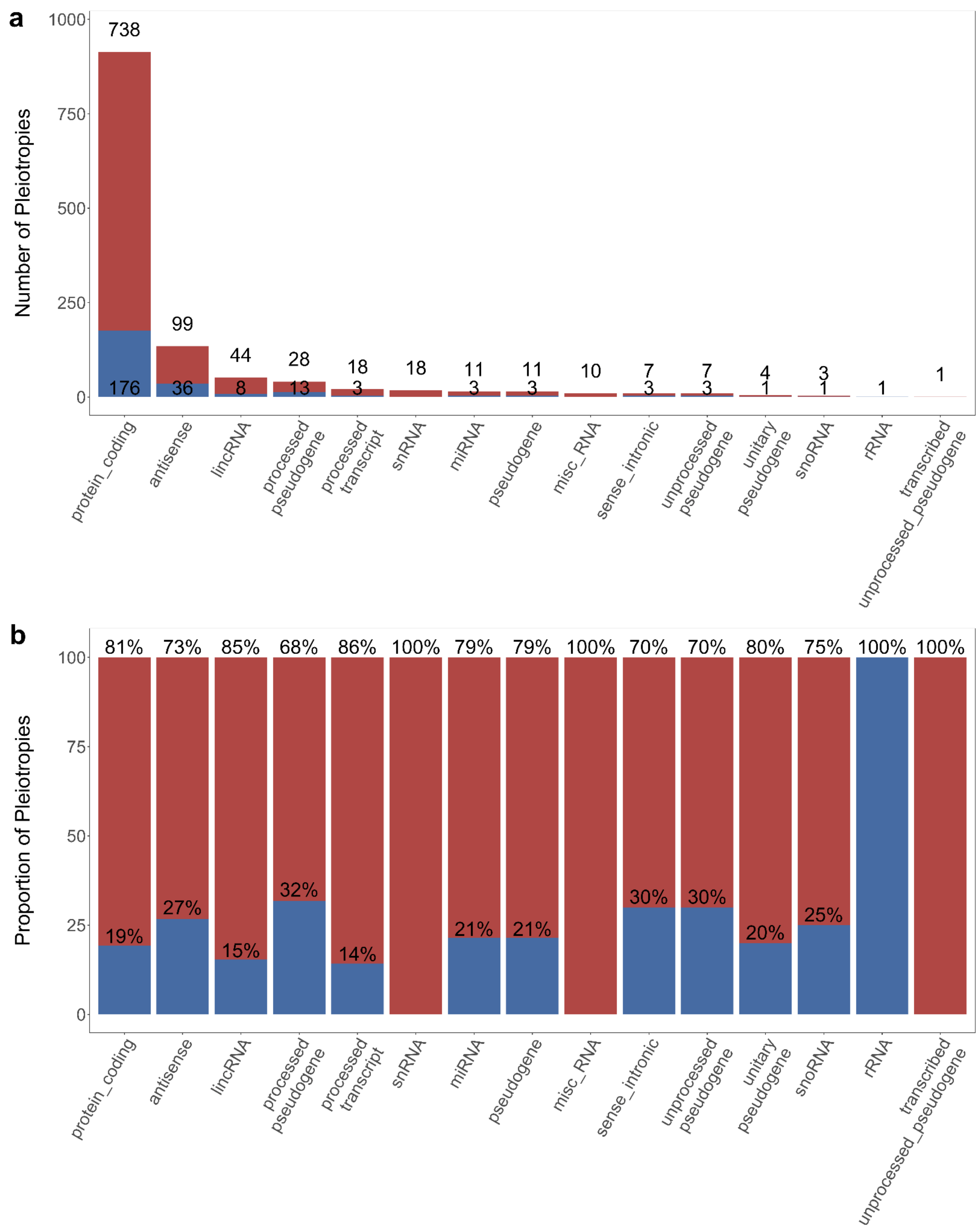

**Supplementary Figure 13**. **Distribution of longevity-associated pleiotropies across biotypes: counts and proportions.** Descriptive bar charts illustrating positive and negative pleiotropies with longevity by biotype from Ensembl Variant Effect Predictor (VEP). Blue bars denote positive pleiotropies, whereas red bars indicate negative pleiotropies. a) The total counts of positive and negative pleiotropies within each biotype. b) The relative proportions of positive and negative pleiotropies across biotypes.

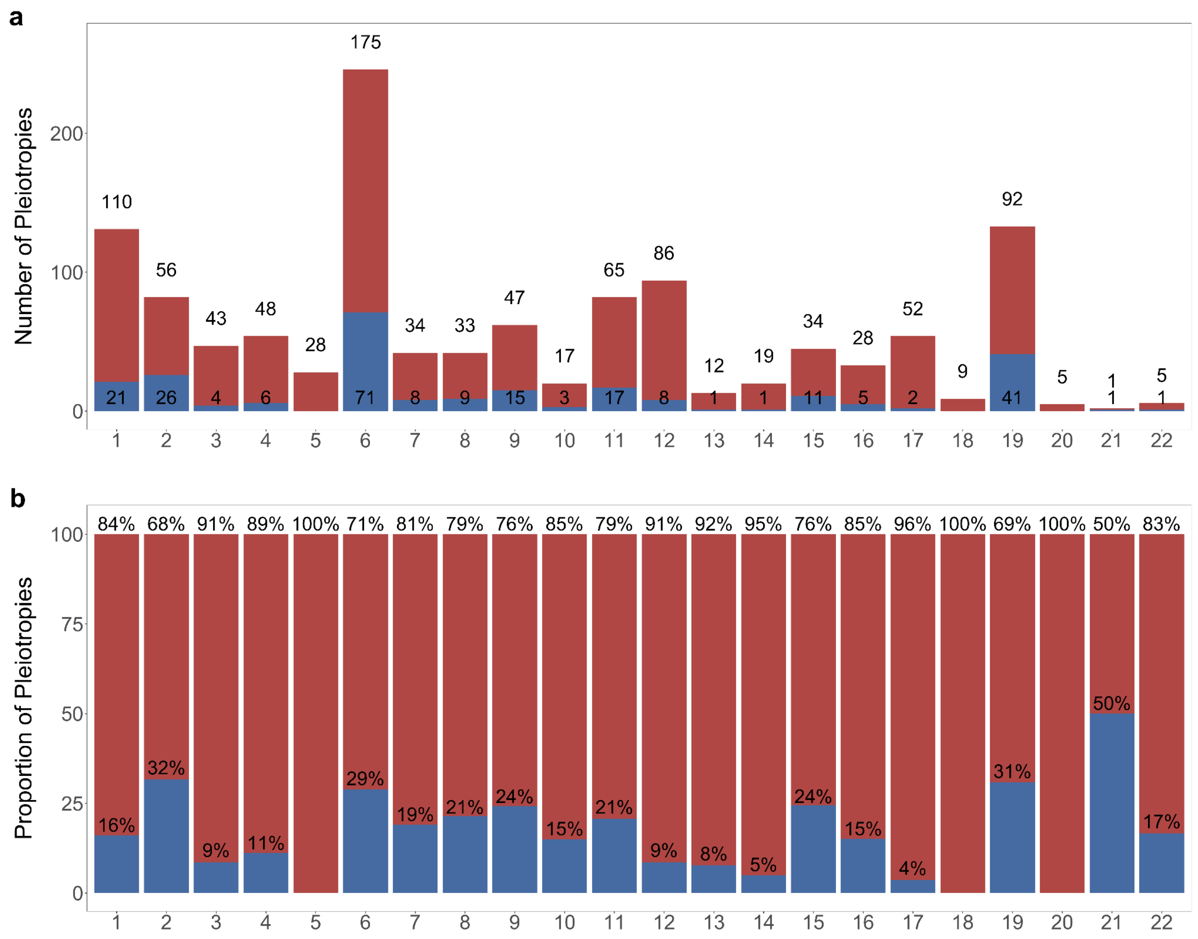

**Supplementary Figure 14**. **Chromosomal distribution of longevity-associated pleiotropies: counts and proportions.** Descriptive bar charts illustrating positive and negative pleiotropies with longevity by chromosome. Blue bars denote positive pleiotropies, whereas red bars indicate negative pleiotropies. a) The total counts of positive and negative pleiotropies within each chromosome. b) The relative proportions of positive and negative pleiotropies across chromosomes.

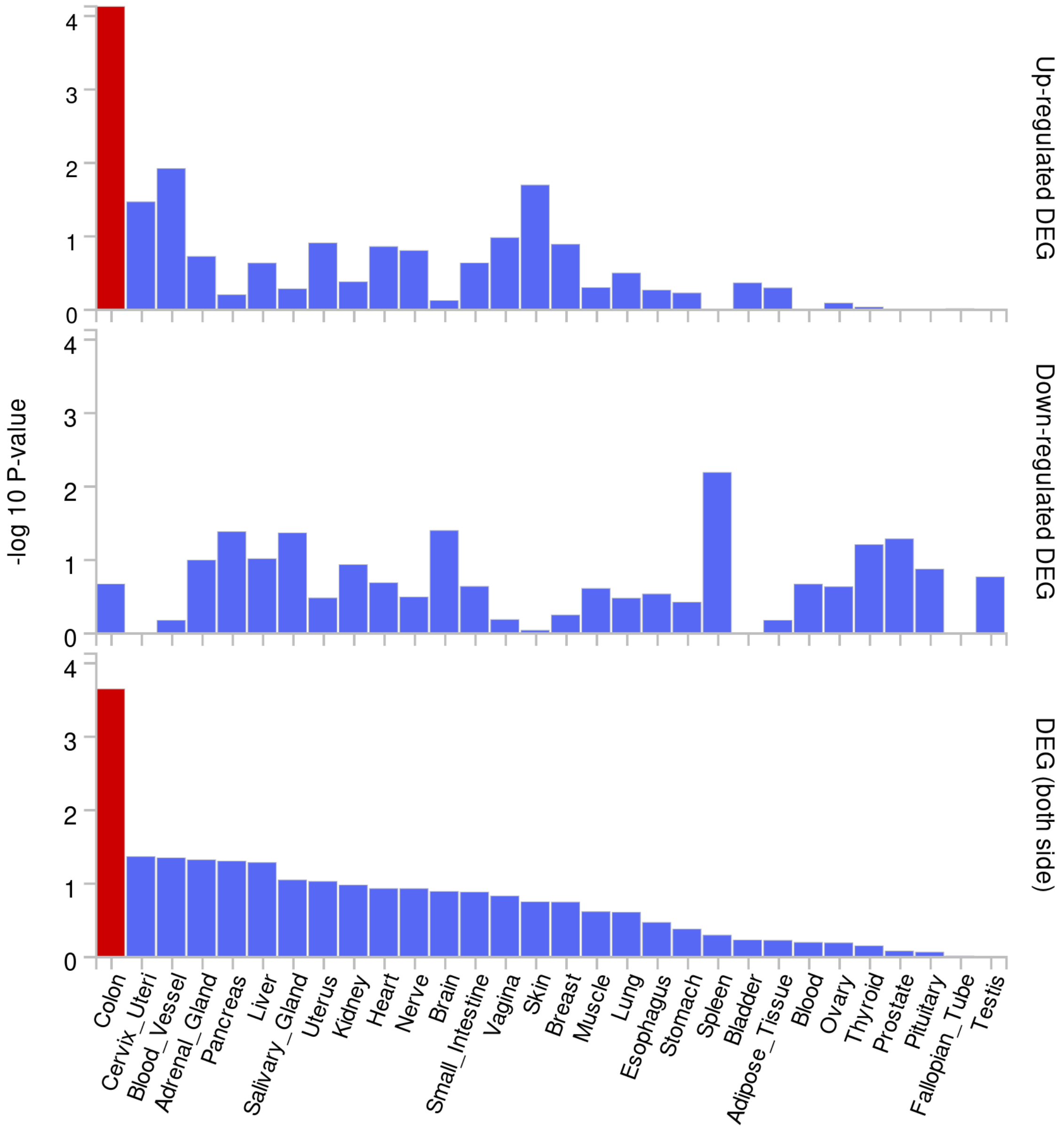

**Supplementary Figure 15. FUMA tissue expression analysis for the negative pleiotropies with longevity.** FUMA GENE2FUNC analysis results for differentially expressed genes (DEG) across GTEx v8 30 general tissue types. The x-axis represents tissue types, and the y-axis displays the $-\log_{10}$(p-value) for the DEG analysis. Bars exceeding the Bonferroni-corrected significance threshold ($P_{bon} < 0.05$) are highlighted in red and indicate tissues with differentially expressed genes. This analysis identifies tissues where negative pleiotropies between diseases and longevity are preferentially up- or down-regulated, providing insights into relevant biological pathways.

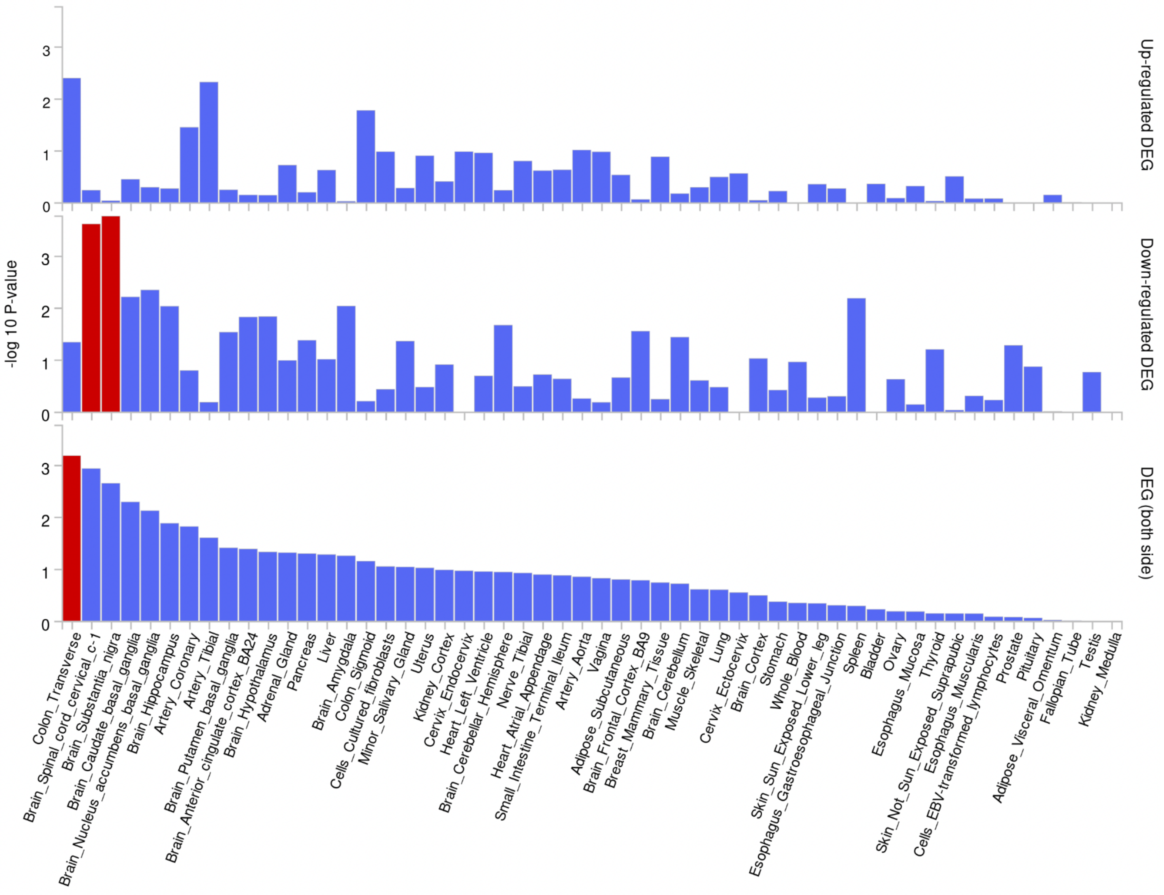

**Supplementary Figure 16. FUMA tissue expression analysis for the negative pleiotropies with longevity.** FUMA GENE2FUNC analysis results for differentially expressed genes (DEG) across GTEx v8 53 specific tissue types. The x-axis represents tissue types, and the y-axis displays the −log10(p-value) for the DEG analysis. Bars exceeding the Bonferroni-corrected significance threshold ($P_{bon} < 0.05$) are highlighted in red and indicate tissues with differentially expressed genes. This analysis identifies tissues where negative pleiotropies between diseases and longevity are preferentially up- or down-regulated, providing insights into relevant biological pathways.

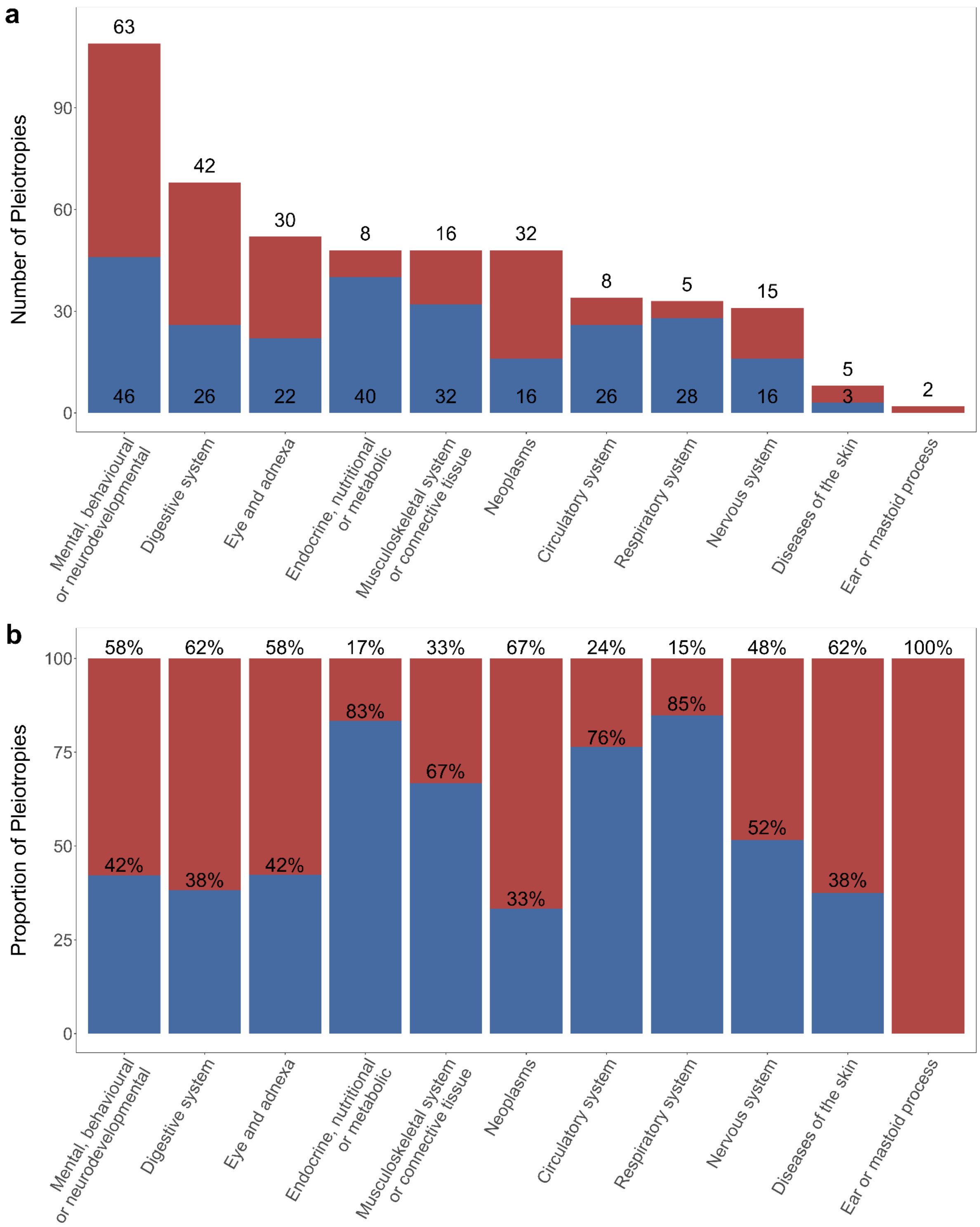

**Supplementary Figure 17**. **Distribution of fertility-associated pleiotropies across disease domains: counts and proportions.** Descriptive bar charts illustrating positive and negative pleiotropies with fertility by disease domain. Blue bars denote positive pleiotropies, whereas red bars indicate negative pleiotropies. a) The total counts of positive and negative pleiotropies within each domain. b) The relative proportions of positive and negative pleiotropies across domains.

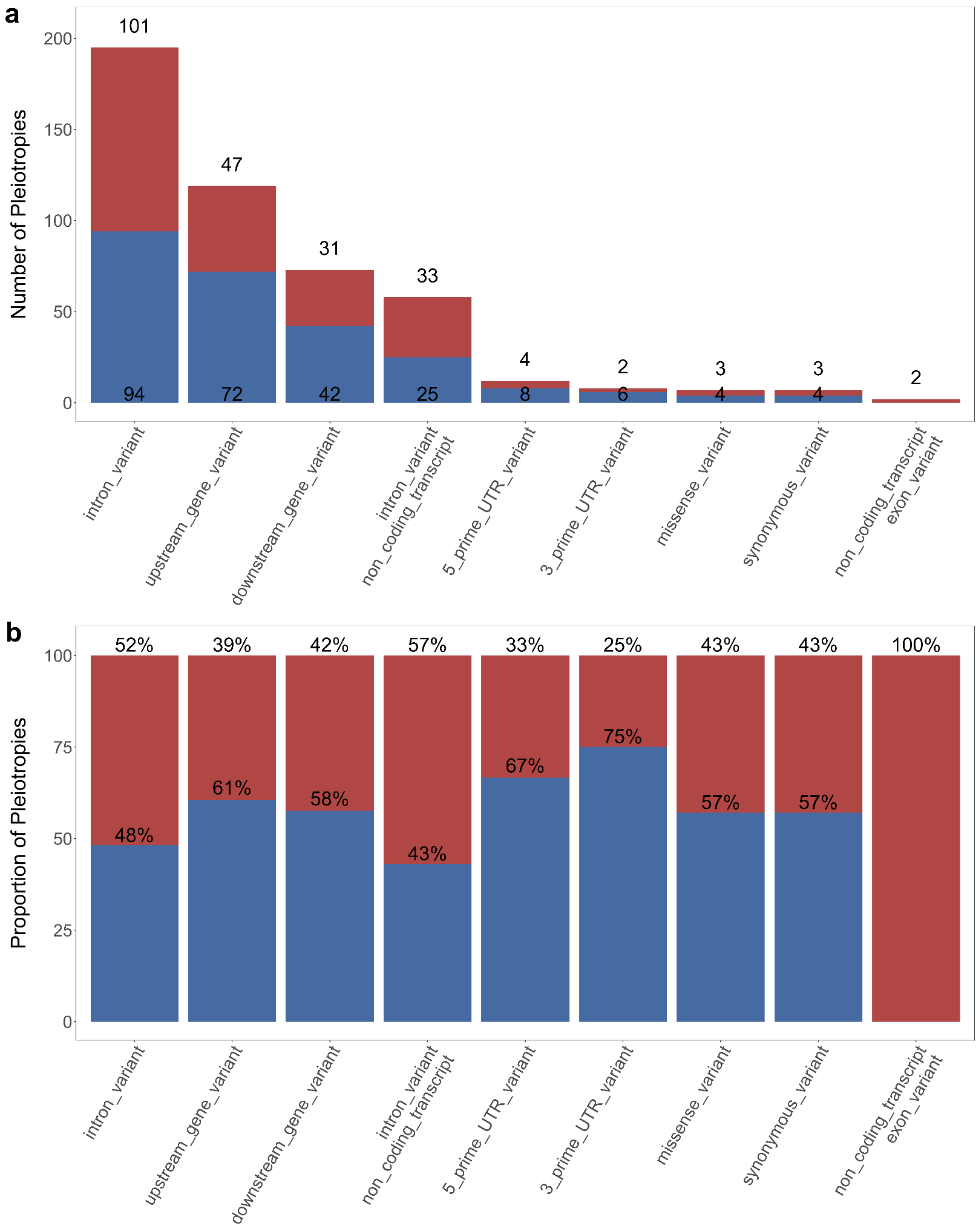

**Supplementary Figure 18**. **Distribution of fertility-associated pleiotropies across functional consequences: counts and proportions.** Descriptive bar charts illustrating positive and negative pleiotropies with fertility by functional consequence from Ensembl Variant Effect Predictor (VEP). Blue bars denote positive pleiotropies, whereas red bars indicate negative pleiotropies. a) The total counts of positive and negative pleiotropies within each consequence. b) The relative proportions of positive and negative pleiotropies across consequences.

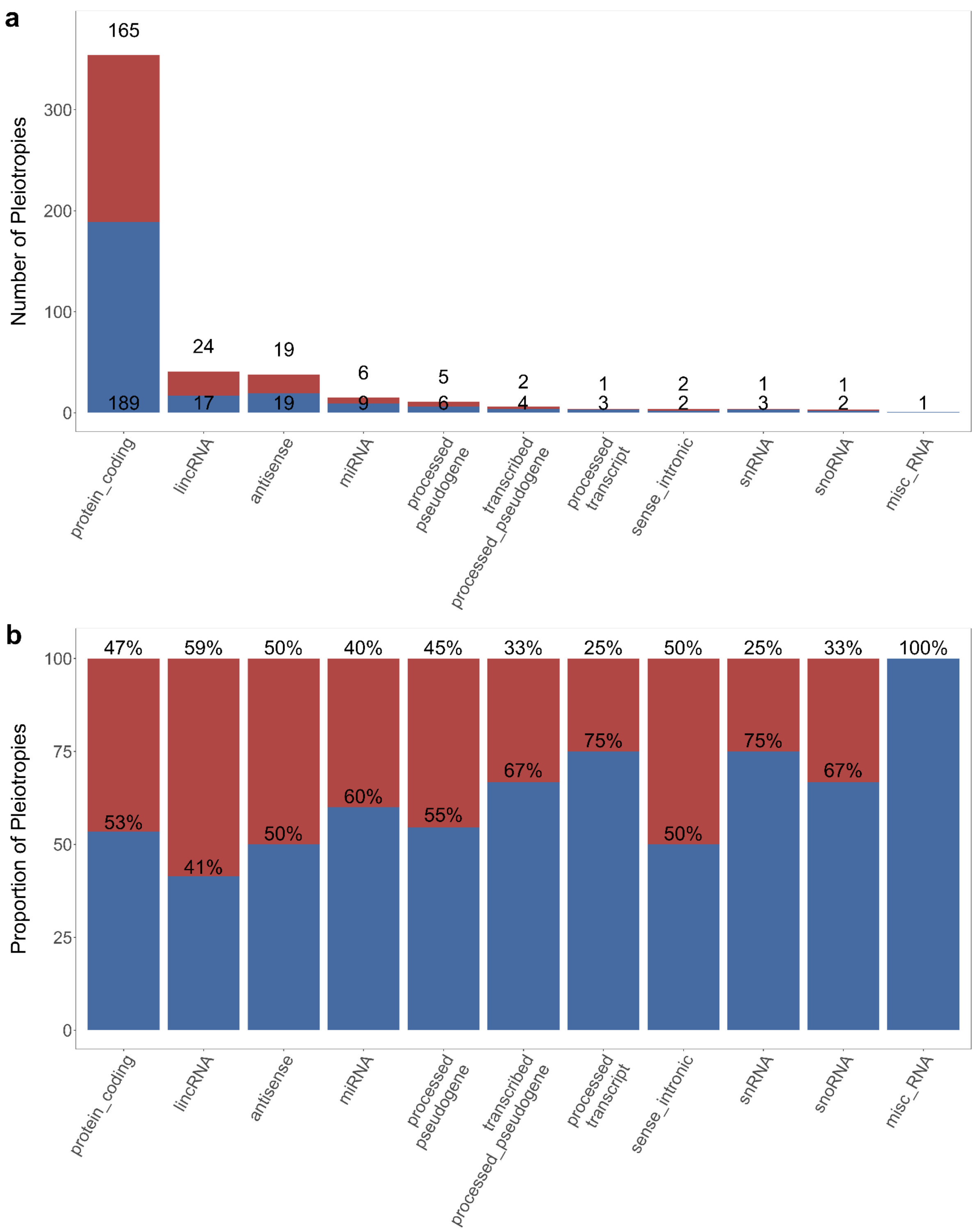

**Supplementary Figure 19**. **Distribution of fertility-associated pleiotropies across biotypes: counts and proportions.** Descriptive bar charts illustrating positive and negative pleiotropies with fertility by biotype from Ensembl Variant Effect Predictor (VEP). Blue bars denote positive pleiotropies, whereas red bars indicate negative pleiotropies. a) The total counts of positive and negative pleiotropies within each biotype. b) The relative proportions of positive and negative pleiotropies across biotypes.

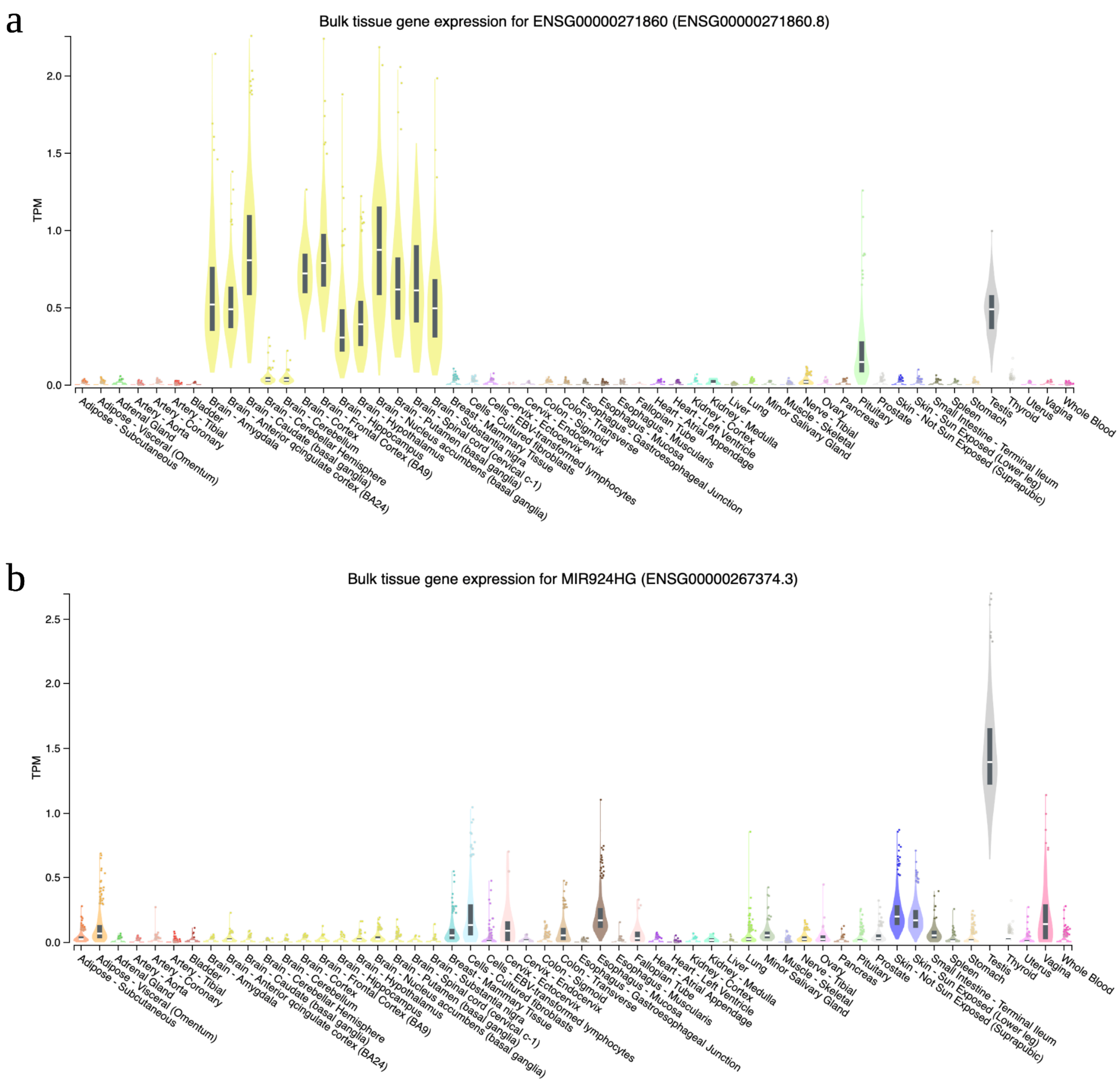

**Supplementary Figure 20**. **Tissue-specific expression of longevity- and fertility-associated lincRNAs.** Display of bulk tissue gene expression profiles (TPM) for 54 tissue types obtained from the GTEx Portal. Panels illustrate two long intergenic non-coding RNAs (lincRNAs) with pleiotropic associations to both longevity and fertility: panel (a) corresponds to ENSG000000271860.8 and panel (b) to ENSG000000267374.3.

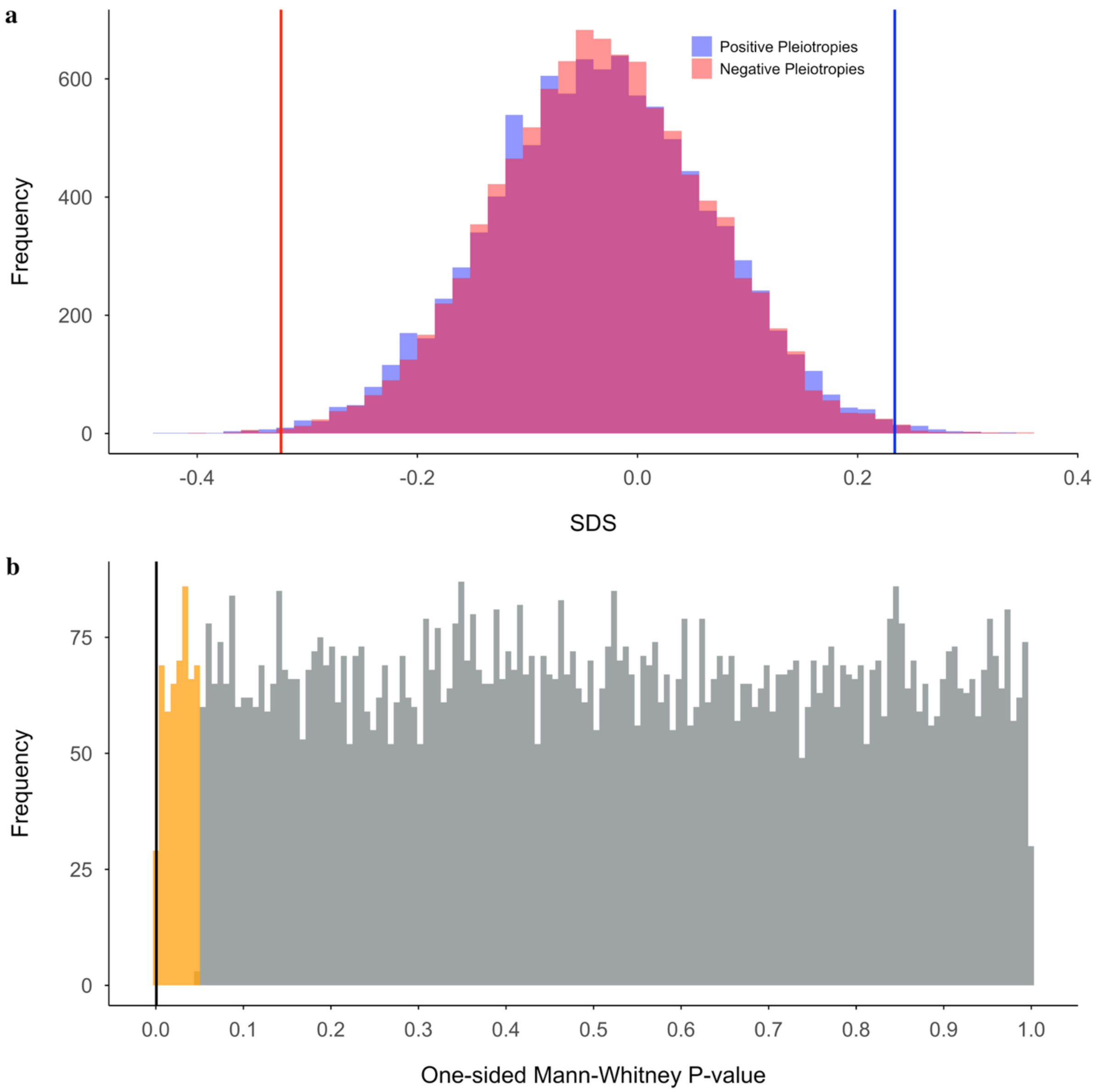

**Supplementary Figure 21. Resampling analysis of SDS scores for fertility-associated pleiotropies.** Panel (a) presents histograms of resampled Singleton Density Scores (SDS) for positive (blue) and negative (red) pleiotropy groups, with overlapping values shown in purple. The red vertical line marks the mean SDS for observed negative pleiotropies, and the blue line indicates the SDS for observed positive pleiotropies. Panel (b) displays the distribution of one-sided Mann-Whitney U-test p-values comparing the resampled means between the two groups. The yellow shaded area highlights p-values below 0.05, indicating statistical significance. The black vertical line represents the observed p-value, which falls within the most extreme 5% of the resampled distribution.

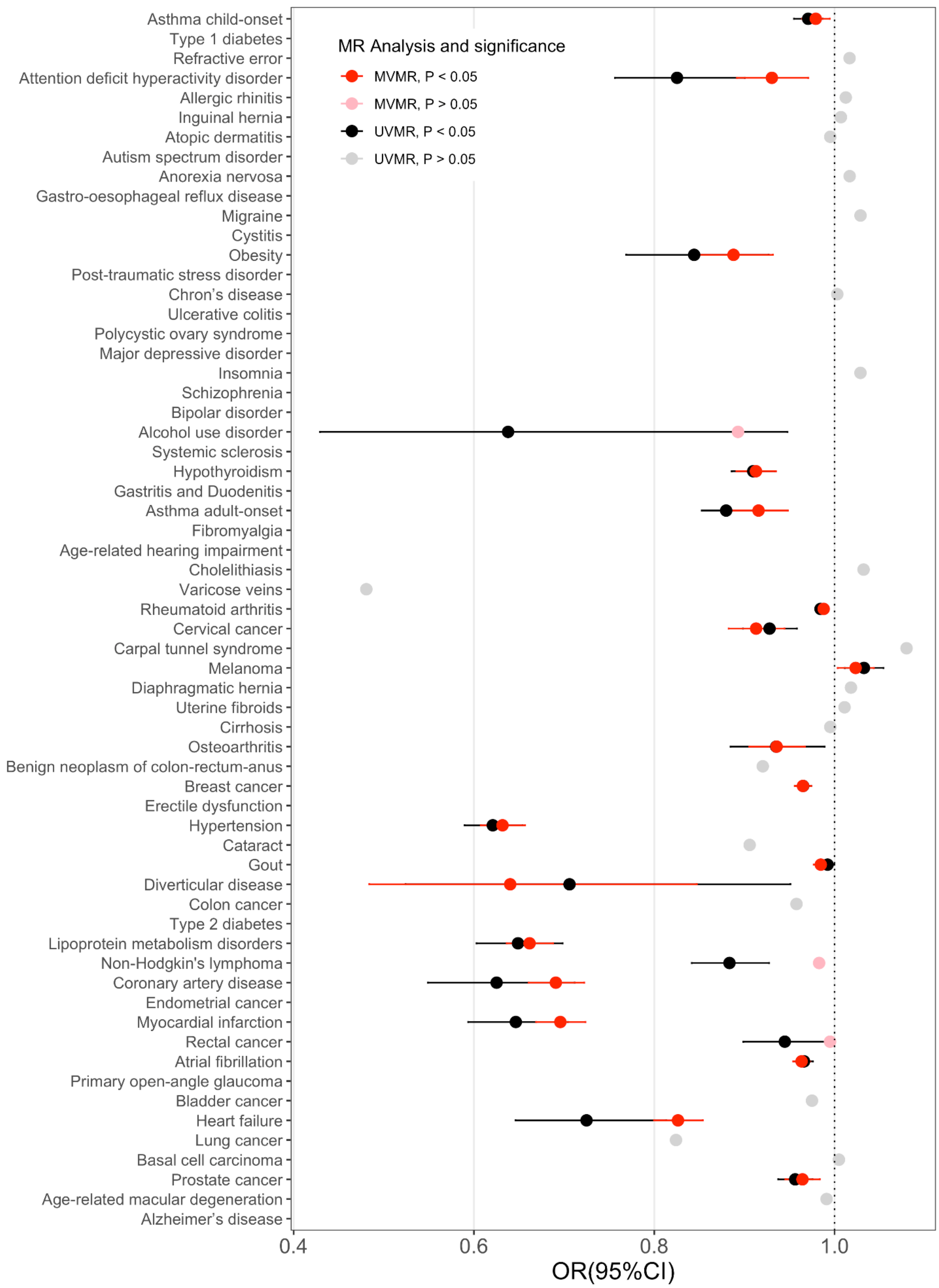

**Supplementary Figure 22. Mendelian Randomization analysis of the causal relationship between complex diseases and longevity.** The x-axis displays odds ratios of causal estimates from the MR analysis assessing the impact of complex diseases (exposures) on longevity (outcome). Two MR approaches were employed. In the univariable MR, significant estimates ($P < 0.05$) are shown in black with confidence intervals (CIs), while non-significant estimates are depicted in gray without CIs for clarity. In the multivariate MR, which incorporates education as a mediator, significant estimates ($P < 0.05$) are shown in red, and non-significant estimates in lighter pink, also without CIs. For some diseases the analysis could not be performed, then no information appears in this plot.

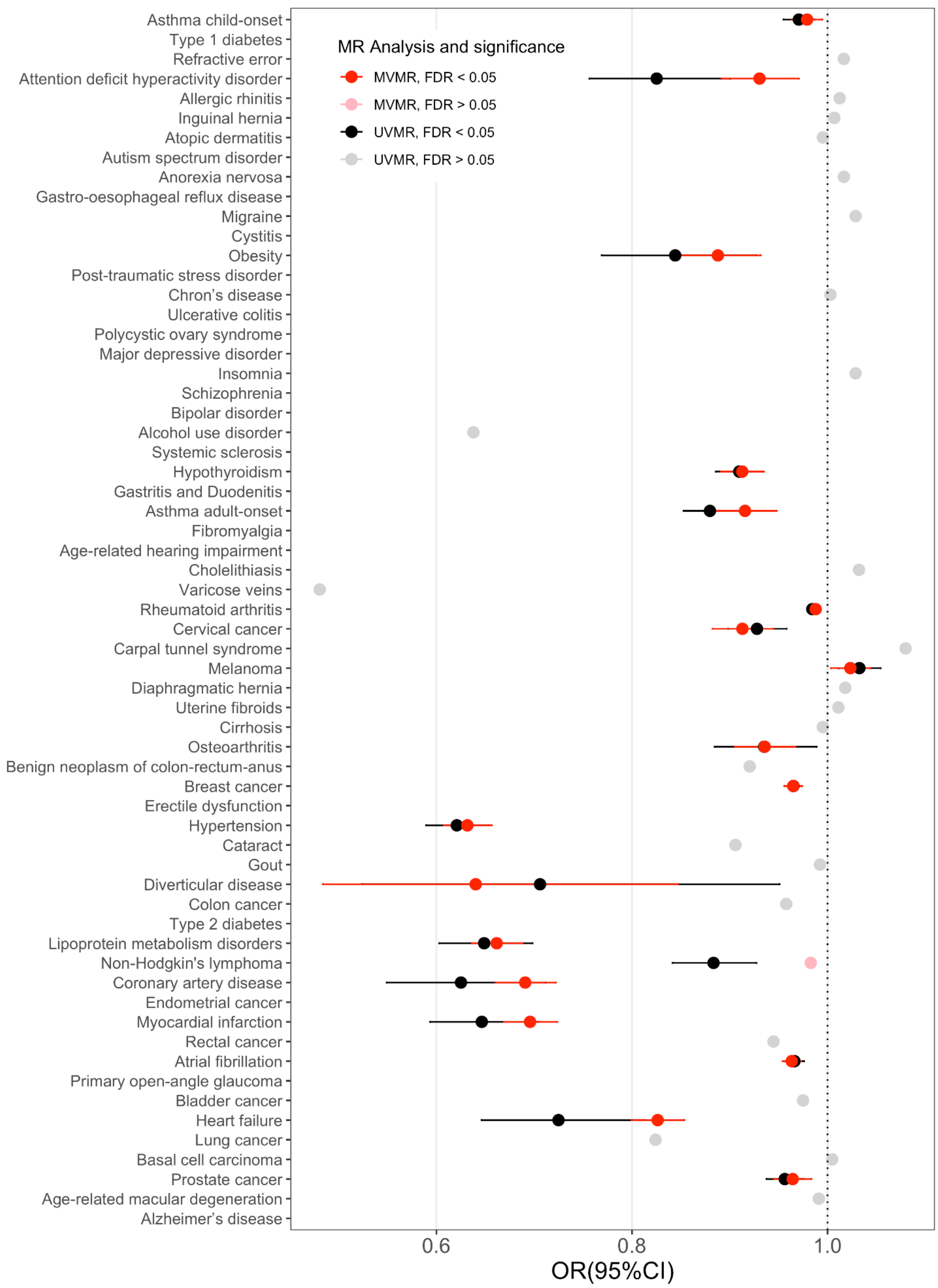

**Supplementary Figure 23. Mendelian Randomization analysis of the causal relationship between complex diseases and longevity corrected for multiple testing.** The x-axis displays odds ratios of causal estimates from the MR analysis assessing the impact of complex diseases (exposures) on longevity (outcome). Two MR approaches were employed. In the univariable MR, significant estimates (FDR < 0.05) are shown in black with confidence intervals (CIs), while non-significant estimates are depicted in gray without CIs for clarity. In the multivariate MR, which incorporates education as a mediator, significant estimates (FDR < 0.05) are shown in red, and non-significant estimates in lighter pink, also without CIs. For some diseases the analysis could not be performed, then no information appears in this plot.

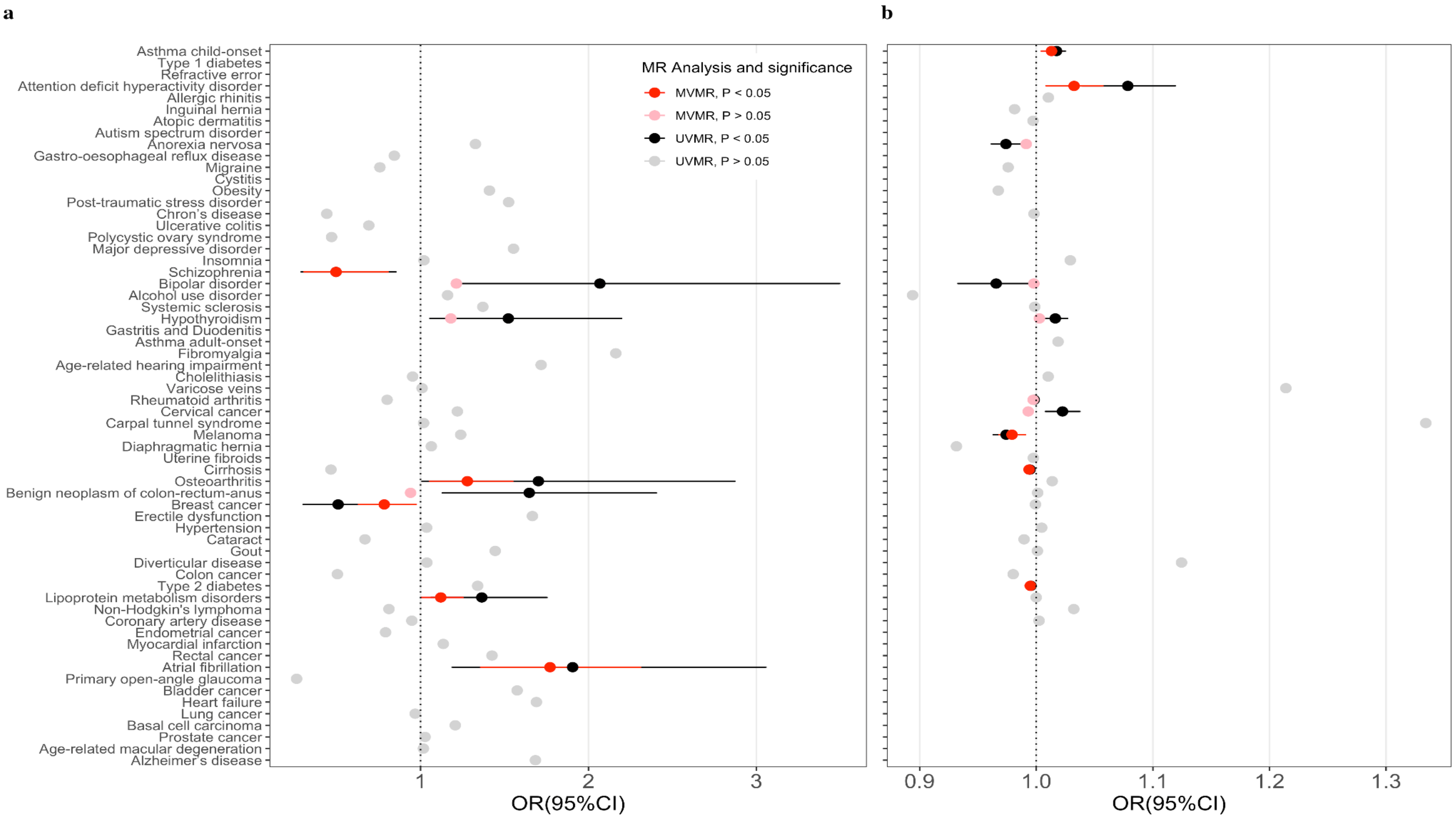

**Supplementary Figure 24. Mendelian Randomization analysis of the causal relationship between complex diseases and fertility.** The x-axis displays the odds ratios from MR analysis, which evaluates the causal effects of exposure on an outcome. We conducted a bidirectional analysis: Panel (a) shows the results when fertility is the exposure and causality is assessed on complex diseases, and Panel (b) presents the results when complex diseases are the exposure and causality is assessed on fertility. Two MR approaches were used. In the univariable MR, significant estimates ($P < 0.05$) are shown in black with confidence intervals (CIs), while non-significant estimates appear in gray without CIs for clarity. In the multivariate MR incorporating education as a mediator, significant estimates are depicted in red, and non-significant estimates in lighter pink, also without CIs. For some diseases, the analysis could not be performed; thus, no data are presented for those conditions.

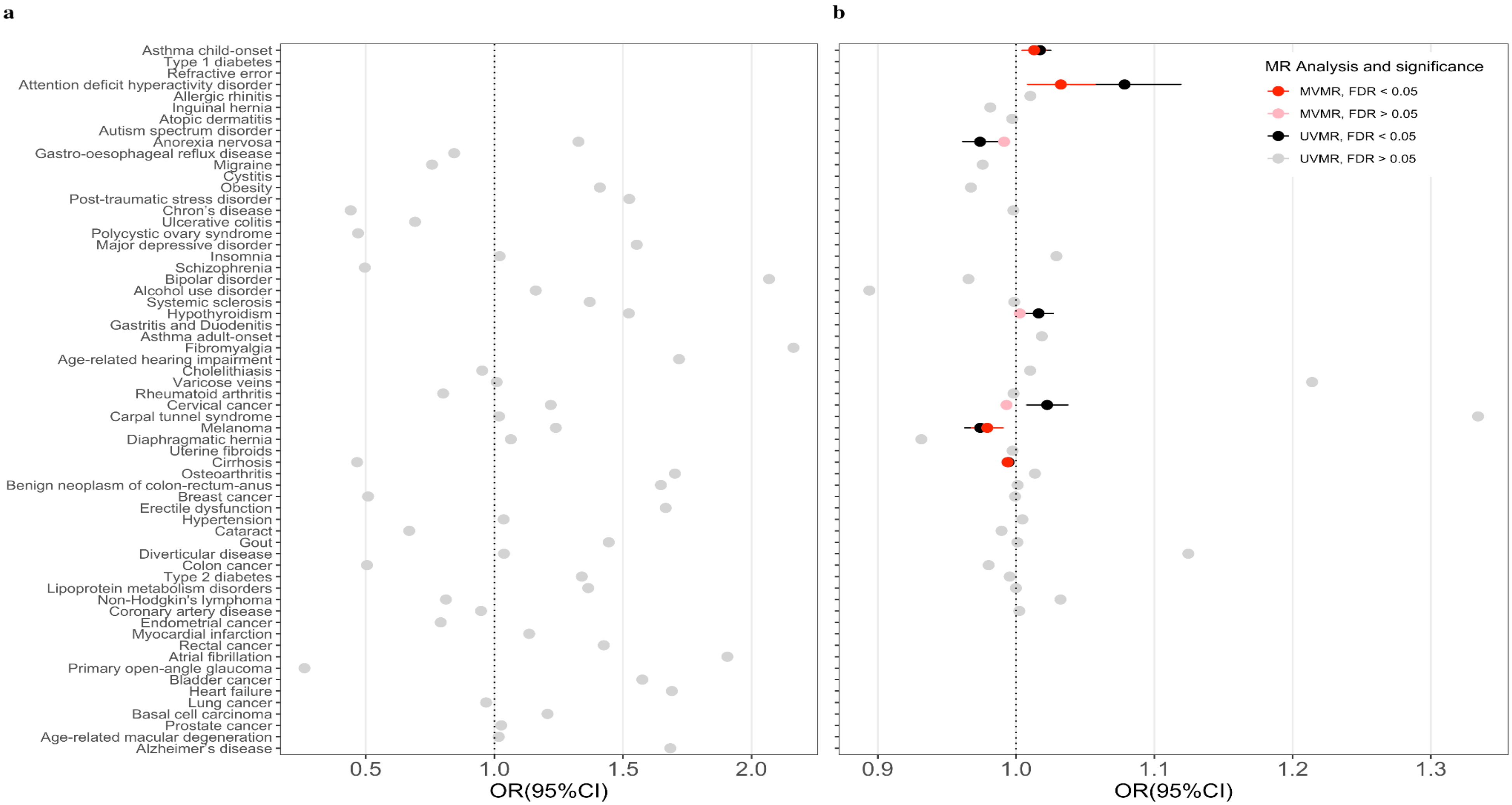

**Supplementary Figure 25. Mendelian Randomization analysis of the causal relationship between complex diseases and fertility corrected by multiple testing.** The x-axis displays the odds ratios from MR analysis, which evaluates the causal effects of exposure on an outcome. We conducted a bidirectional analysis: Panel (a) shows the results when fertility is the exposure and causality is assessed on complex diseases; to keep temporal plausibility diseases with onset ages < 5 years were not considered. Panel (b) presents the results when complex diseases are the exposure and causality is assessed on fertility; to keep temporal plausibility diseases with onset ages > 45 years were not considered. Two MR approaches were used. In the univariable MR, significant estimates (FDR < 0.05) are shown in black with confidence intervals (CIs), while non-significant estimates appear in gray without CIs for clarity. In the multivariate MR incorporating education as a mediator, significant estimates are depicted in red (FDR < 0.05), and non-significant estimates in lighter pink, also without CIs. For some diseases, the analysis could not be performed; thus, no data are presented for those conditions.

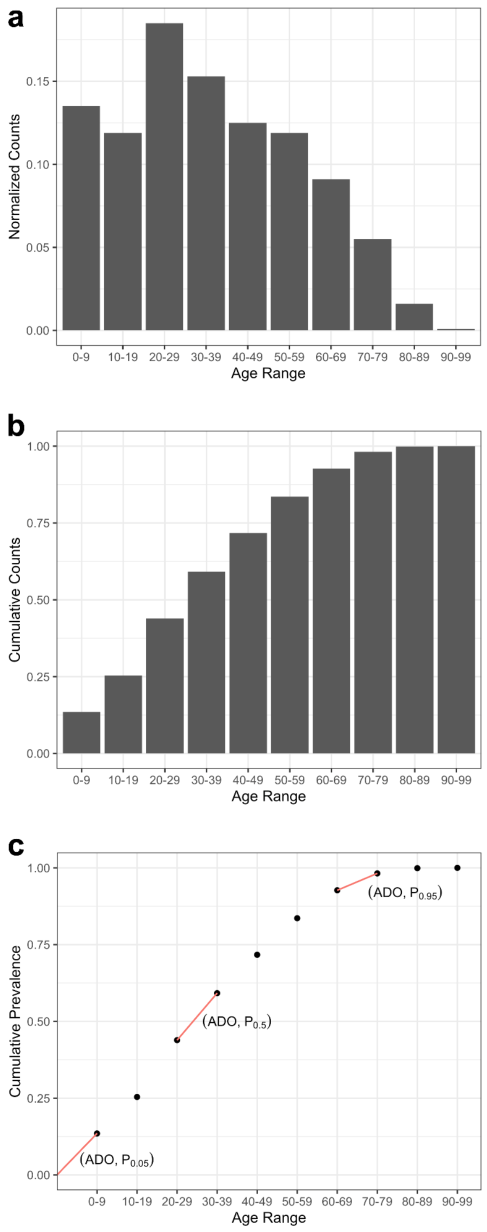

**Supplementary Figure 26. Prevalence distribution curves by age at first diagnosis for atopic dermatitis (ID: 1134)**. (a) Normalized prevalence curve for atopic dermatitis, where the x-axis represents the age ranges defined in Risteys, and the y-axis shows the normalized individual counts per age range. Each histogram bin reflects the proportionate prevalence for the corresponding age range. (b) Cumulative prevalence curve for atopic dermatitis, with the x-axis showing the age ranges and the y-axis representing the cumulative proportion of counts. The final bin represents the total cumulative prevalence, summing to 1. (c) Density prevalence curve. The red segments connecting two age ranges are the estimated linear interpolation used to calculate the age at disease onset (ADO) at different prevalence ratios: 5%, 50%, and 95%.

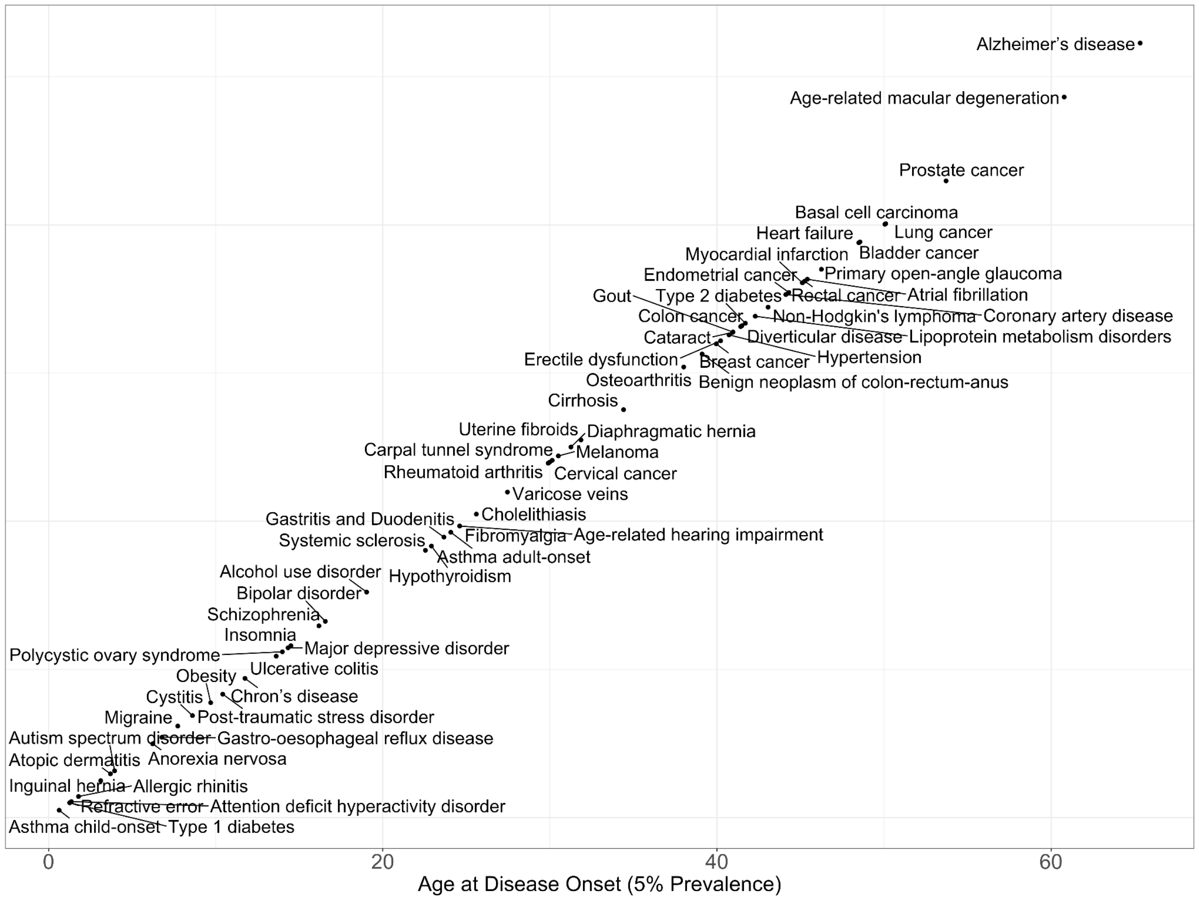

**Supplementary Figure 27**. **Age distribution of disease onset for 62 complex disorders**. This figure illustrates the onset age, defined as the highest age observed among the 5% of the youngest individuals with the disease. The x-axis represents the age distribution for disease onset, while the y-axis lists the diseases, arranged diagonally to enhance visual clarity and facilitate comparison. Childhood-onset asthma exhibits the earliest onset age among the studied disorders; and Alzheimer's disease has the latest onset age in the dataset.

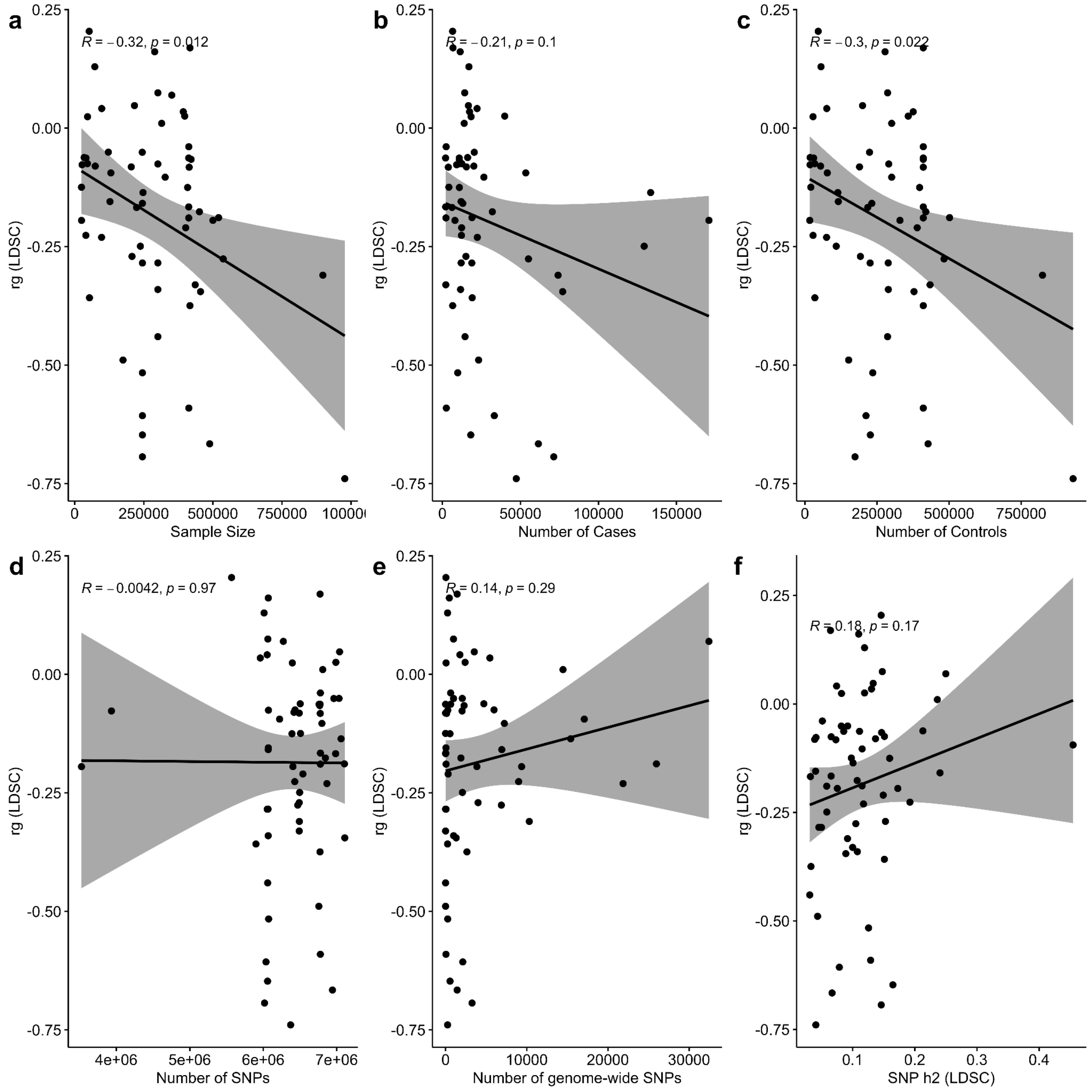

**Supplementary Figure 28**. **Evaluation of potential GWAS-related confounders in genetic correlations between complex diseases and longevity**. To account for potential biases in the observed genetic correlation effect sizes, Pearson correlations were computed across six key characteristics of the 62 GWAS datasets for complex diseases. These characteristics included: (a) sample size, (b) number of cases for 60 binary traits, (c) number of controls for 60 binary traits, (d) number of SNPs in the formatted summary statistics, (e) number of genome-wide significant SNPs, and (f) SNP-based heritability was calculated using LD Score Regression (LDSC), expressed on the liability scale for 60 binary traits and as total observed heritability for two continuous traits.

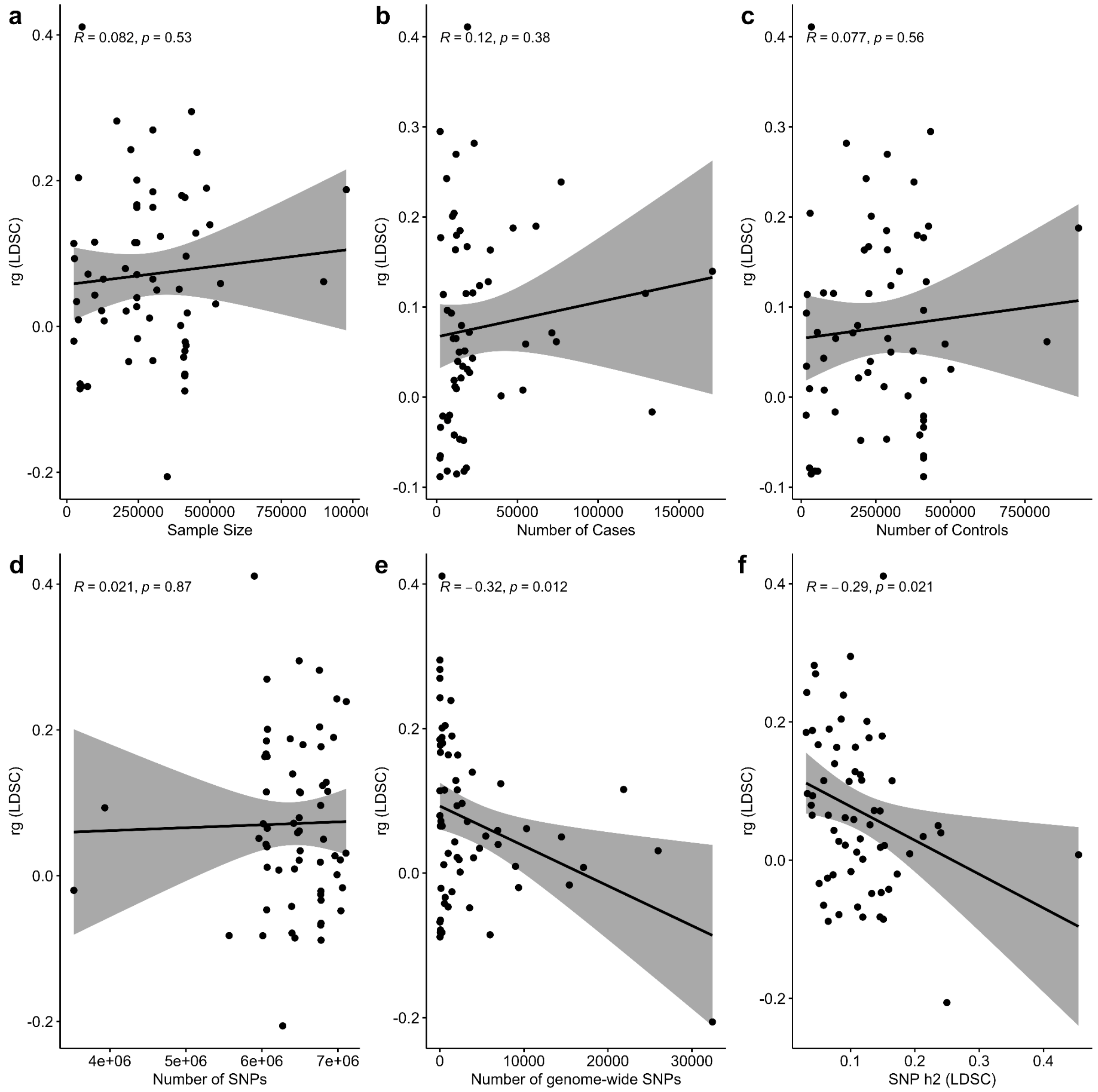

**Supplementary Figure 29**. **Evaluation of potential GWAS-related confounders in genetic correlations between complex diseases and fertility**. To account for potential biases in the observed genetic correlation effect sizes, Pearson correlations were computed across six key characteristics of the 62 GWAS datasets for complex diseases. These characteristics included: (a) sample size, (b) number of cases for 60 binary traits, (c) number of controls for 60 binary traits, (d) number of SNPs in the formatted summary statistics, (e) number of genome-wide significant SNPs, and (f) SNP-based heritability was calculated using LD Score Regression (LDSC), expressed on the liability scale for 60 binary traits and as total observed heritability for two continuous traits.

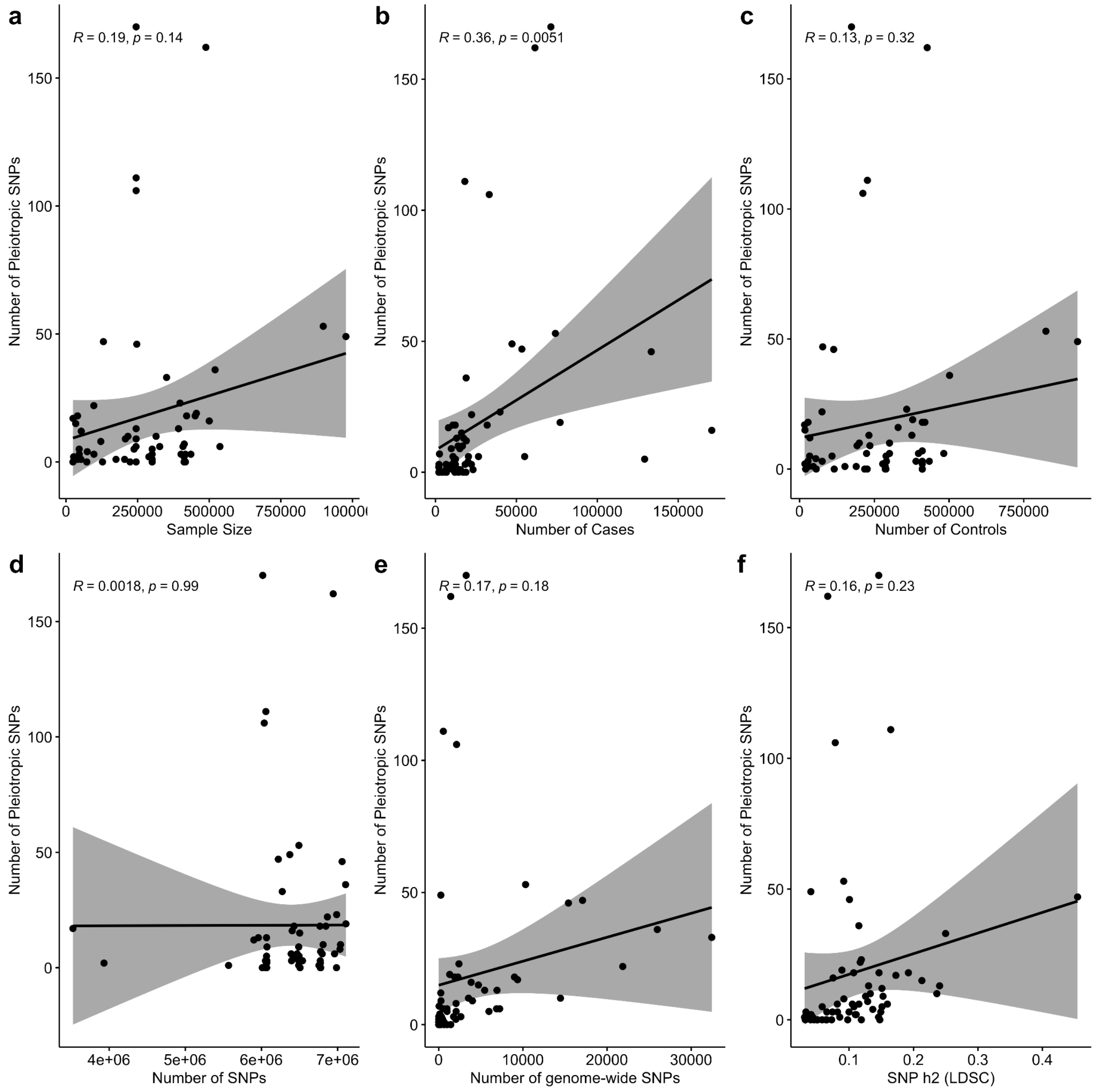

**Supplementary Figure 30**. **Evaluation of potential GWAS-related confounders in pleiotropy counts between complex diseases and longevity**. To account for potential biases in the observed number of pleiotropies, Pearson correlations were computed across six key characteristics of the 62 GWAS datasets for complex diseases. These characteristics included: (a) sample size, (b) number of cases for 60 binary traits, (c) number of controls for 60 binary traits, (d) number of SNPs in the formatted summary statistics, (e) number of genome-wide significant SNPs, and (f) SNP-based heritability was calculated using LD Score Regression (LDSC), expressed on the liability scale for 60 binary traits and as total observed heritability for two continuous traits.

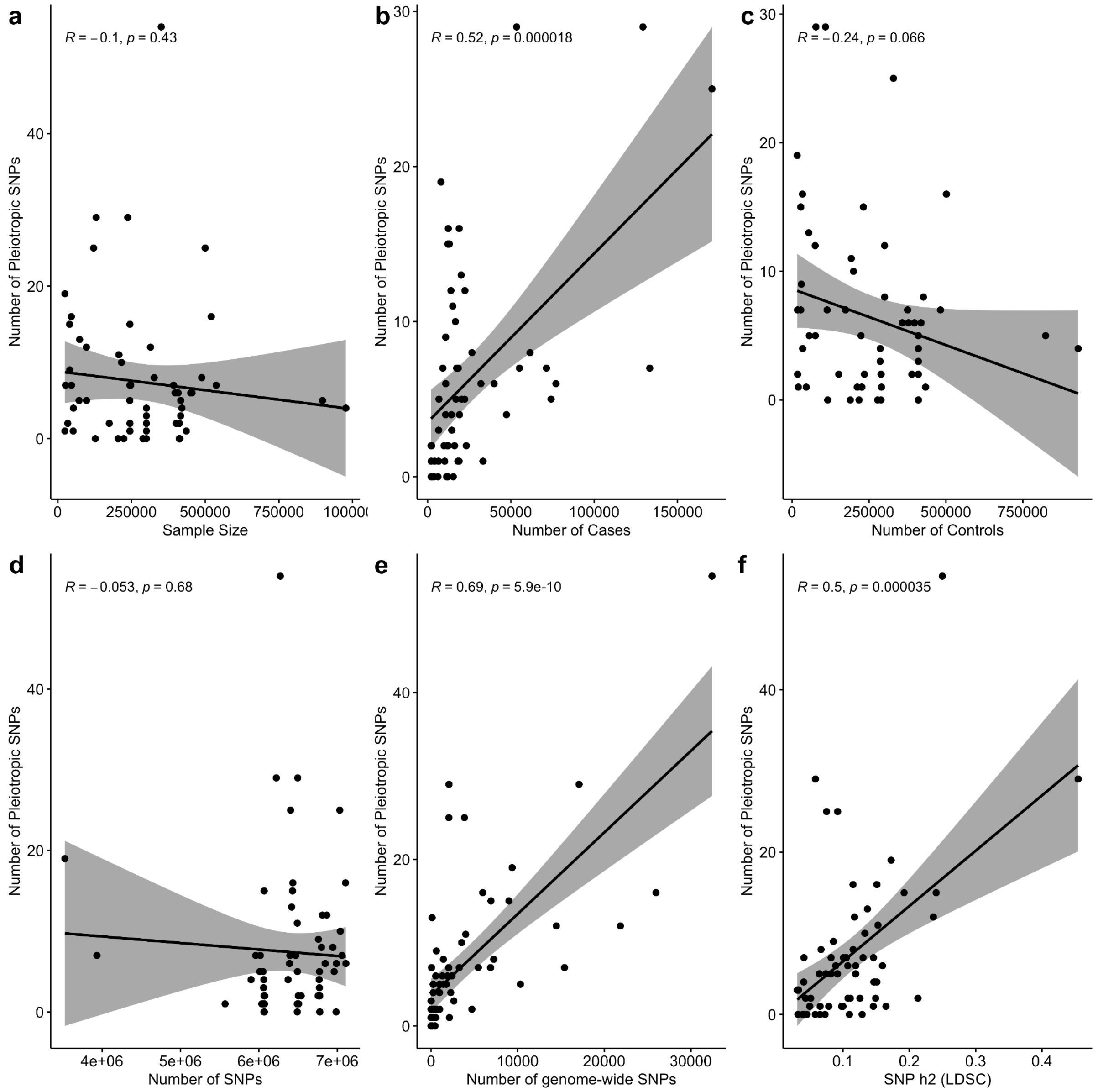

**Supplementary Figure 31**. **Evaluation of potential GWAS-related confounders in pleiotropy counts between complex diseases and fertility**. To account for potential biases in the observed number of pleiotropies, Pearson correlations were computed across six key characteristics of the 62 GWAS datasets for complex diseases. These characteristics included: (a) sample size, (b) number of cases for 60 binary traits, (c) number of controls for 60 binary traits, (d) number of SNPs in the formatted summary statistics, (e) number of genome-wide significant SNPs, and (f) SNP-based heritability was calculated using LD Score Regression (LDSC), expressed on the liability scale for 60 binary traits and as total observed heritability for two continuous traits.

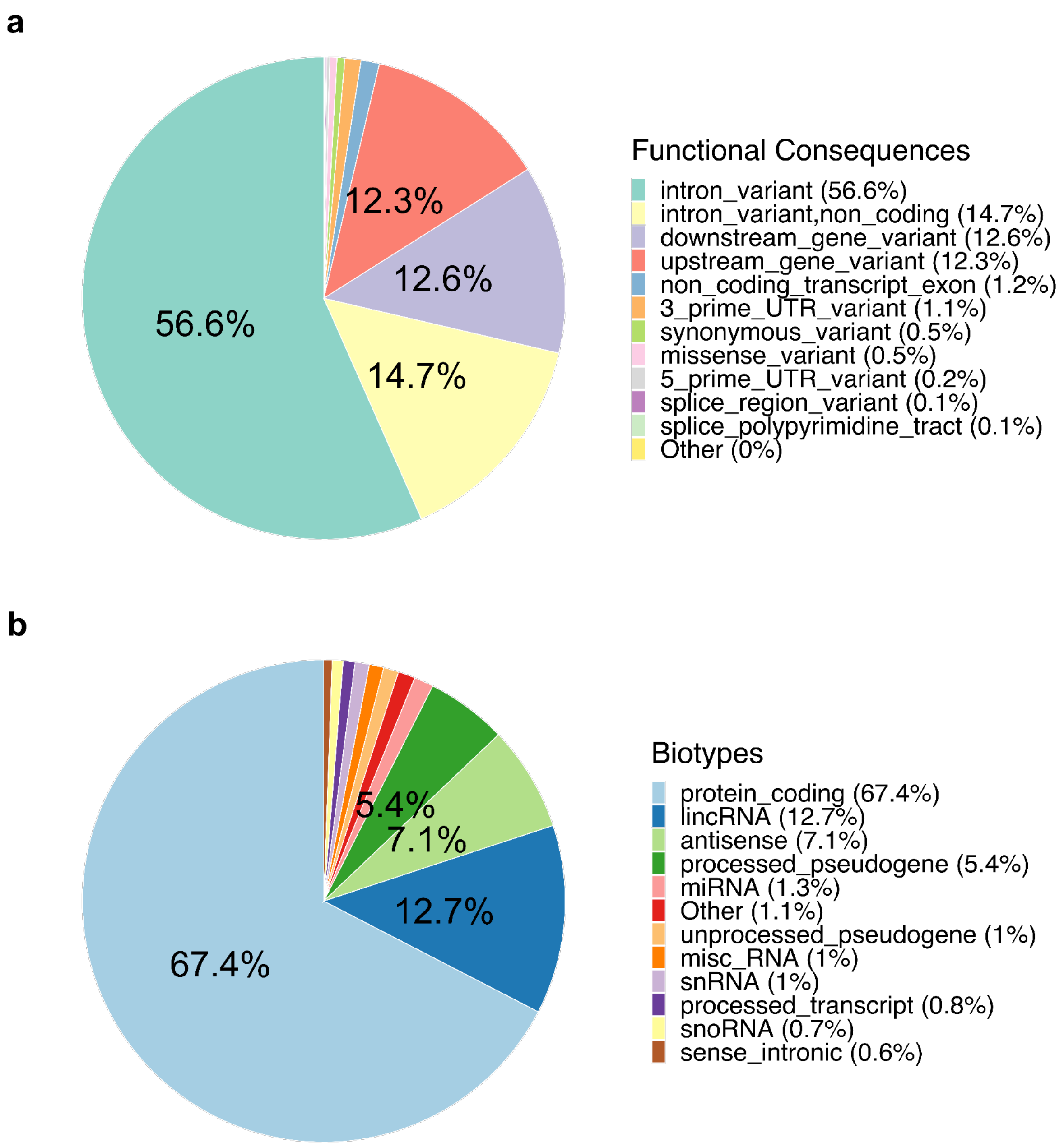

**Supplementary Figure 32.** Annotations of the 2.2 million SNP background set generated using the Ensembl Variant Effect Predictor (VEP). This background set corresponds to the common SNPs between the 62 complex diseases and the two life-history traits. a) Pie chart illustrating the distribution of functional variant consequences. b) Pie chart showing the distribution of variant biotypes. For clarity, only categories exceeding 5% of the total are labeled in each pie chart.

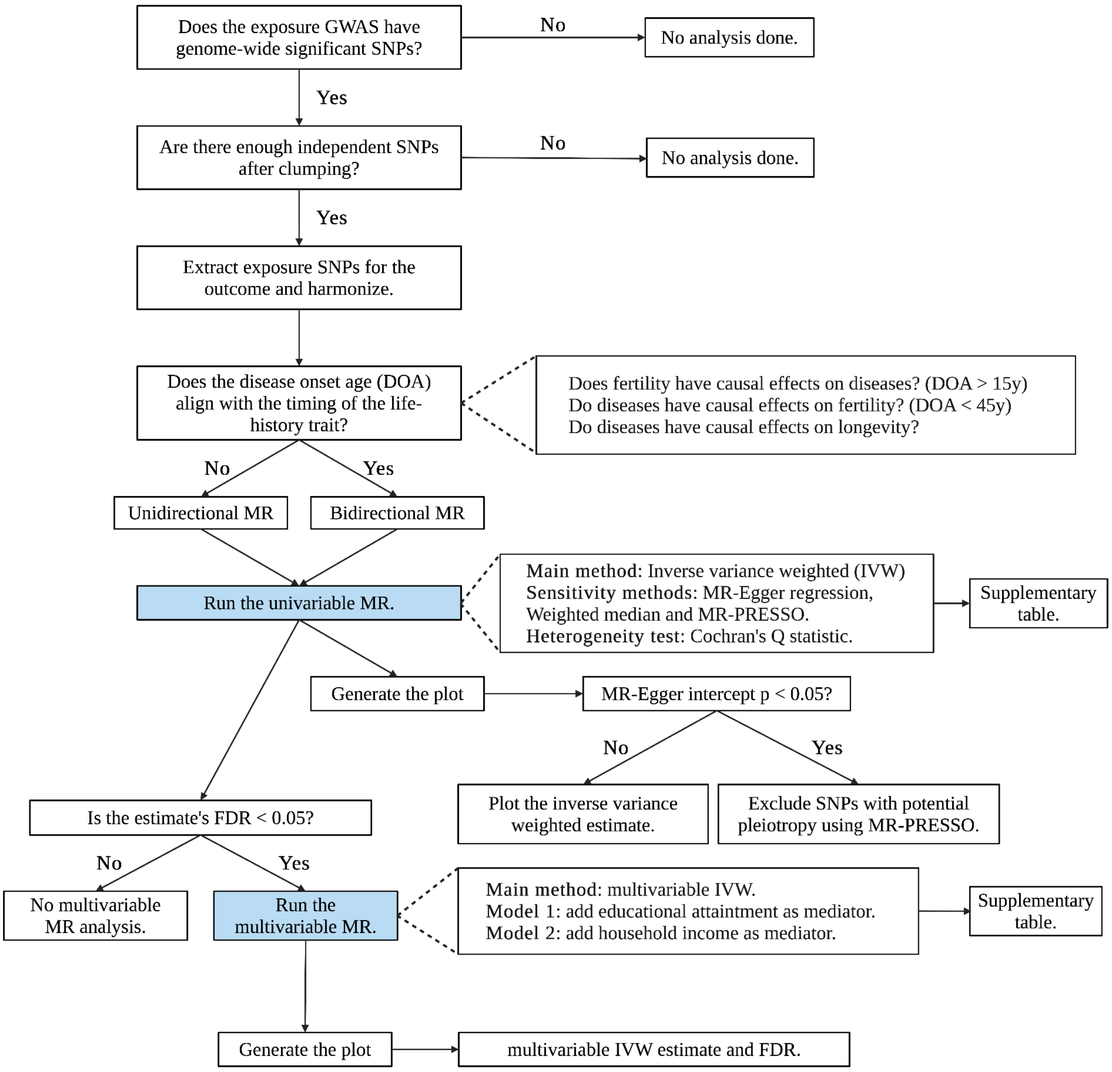

**Supplementary Figure 33**. Flow chart of the Mendelian randomization design.

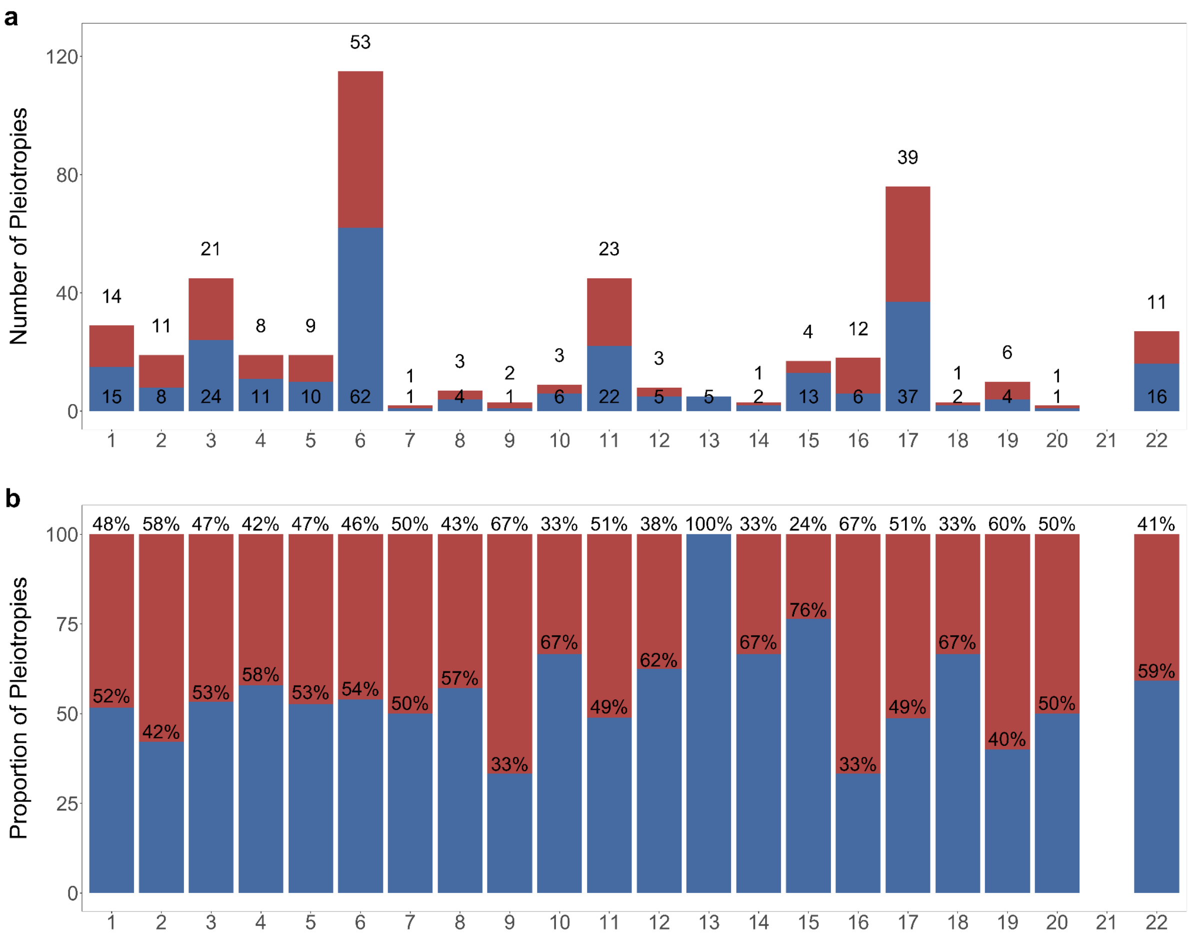

**Supplementary Figure 34. Chromosomal distribution of fertility-associated pleiotropies: counts and proportions.** Descriptive bar charts illustrating positive and negative pleiotropies with fertility by chromosome. Blue bars denote positive pleiotropies, whereas red bars indicate negative pleiotropies. a) The total counts of positive and negative pleiotropies within each chromosome. b) The relative proportions of positive and negative pleiotropies across chromosomes.

### Supplementary Note

### 1. Genome-wide association analysis of fertility

#### 1.1. The rationale for conducting a meta-analysis

In an evolutionary context, reproduction is a backbone component of Darwinian fitness and represents the potential of an individual to contribute with offspring in the next generation. Throughout this study we use the term fertility to denote the potential for reproduction.

We conducted a meta-analysis of three related measures on the number of offspring using the software METAL (Willer, Li, and Abecasis 2010). This approach enhances the statistical power to detect genetic variants with modest effect sizes, which is particularly beneficial for traits with smaller genetic contributions to phenotypic variance and where genome-wide association studies (GWAS) capture only a limited proportion of trait heritability (Manolio et al. 2009; Young 2019; Briley, Tropf, and Mills 2017).

This methodology is especially relevant for life-history traits such as fertility, as GWAS results are inherently context-dependent, capturing common genetic variance at a specific time point and for a particular population (Uffelmann et al. 2021). The importance of this context-specificity is accentuated by the dramatic changes in reproductive patterns observed over recent decades, particularly following the demographic transition (Kirk 1996; Galor 2012). In the scope of our research, we paid special attention to the fact that demographic transition varies across populations, with different regions experiencing unique fertility trajectories (Frejka 2016). Given this variability, we acknowledge that the UK Biobank served as the primary source of samples for the diverse GWASs on complex diseases and life-history traits utilized in this study (**Supplementary Table 14**). The UK Biobank is a large-scale biomedical database containing in-depth genetic and health information from half a million UK participants aged 40-69 at recruitment (Sudlow et al. 2015). When considering selection analyses this is also relevant because to infer polygenic adaptation, we used the singleton density score (SDS) that measures allele frequency changes in contemporary genomes from the UK10K Project (See Supplementary Methods 2.6) (Field et al. 2016). The use of similar data in both our primary GWAS analysis and the SDS method ensured consistency in our research approach and allowed for more robust conclusions about the evolutionary trade-offs between complex diseases and life-history traits.

#### 1.2. Meta-analysis of fertility related traits using METAL

Our study focused on individuals of European ancestry, combining genome-wide association data from three sources: the Social Science Genetic Association Consortium (SSGAC) study on the number of children ever born (Barban et al. 2016), and two UK Biobank studies analyzing the number of live births in females and the number of children fathered by males (Watanabe et al. 2019). We conducted a meta-analysis of these datasets using METAL software (version 2020-05-05) (Willer, Li, and Abecasis 2010) (**Supplementary Figure 1**). The analysis employed a sample size weighting scheme to account for varying sample sizes across input studies (**Supplementary Table 14**) and implemented overlap correction to address shared participants, primarily from the UK Biobank. Additionally, we applied genomic control to mitigate potential population stratification effects. The meta-analysis identified 5 associated loci, corresponding to 10 independent single nucleotide polymorphisms (SNPs) as outlined in **Supplementary Table 1**. Notably, all these loci were previously identified in GWAS of fertility-related traits (Mathieson et al. 2023). The computed SNP-heritability was determined to be 0.03 (1.4e-03). MAGMA tissue expression analysis implemented in FUMA revealed that the tissues most likely involved in the biological processes underlying fertility were within the brain (B=0.024, SE=0.006, P=0.00022) and ovary (B=0.031, SE=0.011, P=0.0013) (**Supplementary Figure 2a**) (Watanabe et al. 2017). The significant tissue-specific gene expression patterns show a particularly strong enrichment in the brain anterior cingulate cortex (B=0.024, SE=0.007, P=0.00027), the amygdala (B=0.026, SE=0.007, P=0.00038) and the hypothalamus (B=0.026, SE=0.008, P=0.00041) (**Supplementary Figure 2b**). This finding aligns with previous research highlighting the importance of neuroendocrine pathways in reproductive behaviors. Key reproductive processes, such as puberty and the menstrual cycle, are governed by steroid hormones, which are tightly regulated by the hypothalamus-pituitary-gonadal (HPG) axis (Garg and Berga 2020; Parhar, Ogawa, and Ubuka 2016). Prioritized genes were the estrogen receptor 1 (ESR1), C2H2C-type zinc finger transcription factor (ST18) and cell adhesion molecule 2 (CADM2) (**Supplementary Table 1**). ESR1 and CADM2 are protein-coding genes with a very high probability of being loss-of-function intolerant (pLI>0.98) which mean that these are likely essential genes that might be under strong evolutionary pressures as mutations in these genes might lead to negative consequences that reduce reproductive fitness. Notably, the analysis of tissue-specific expression patterns for prioritized genes revealed that ESR1 exhibited high expression levels in several reproductive tissues, including the fallopian tubes, uterus, vagina and cervix. Gene information for ESR1

(Gene ID: 2099) from the National Library of Medicine (NLM) at the National Institutes of Health (NIH) reports numerous transcript variants resulting from alternative promoters and splicing events. The protein encoded by ESR1, a ligand-activated transcription factor, regulates many estrogen-induced genes involved in critical reproductive functions, including gestation and sexual development. Polymorphisms in ESR1 have been associated with measures of ovarian function quality, particularly in response to assisted reproduction techniques (de Mattos et al. 2014). Furthermore, ESR1 variants are implicated in the susceptibility to diseases such as breast cancer and endometriosis, highlighting its role in both reproductive health and disease pathology (Jeselsohn et al. 2015; Lamp et al. 2011). In contrast, ST18 and CADM2 showed elevated expression across multiple brain regions (**Supplementary Figure 3**). CADM2, which encodes a member of the synaptic cell adhesion molecule 1 (SynCAM) family, plays a crucial role in synapse organization. CADM2 has been implicated in the development of endometriosis, as well as impulsive personality traits and other behavioral characteristics (Z. Wang et al. 2024; Sanchez-Roige et al. 2023). These tissue-specific expression patterns provide valuable insights into the potential functional pathways through which genetic variants may influence fertility, highlighting both nervous system and reproductive system involvement.

Additionally, to validate our findings, LD score regression was conducted to evaluate the genetic correlation between our GWAS meta-analysis and the latest study on reproductive success (Mathieson et al. 2023), demonstrating a strong genetic correlation of nearly 1 ($r_g$=0.9974, SD=7.4e-03).

#### 1.3. Polygenic score validation in an independent sample

To validate our fertility meta-analysis GWAS, we constructed a polygenic score (PGS) using PRScs, a Bayesian regression framework that infers posterior effect sizes of SNPs using GWAS summary statistics and an external linkage disequilibrium reference panel (Ge et al. 2019). As European reference samples, we used data from the UK Biobank (application number: 67292) after standard genetic quality control. To evaluate the performance of the fertility PGS, we applied it to an independent European-ancestry cohort from the Database of Genotypes and Phenotypes (dbGaP), specifically the GENEVA Genome-Wide Association Study of Venous Thrombosis (phs000289, study accession number 33199, n = 2,555).

We only included female participants aged 40 years and older, as the available fertility measures in the independent cohort were based on several pregnancy outcomes. Very young participants were excluded to minimize age-related bias in the number of children, while still striving to maximize the sample size.

Genetic data underwent rigorous quality control using PLINK, which included ancestry verification, checks for missingness, minor allele frequency threshold (MAF<0.005) sex discrepancies, deviations from Hardy-Weinberg equilibrium, heterozygosity levels, and relatedness (Chang et al. 2015; Marees et al. 2018). After quality control, 988 samples and 523,900 positions remained. Imputation was performed on the TOPMed server using the TOPMed R3 (hg38) reference panel. We applied an imputation quality filter (rsq≥0.3) and subsequently excluded variants with MAF<0.001, resulting in 11,262,644 positions (Das et al. 2016).

The PGS was calculated on the imputed data by summing the weighted risk alleles, with weights derived from our fertility meta-analysis. To assess the predictive power of this fertility-PGS, we tested its association with three reproductive outcome measures: number of live-birth pregnancies (NLB), number of completed pregnancies (NCP), and number of times pregnant (NTP). Linear regression models were fitted for each outcome, adjusting for age, missingness and the first 10 genetic principal components to account for population stratification.

As expected, our analysis revealed that age was the strongest predictor across all reproductive measures. Additionally, we observed a degree of population stratification captured by the eighth principal component (PC8), which contributed to explaining variation in all three pregnancy-related measures (**Supplementary Table 2**). Notably, the polygenic score demonstrated predictive utility for two outcomes: the number of completed pregnancies (NCP, P=0.0448) and the number of live-birth pregnancies (NLB, P=0.0419). However, its association with the total number of times pregnant (NTP) did not reach statistical significance (P=0.0725). In terms of overall model performance, the adjusted $R^2$ values were approximately 11% for NLB and NCP, indicating a modest but meaningful proportion of variance explained. For NTP, the adjusted $R^2$ was slightly lower at around 8%. These findings suggest that while the PGS contributes to the prediction of reproductive outcomes, its explanatory power remains limited compared to non-genetic factors such as age.

In line with the objectives of our analysis, the observed association between the polygenic score and complementary pregnancy outcomes in an independent cohort indicates that the identified genetic variants collectively help explain variation in fertility-related phenotypes. Notably, the predictive strength of the PGS is more pronounced for biologically meaningful outcomes, such as live births (NLB) and completed pregnancies (NCP), than for the total number of pregnancies (NTP). This finding suggests that our fertility meta-analysis captures genetic variation linked not only to reproductive potential but also to measures that reflect a successful gestation and healthy offspring.

#### 1.4. Human reproductive success phenotype definition

Fertility is inherently difficult to study from a genetic perspective. It is highly polygenic, influenced by numerous small-effect variants, and its definition and measurement are not straightforward (Barban et al. 2016; Benonisdottir et al. 2024). Demographic changes, most notably since the second demographic transition, illustrate how quickly fertility patterns can shift: in just a few generations, human populations have moved from relatively large family sizes to near or below replacement-level fertility (Frejka 2016; Sobotka 2017). This rapid change far outpaces any corresponding shift in the underlying genetic architecture, which largely remains the same (Hayward and Sella 2022). As a result, environmental and socio-cultural factors exert substantial influence on fertility, highlighting its plasticity (Abdellaoui et al. 2025). Moreover, in many industrialized societies, delayed childbearing often reduces total offspring to around two per couple—vastly different from pre-industrial contexts where families were typically much larger (Brewster and Rindfuss 2000). These contrasts underscore the practical challenges for genome-wide association studies (GWAS). When phenotypic variance is low and the trait is highly polygenic, detecting significant genetic signals becomes more difficult, as individual variants tend to have small effect sizes (Uffelmann et al. 2021).

The following topic has been extensively discussed elsewhere (Barban et al. 2016), but a summary is provided here for completeness. In a biological sense, fertility, measured as the number of offspring, is linked to "fitness," or reproductive success. However, reproductive success itself is shaped by multiple components like biological fecundity (Zietsch et al. 2014), reproductive behaviour (Alvergne, Jokela, and Lummaa 2010; Jokela et al. 2011), individual choices (Tanturri and Mencarini 2008), socioeconomic factors (M. Mills et al. 2011), sex-specific genetic effects or gene-environment interactions (Barban et al. 2016). Together, these elements create a phenotype that is both multifaceted and context dependent.

Twin studies further suggest that broad-sense heritability can range from approximately 15% to 45%—a valuable clue for understanding how much of fertility is genetically determined (M. C. Mills and Tropf 2015). However, as with other highly polygenic traits, a gap often remains between broad-sense heritability and the heritability captured by standard GWAS. A GREML-based study on the number of children ever born—partially overlapping with the cohort used here—reported no detectable dominance effects, with additive genetic variation explaining around 10% of the phenotypic variance (Tropf et al. 2015; Barban et al. 2016). Although modest, such estimates still offer hope that GWAS can capture a meaningful, if incomplete, portion of the genetic component of fertility.

We sought to elucidate the shared genetic architecture between our fertility meta-analysis and a range of fertility-related traits, aiming to enhance the biological interpretation of our phenotype. To this end, we leveraged publicly available GWAS summary statistics from the GWAS Atlas and categorized the selected traits into three domains: (1) biological aspects of reproduction, (2) reproductive behavior, and (3) socioeconomic status indicators.

For traits related to the biology of reproduction, we observed positive genetic correlations between fertility and both vitamin D levels (atlas ID: 3981, $r_g$=0.17, P=1.9e-02, FDR=2.41e-02) (Jiang et al. 2018) and age at voice broke (atlas ID: 3325, $r_g$=0.07, P=2.19e-02, FDR=2.41e-02) (Watanabe et al. 2019). In contrast, age at menopause exhibited negative genetic correlations with fertility (atlas ID: 3366, $r_g$=-0.07, P=4.1e-03, FDR=6.15e-03) (Watanabe et al. 2019). And age at menarche showed no statistically significant genetic correlation (atlas ID: 3339, $r_g$=0.01, P=5.3e-01, FDR=5.32e-01) (Watanabe et al. 2019). These findings are consistent with findings linking vitamin D levels to reproductive health in both men and women (Lerchbaum and Obermayer-Pietsch 2012).

Within the reproductive behavior domain, higher number of sexual partners (atlas ID: 3305, $r_g$=0.27, P=2.2e-02, FDR=2.41e-02) and ever-use of contraceptives (atlas ID: 3346, $r_g$=0.17, P=2.2e-02, FDR=6.15e-03) were genetically correlated with enhanced fertility, whereas older age at first sexual intercourse (atlas ID: 3304, $r_g$=-0.53, P=1.37e-140, FDR=1.65e-139), older age at first birth (atlas ID: 3343, $r_g$=-0.67, P=3.3e-137, FDR=1.99e-136), and younger age at last contraceptive (atlas ID: 3305, $r_g$=-0.51, P=1.08e-20, FDR=3.23e-20) were associated with higher fertility. These patterns suggest that delayed reproductive behaviors are associated with reduced fertility, while a more active sexual life is linked to higher fertility. Such associations have been reported in previous demographic studies (Guzzo 2014; Sobotka 2017).

Regarding socioeconomic factors, we found that higher fertility was genetically correlated with lower educational attainment (atlas ID: 4066, $r_g$=-0.30, P=3.47e-58, FDR=1.39e-57) (J. J. Lee et al. 2018), reduced household income (atlas ID: 3195, $r_g$=-0.15, P=8.2e-08, FDR=1.97e-07), and greater material deprivation as measured by the Townsend deprivation index ($r_g$=0.27, P=5.38e-07, FDR=1.07e-06) (Hill et al. 2016). These results are in line with established demographic trends, which have demonstrated inverse associations between fertility and indicators of socioeconomic advantage (Martín 1995; Skirbekk 2008).

### 2. Methods and analytical procedures

#### 2.1. International classification of diseases

The International Classification of Diseases (ICD) is a standardized system developed by the World Health Organization (WHO) for recording, reporting, and analyzing health conditions and causes of death (World Health Assembly 1990). It serves as a global standard for diagnostic health information, enabling consistent tracking of diseases, epidemiological research, and healthcare planning across different countries and healthcare systems. First introduced in the 19th century, the ICD has undergone multiple revisions to incorporate advances in medical science and to address changing healthcare needs.

In this study, we used the 10th revision of the ICD, commonly referred to as ICD-10, which was released in 1994 and has since been widely adopted internationally (Hirsch et al. 2016). The ICD-10 provides a hierarchical framework for coding diseases, disorders, injuries, and other health-related conditions. It organizes diseases into 21 main chapters based on specific body systems or general disease categories, with each chapter further subdivided into blocks of related conditions. For example, Chapter I includes "Certain infectious and parasitic diseases" (codes A00–B99), while Chapter IX covers "Diseases of the circulatory system" (codes I00–I99).

Each health condition in the ICD-10 is assigned a unique alphanumeric code consisting of a letter followed by up to four digits (e.g., E10 for type 1 diabetes mellitus). This coding structure allows for the classification of over 14,000 individual conditions and facilitates the identification of comorbidities and disease patterns in large datasets.

We obtained ICD-10 codes by consulting the definitions and related information provided in the original GWAS studies. Whenever the original study explicitly reported ICD-10 codes—whether one or multiple—we used those directly. In cases where the codes were not specified, we referred to the phenotype definition and description to identify the most appropriate ICD-

10 code. Refer to **Supplementary Table 14** for a complete description of the ICD-10 codes by disease, along with related information.

ICD-10 codes are widely used in clinical settings, research, and public health initiatives. In this study, they were instrumental in defining and categorizing disease phenotypes for analysis. The systematic classification provided by the ICD-10 ensured consistency in disease definitions, enabling robust comparison and aggregation of health data across different studies and biobanks.

#### 2.2 Age at disease onset

##### FinRegistry Release 11, a nationwide health data resource

Risteys, is a publicly available web portal developed as part of the FinnGen project. It allows users to explore Finnish health registry data and FinnGen GWAS results at the phenotypic level.

Risteys enables users to explore clinical endpoint definitions and review descriptive statistics derived from both FinnGen and FinRegistry. FinnGen is a public-private research initiative that combines genomic data from Finnish biobanks, encompassing around 500,000 individuals that represents around 10% of Finland's population (Kurki et al. 2023). In contrast, FinRegistry is a curated dataset integrating nationwide registry data for approximately 7.2 million individuals (Viippola et al. 2023). FinRegistry was selected over FinnGen because it provides broader population coverage, enabling more accurate estimates of both population prevalence and ages at disease onset. The larger sample size allowed us to assess prevalence levels across each decade of life from 0 to 90 years, thereby improving the representation of true population-level disease patterns and reducing potential selection biases.

To estimate the age of onset for each disease, we utilized prevalence data from the Risteys web application (https://r11.risteys.finregistry.fi/), using the R11 release accessed in August 2023. Because the disease endpoints in Risteys are harmonized with international disease ontologies, we could easily align our phenotypes with the clinical endpoints via the ICD-10 codes. For most diseases, a single ICD-10 code defines the phenotype. However, for certain diseases, multiple codes were used in the original GWAS to select the case participants. In these instances, we aggregated incidence data across all relevant ICD-10 codes to provide a comprehensive view of the disease onset age. **Supplementary Table 14** details the mapping of disease phenotypes, their associated ICD-10 codes, and the corresponding Risteys endpoints.

#### Estimating the age at disease onset

Prevalence data in Risteys are presented in a histogram format, showing the age ranges that define each bin and the corresponding sample counts within these ranges. These sample counts represent the number of individuals diagnosed with a particular condition for the first time within each age range. To calculate the age at disease onset, we first normalized the prevalence values for each disease. The normalized prevalence was calculated by dividing the prevalence of each age group by the total prevalence. Thus, normalized prevalence values are the relative proportions per age group (**Supplementary Figure 26a**). The cumulative prevalence was then calculated by adding the normalized prevalence values across all age groups. Thus, for the last age range, cumulative prevalence equals 1 (**Supplementary Figure 26b**).

**We defined the onset age as the highest age observed among the 5% of the youngest individuals with the disease** (Jia et al. 2019). Following the example on atopic dermatitis, the age at disease onset at 5% prevalence was between the ages of zero and nine years old. After constructing the cumulative prevalence curve, we could use it to identify the age range that corresponds to 5% prevalence (**Supplementary Figure 26c**).

However, as ages are coded in ranges, the estimation of a particular onset age requires linear interpolation. We used the following mathematical expressions to obtain the age at disease onset:

$$(1)\ \Delta P = P_X - P_I$$

$$(2)\ \Delta A = ADO - A_I$$

$$(3)\ \Delta P = \Delta A \cdot m$$

$$(4)\ m = \frac{(P_F - P_I)}{(A_F - A_I)}$$

$$(5)\ ADO = (P_X - P_I) \cdot \frac{(A_F - A_I)}{(P_F - P_I)} + A_I$$

where $P_X$ is the prevalence percentage of interest and ADO is the age at disease onset for the corresponding prevalence percentage.

To determine the age of disease onset, we first identified the age range at which each disorder reached a 5% prevalence in the population. For atopic dermatitis, at 10 years old, 13.5% of samples have been diagnosed with the disease. Therefore, the initial age and prevalence represented as ($A_I$, $P_I$) in the linear interpolation mathematical expression were zero (0, 0). While the final age and prevalence, represented as ($A_F$, $P_F$) in the linear interpolation mathematical expression, were (10, 0.135). Resolving the formula gave an age at disease onset of 3.7 at 5% prevalence for atopic dermatitis. **Supplementary Table 14** contains comprehensive data on the age at disease onset for all 62 diseases analyzed. Notably, the onset ages corresponding to a 5% prevalence threshold vary considerably among diseases, thereby offering a robust approach for accounting for age-specific effects (**Supplementary Fig. 27**). The diverse age-of-onset patterns across diseases offer unique opportunities to investigate the complex interplay between biological aging, disease-specific mechanisms, and reproductive pathways. These natural experiments illuminate how pleiotropic genes may mediate trade-offs between longevity, health, and fertility across the human lifespan.

We acknowledge the limitations of using a single prevalence threshold to determine disease onset, as this approach does not fully capture the complexity of disease progression. Prevalence curves can vary significantly between diseases; some conditions may reach the 5% threshold rapidly and continue to increase sharply while others might plateau near the threshold for extended periods. While our method does not account for these nuanced progression characteristics, it serves a specific purpose: to provide a standardized framework for comparing onset patterns across a diverse spectrum of complex diseases. This approach offers several advantages, including the simplicity in replication and interpretation, and broad applicability to a wide range of diseases, facilitating large-scale comparative analyses. Our primary goal was to classify diseases using a consistent metric, enabling insights into age-related disease susceptibility patterns.

##### FinRegistry: a comprehensive model for European disease prevalence estimation

Ages at disease onset were calculated using Finland as the reference population. However, individuals included in the GWAS studies are of European ancestry. European ancestry includes, but is not limited to, Finnish populations. Hence, we validated the obtained ages with the GHDx data from the General Burden of Disease (GBD) study (Christopher J L Murray 2020). GHDx data comprises the world's most comprehensive catalog of surveys, censuses, vital statistics, and other health-related data. For our purposes, we used prevalence data from the Western European region, for both sexes, and from the year 2019 (https://vizhub.healthdata.org/gbd-results/).

GHDx prevalence data was obtained for twenty different age-groups: < 1 year, 1-to-4, 5-to-9, 10-to-14, 15-to-19, 20-to-24, 25-to-29, 30-to-34, 35-to-39, 40-to-44, 45-to-49, 50-to-54, 55-to-59, 60-to-64, 65-to-69, 70-to-74, 75-to-79, 80-to-84, 85-to-89 and 90-to-94. Here, the age ranges were different from those in Risteys, but the way of computing the age at disease onset followed the same rationale. From normalized prevalence values, we generated a cumulative prevalence curve and used linear interpolation to calculate the age at disease onset for 5% prevalence ratios. Out of the 62 diseases examined, GHDx only had data for 31 of them. A Pearson correlation analysis showed a strong positive relationship ($r = 0.92$, $p = 1.8e-13$) between the 5% prevalence rate estimates from FinRegistry and GHDx. The high concordance between the 5% prevalence-based onset ages estimated from the Finnish FinRegistry and those derived from Western European GHDx data validates our approach across European populations.

#### 2.3. Genetic Correlation

We employed LD score regression (LDSC) to evaluate the genetic overlap between life-history traits and complex diseases, thereby elucidating their shared genetic architecture and potential underlying pleiotropy (Bulik-Sullivan et al. 2015). Using the LDSC tool (https://github.com/bulik/ldsc), we estimated SNP heritability for the 62 complex diseases and calculated pairwise genetic correlations between life-history traits and these diseases. In our analyses, we utilized precalculated linkage disequilibrium (LD) scores derived from 1000 Genomes Project European (1000G EUR) populations, while excluding the MHC region (25–34 Mb) due to its complex LD patterns. LDSC employs a regression framework where the product of z-scores from two GWAS are regressed against LD scores for each SNP. The resulting slope of this regression provides an estimate of the genetic covariance, which is then normalized to yield the genetic correlation between the traits. Simultaneously, the intercept of this regression is used to account for potential biases arising from sample overlap between the two GWAS and residual population stratification, ensuring that the genetic correlation estimate is not inflated by these confounding factors (Bulik-Sullivan et al. 2015).

For all datasets, we maintained the original number of SNPs after quality control and transformed odds ratios (ORs) and beta coefficients from the summary statistics to z-scores for consistency. For binary traits, we computed SNP heritability on the liability scale using population prevalence data from the FinRegistry 11th release and derived sample prevalence from GWAS case numbers (Viippola et al. 2023). GWAS disease phenotypes were matched to FinRegistry endpoints using the standardized codes from the International Classification of Diseases (ICD-10) (Hirsch et al. 2016).

To evaluate potential confounding factors, we calculated Pearson correlations between the genetic correlations and various GWAS characteristics, including age at disease onset, sample sizes, number of cases, SNP counts, number of genome-wide SNPs, and SNP-based heritability. Finally, to address potential biases in the association between common genetic etiology and age at disease onset, we performed a leave-one-out cross-validation analysis, iteratively removing each disease and repeating the analysis. This approach ensured the robustness of our findings by mitigating the influence of any single disease on the overall results.

#### 2.4. Pleiotropy

The pleiotropy-informed false discovery rate (pleioFDR) method is a statistical approach designed to identify pleiotropic loci—genetic regions associated with multiple phenotypes—by leveraging shared genetic signals between traits. It extends traditional false discovery rate (FDR) approaches by incorporating information from multiple phenotypes to improve the detection of true associations while controlling for false positives. This method is particularly effective for complex traits, where overlapping genetic architectures are common (Andreassen et al. 2013).

The conditional FDR is calculated as the posterior probability that a given SNP is null for one phenotype, conditioned on its association with another phenotype. This conditioning allows the method to prioritize SNPs that exhibit strong associations with both traits, thus enhancing the statistical power to detect pleiotropic loci. By incorporating this additional layer of information, pleioFDR refines the identification of SNPs with shared effects across phenotypes, increasing confidence in their biological relevance (Andreassen et al. 2013).

The conjunctional FDR extends this approach by computing the conditional FDR in both directions (i.e., phenotype 1 conditioned on phenotype 2, and vice versa) and selecting the more conservative value. This ensures that the final FDR is robust to directional biases, taking the maximum value from the two computations as the conjunctional FDR. SNPs with conjunctional FDR values below a specified threshold are considered pleiotropic, signifying their involvement in both traits of interest (Andreassen et al. 2013). We used the recommended threshold of 0.05 to identify the pleiotropic SNPs.

To delineate pleiotropic loci, significant SNPs identified through pleioFDR are mapped to independent genomic regions. This involves merging physically overlapping lead SNPs into loci when they are located within linkage disequilibrium (LD) blocks less than 250 kb apart. LD boundaries are determined by identifying all SNPs in LD ($r2 \geq 0.1$) with the significant SNPs in the region, effectively grouping them into a single locus. The most statistically significant SNP within the locus is then selected as the lead SNP, representing the locus. The genomic structure of these loci is determined using the LD patterns derived from European populations in the 1000 Genomes Project (1KGP) as a reference. This ensures that the identified loci accurately reflect population-specific genetic architecture.

Certain genomic regions known for structural complexity or susceptibility to rearrangements are excluded to prevent spurious associations. These include the extended major histocompatibility complex (MHC) region (chr6: 25,119,106–33,854,733, hg19) and the 8p23.1 region (chr8: 7,200,000–12,500,000). These regions often exhibit high levels of LD and structural variation, complicating the interpretation of genetic associations (Smeland et al. 2020).

The pleioFDR method provides several advantages over traditional GWAS approaches. By leveraging pleiotropy, it improves statistical power to detect loci shared across traits, even when individual trait associations might not reach genome-wide significance. Additionally, the use of conjunctional FDR ensures a conservative and robust identification of pleiotropic SNPs, minimizing false positives while highlighting biologically meaningful overlaps. This makes pleioFDR an invaluable tool for uncovering the shared genetic underpinnings of complex traits, contributing to a deeper understanding of their etiology and potential therapeutic targets (Andreassen et al. 2013).

We applied conjFDR for every pair of disease-fitness components using the common set of SNPs between all the studied traits ($N_{SNPS}$ = 2,241,316) which included the 62 complex diseases, longevity and fertility. For all identified pleiotropic SNPs (conjFDR $< 0.05$), we identified the disease risk alleles and classified them based on their effects on the fitness components.

Positive pleiotropy was defined as cases where the disease risk allele was associated with increased longevity (that match antagonistic pleiotropy expectations) or fertility. Conversely, negative pleiotropy was characterized by disease risk alleles associated with reduced longevity or fertility. Please refer to the glossary on the main text for further details.

#### 2.5. Functional annotation

We utilized the Ensembl Variant Effect Predictor (VEP, http://grch37.ensembl.org/Tools/VEP) release 113 (October 2024) with the GRCh37 (hg19) assembly as the reference genome to perform gene-based annotation and identify the functional consequences of pleiotropic SNPs. Pleiotropic SNPs (conjFDR < 0.05) were mapped to genes by physical proximity, involving a 5 kilobase symmetric window around the transcription start and stop sites for each gene to capture variants in regulatory regions.

For each variant mapped to the reference genome, VEP identifies all overlapping Ensembl transcripts and employs a rule-based approach to predict the effects of each allele on each transcript (McLaren et al. 2016; Harrison et al. 2024). The potential consequences are defined using Sequence Ontology terms, with each allele potentially having distinct effects across different transcripts (Eilbeck et al. 2005).

Then, we performed a functional enrichment analysis using common SNPs from the conjFDR analysis as the background gene set. The significance of pathway enrichment was determined using a hypergeometric test with a p-value threshold of 0.05. We employed the FUMA GENE2FUNC (Watanabe et al. 2017) tool to systematically identify differentially expressed genes, as well as to determine relevant tissues and associated diseases using data from the GTEx v.8 (August 2019) on 30 general and 54 specific tissue types (THE GTEX CONSORTIUM 2020). Gene data were obtained from the NCBI Gene database (https://www.ncbi.nlm.nih.gov/gene/), maintained by the National Library of Medicine (NLM) at the National Institutes of Health (NIH). This resource integrates gene-specific information from multiple sources, including RefSeq genomes, curated annotations, and automated data from various databases and consortia.

#### 2.6. Evolutionary analysis of pleiotropic SNPs

Positive selection, in the context of evolutionary biology, refers to the process by which advantageous traits or genetic variations that enhance an organism's fitness are favored and become more prevalent in a population over time (Maynard Smith 1998). This selection mechanism promotes the survival and reproduction of individuals carrying these beneficial traits, leading to their increased frequency in subsequent generations. Positive selection plays a crucial role in driving evolutionary changes and adaptations in response to environmental pressures or selective forces (Nielsen 2005).

We conducted an analysis on signatures of natural selection in the human genome using pre-computed scores that allow the detection of positive selection over human evolutionary history. These are 1) integrated haplotype score (iHS) (Voight et al. 2006) and 2) singleton density score (SDS) (Field et al. 2016).

The iHS is based on the extended haplotype homozygosity (EHH) statistic, which measures the homozygosity of haplotypes surrounding a particular allele (Voight et al. 2006). The iHS is calculated by comparing the EHH of the ancestral and derived alleles at a given SNP and integrating the EHH curves to obtain a single value for each allele. The iHS is then calculated as the log ratio of the integrated EHH values for the ancestral and derived alleles.

The SDS is a statistical method used to detect very recent positive selection in genomic data. It is based on the idea that recent selection can be observed in a sample's genealogy as differences in the branch length distribution (Field et al. 2016). It assumes that derived alleles increasing in frequency have shorter branches and ancestral alleles decreasing in frequency have longer branches, which should result in fewer mutations in the derived alleles (Field et al. 2016). The SDS method calculates the distance to the nearest singleton upstream and downstream from each SNP. It then uses the distributions of distances for each of the three genotypes at the test SNP to compute a maximum likelihood estimate of the log ratio of mean tip-branch lengths for the derived vs. ancestral alleles.

Both iHS and SDS provide an estimate per SNP that is the result of a ratio between the ancestral and derived states, where the two alleles serve as an internal control for each other. We took advantage of this configuration to specifically target the disease-risk allele. Therefore, for this analysis, positive pleiotropies refer to the disease-risk allele, which is also associated with increased fertility and longevity. In contrast, in negative pleiotropies, the disease-risk allele is associated with decreased fertility and longevity. In this way, the difference between positive and negative pleiotropies is due to their opposite effects on fertility and longevity.

First, we needed to match the disease-risk and protective alleles with their corresponding ancestral or derived states. For SDS, we used the same ancestral and derived state definition that is provided in the pre-computed resource (Voight et al. 2006). And for iHS, we used pre-computed values from the PopHumanScan online catalog (Murga-Moreno et al. 2019). Then, we needed to adjust the ratio so that the denominator corresponds to the disease-risk allele. When the disease-risk allele was already the denominator, we left the ratio untouched. But if the disease-risk allele was the nominator, then we changed signs. Due to this modification, we show the iHS and SDS values of the disease-risk alleles at each position.

We noted that approximately 25% of pleiotropic SNPs recurred across multiple diseases, causing the same variant to appear repeatedly among positive and negative pleiotropies. To avoid duplicative counting in the evolutionary analysis, we retained only the instance tied to the earliest-onset disease, capturing the variant at its first phenotypic effect. If a variant was classified as both positive and negative, we kept its earliest occurrence in each category. Finally, to ensure our approach did not introduce bias, we reanalyzed the data after removing duplicated pleiotropic variants with potential ambiguous effects and conducted a leave-one-out cross-validation, excluding one disease at a time.

To investigate differences between positive and negative pleiotropies that exhibited significant variation on positive selection indices, we performed a resampling-based analysis. This approach assessed the stability of observed differences and evaluated statistical significance through non-parametric testing. Specifically, we conducted 10,000 rounds of resampling, each time drawing random samples with replacement from both pleiotropy groups while preserving their size. For each iteration, we computed mean values in each group, applied a Mann-Whitney U test, and recorded the resulting p-values. We then determined the proportion of iterations yielding p-value $< 0.05$ to gauge result robustness. For additional context, we compared these outcomes with the observed means and p-values calculated from the original dataset.

#### 2.7. Mendelian randomization

Mendelian randomization leverages natural genetic variation to mimic randomized controlled trials, offering a powerful approach to establish causal relationships between traits (Davey Smith and Hemani 2014). By using genetic variants as instrumental variables (IVs), MR mitigates confounding biases inherent in observational studies, allowing for robust inference about the direction and strength of causal effects.

**Supplementary Figure 33** displays a flowchart with step-by-step key concepts and details.

In this study, we employed a two-sample MR framework, utilizing GWAS summary statistics to assess the relationships between longevity, fertility, and 62 complex diseases. The use of large-scale GWAS data enhances the statistical power and generalizability of MR findings. However, sample overlap, particularly involving datasets derived from the UK Biobank, requires careful interpretation of the results (**Supplementary Table 14**). Sensitivity analyses, including MR-Egger regression (Bowden, Davey Smith, and Burgess 2015) and MR-PRESSO (Verbanck et al. 2018), were implemented to ensure the robustness of causal estimates while accounting for potential pleiotropic effects.

Multivariable MR (MVMR) extends the standard MR framework by incorporating additional exposures into the analysis. This approach is particularly valuable when examining traits influenced by mediating factors, such as socioeconomic variables. MVMR allows for the simultaneous evaluation of multiple causal pathways, estimating the direct effect of each exposure on the outcome while adjusting for shared genetic influences (Hemani, Bowden, and Davey Smith 2018).

In this study, MVMR was used to account for potential confounding by socioeconomic factors, such as educational attainment, in the relationships between life-history traits and complex diseases. By addressing these additional pathways, MVMR enhances the precision and validity of causal estimates, providing deeper insights into the biological and environmental mechanisms underlying these associations. Although this approach does not fully account for the influence of socioeconomic factors on the fertility-disease and longevity-disease relationships, it does allow for the assessment of potential causal relationships between these socioeconomic factors and life-history traits.

We selected genome-wide significant SNPs ($p < 5x10^{-8}$) from each exposure GWAS as IVs, clumping them using a linkage disequilibrium (LD) threshold of ($r^2 < 0.1$) within a 250 kb window. Analyses were not conducted for traits with fewer than two IVs, ensuring the reliability of the results.

The primary analysis was conducted using the fixed-effects inverse variance weighted (IVW) method, which assumes no horizontal pleiotropy and offers the highest statistical power when MR assumptions are met (Hemani, Bowden, and Davey Smith 2018). Sensitivity analyses, including MR-Egger regression, Cochran's Q statistic, and MR-PRESSO, were performed to test for horizontal pleiotropy and heterogeneity. MR-Egger regression accounts for directional pleiotropy by allowing a non-zero intercept (Bowden, Davey Smith, and Burgess 2015), while MR-PRESSO identifies and corrects pleiotropic outliers by removing SNPs contributing excessive heterogeneity (Verbanck et al. 2018).

For early-onset diseases, bi-directional MR analyses assessed causal relationships in both directions between fertility and disease risk. For late-onset diseases and longevity, unidirectional analyses were employed to align with the temporal progression of these traits. This design reflects the biological plausibility of these relationships, as late-onset diseases are unlikely to influence fertility directly.

Given the limited statistical power of the MR analyses, we refrain from making definitive conclusions about the causality of the observed relationships. Instead, our focus is on interpreting the point estimates and their directions of effect in the context of the genetic correlation and pleiotropy results.

#### 2.8. Demographic analysis

To clarify the role of disease status in the observed positive genetic correlation between fertility and complex disease risk, we analyzed fertility outcomes in the UK Biobank using disease-specific polygenic scores (PGS).

##### UK Biobank genetic data and QC

The UK Biobank (UKBB) is a large, population-based cohort that contains genetic and phenotypic data for over 500,000 participants (Sudlow et al. 2015). We obtained the version 3 (2018) release of the imputed UKBB genetic data and conducted standard quality control (QC) procedures with PLINK v1.9 (Purcell et al. 2007). At the variant level, we retained autosomal variants with a minor allele frequency (MAF) $> 0.01$, a call rate $> 0.98$, and Hardy–Weinberg equilibrium p-value $\geq 10^{-6}$. At the individual level, we included only self-identified White British participants, excluded those with missingness $> 0.02$, first- or second-degree relatedness, or a heterozygosity rate exceeding three standard deviations from the mean. Finally, we performed principal component analysis (PCA) on the final set of individuals to adjust for population structure (Collister, Liu, and Clifton 2022). The latest update to remove withdrawn participants was done in January 2025.

##### Polygenic scores for complex diseases

PGS were generated for each disease using PRScs (https://github.com/getian107/PRScs), a Bayesian method that applies continuous shrinkage priors to SNP effect sizes (Ge et al. 2019). In this approach, the degree of shrinkage for each SNP depends on its GWAS association signal, while local linkage disequilibrium patterns are modeled jointly. We then used PLINK v1.9 (Purcell et al. 2007) to calculate individual-level PGS for all UK Biobank participants.

To account for technical and population-structure artifacts, we regressed each PGS on sex, age, sample missingness, genotyping array type, the first ten principal components (PCs), and two socioeconomic factors—education score (UKBB field 26414) and income score (UKBB field 26411). We subsequently used the regression residuals for all downstream analyses. Participants were then classified as "diagnosed" or "undiagnosed" for each disease, and their PGS values were binned into deciles. Relative odds ratios (ORs) were calculated for each decile using the fifth decile as the reference.

We assessed the predictive performance of each PGS by calculating the variance explained ($R^2$) on the liability scale, incorporating unadjusted period prevalences from the FinRegistry R11 release (S. H. Lee et al. 2012; Choi, Mak, and O'Reilly 2020). All analyses were carried out in R version 3.6.0 (R Core Team 2022).

### 3. Extended Results and Discussion

#### 3.1. Genetic trade-offs between complex diseases and fitness components

##### Longevity

Among the 62 complex diseases we examined, 41 had significant correlations with longevity ($P<0.05$). Among the diseases with nominal significance, 85% (n = 35) exhibited negative genetic correlations with longevity, suggesting that individuals with higher genetic susceptibility to these diseases tend to have shorter lifespans (**Fig. 1a**). Notably, among the subset of diseases meeting a stricter threshold of FDR < 0.05 (n = 38), 87% (n = 33) also showed negative correlations (**Supplementary Fig. 8**). This consistent pattern highlights the importance of understanding how shared genetic factors influence both disease risk and lifespan, capturing the broad impacts that genetic predisposition to various conditions can have on overall survival. Interestingly, only six diseases ($P<0.05$) showed a positive correlation with longevity; and these associations were generally weaker in both significance and effect size compared to those with negative correlations (**Supplementary Table 4**).

The circulatory, endocrine and metabolic, and musculoskeletal systems emerged as the domains most strongly associated with reduced longevity (**Supplementary Fig. 4**), aligning with evidence that these systems critically influence aging and mortality through interconnected biological mechanisms. For instance, age-related cardiovascular diseases remain the leading cause of mortality worldwide (Abdellatif et al. 2023). Similarly, the musculoskeletal system supports longevity by preserving mobility and physical function; the progressive decline in bone and muscle mass is tied to increased frailty and mortality in older adults (Azzolino et al. 2021). The endocrine and metabolic systems are likewise central to lifespan regulation. Notably, most of the hallmarks of aging involves adverse metabolic changes, and the insulin/IGF-1 pathway has demonstrated lifespan-extending effects in model organisms, indicating promising therapeutic avenues for humans (López-Otín et al. 2016; Junnila et al. 2013).

#### Fertility

In contrast, fertility analysis revealed a striking pattern of predominantly positive genetic correlations with disease risk. Among the 62 diseases analyzed 30 were nominally significant ($P<0.05$), and 87% ($n = 29$) showed positive correlations with fertility (**Fig. 1b** and **Supplementary Table 4**). Notably, 92% ($n = 23$) of the most statistically significant associations ($FDR < 0.05$, $n = 25$) also exhibited positive correlations (**Supplementary Fig. 10**). This counterintuitive finding suggests that genetic backgrounds associated with increased disease susceptibility may confer reproductive advantages. Such observations hint at complex pleiotropic interactions between fertility and disease risk that could be relevant to evolutionary fitness.

Attention Deficit Hyperactivity Disorder (ADHD) demonstrated the strongest positive correlation with fertility ($r_g$=0.41, P=1.1e-27). This finding aligns with previous research examining female reproductive behaviors and psychiatric disorders, which reported that increased genetic liability for ADHD is associated with earlier age at first birth and a higher number of births—both indicators of greater lifetime reproductive success (Ni et al. 2019; Barban et al. 2016).

Interestingly, our results also revealed exceptions to this trend. Anorexia nervosa emerged as one of only two diseases negatively associated with fertility in our analysis ($r_g$=-0.0823, P=0.0191). This observation is consistent with prior studies showing that higher genetic liability for eating disorders, including anorexia nervosa, correlates with later age at first sexual intercourse and birth, reflecting reduced fertility (Ni et al. 2019).

Additionally, we found that refractive error, measuring particularly myopia, exhibited a strong negative genetic correlation with fertility ($r_g$=-0.206, P=1.97e-21). This finding is supported by demographic data from the UK Biobank, which demonstrated an association between myopia and both delayed reproduction and fewer offspring (Long and Zhang 2021). However, authors suggested that this relationship might be indirectly mediated by educational attainment (Long and Zhang 2021), highlighting the complex interplay between genetic factors, environmental influences, and reproductive outcomes.

#### Evolutionary frameworks for early- and late-onset diseases

Evolutionary theories of aging predict a trade-off in the fitness impact of mutations based on the timing of their effects during an individual's life. This concept is rooted in the principle of the “selection shadow”, which posits that the strength of natural selection declines with age (Medawar 1952). This decline occurs because natural selection primarily acts during the reproductive window. Mutations with deleterious effects that manifest later in life, after reproduction has occurred, are less likely to be removed from the genetic pool, as they have already been passed onto offspring. Thus, even mutations with negative impacts on fitness may persist if their effects emerge after the reproductive period.

Two prominent evolutionary theories—Mutation Accumulation (MA) and Antagonistic Pleiotropy (AP)—provide complementary frameworks to understand the forces shaping the prevalence of disease-risk alleles for both early- and late-onset diseases (Medawar 1952; G. Williams 1957). The MA theory posits that harmful mutations that manifest later in life experience weaker selective pressure, allowing them to persist in populations. And the AP theory asserts that mutations conferring benefits to fitness early in life may be retained because they outweigh deleterious impacts that emerge later.

Building on these ideas, we hypothesize that the genetic basis of late-onset diseases is more likely to display stronger negative correlations with longevity. Whereas early-onset diseases may show stronger positive correlations with fertility. To test this, we calculated Spearman correlations between genetic correlations ($r_g$) and age at disease onset (ADO), restricting the analysis to diseases with significant associations ($P < 0.05$). We chose Spearman's rank-based method because $r_g$ follows an approximately normal distribution, whereas ADO does not.

Notably, we found that negative genetic correlations with longevity were more pronounced for later-onset diseases (r=–0.43, P=5e-03, **Supplementary Fig. 5**) and leave-one-out cross-validation confirmed that no single disease drove this trend (**Supplementary Table 3**). This result aligns with MA, indicating that deleterious mutations acting at older ages remain under weaker purifying selection and can accumulate in the population, ultimately reducing lifespan. By contrast, there was no significant overall correlation between fertility genetic correlations and disease onset (r=–0.01, P=9.5e-01, **Supplementary Fig. 6**).

These results highlight distinct patterns in the genetic architecture of early- and late-onset diseases. While the AP framework suggests a potential trade-off favoring fitness-enhancing mutations during early life, the MA theory underscores the persistence of deleterious alleles affecting late-life phenotypes, consistent with evolutionary dynamics. Together, they provide a nuanced evolutionary explanation for why disease-risk alleles persist across diverse age windows, shaping life-history traits and disease outcomes.

#### 3.2. Pleiotropic loci shared between fitness components and complex diseases

##### Pleiotropies with longevity

Across all diseases, 1,142 independent pleiotropic SNPs were identified as significantly associated with both disease risk and longevity (conjFDR < 0.05, **Fig. 1c**). While the overall direction of genetic correlations between disease and longevity was consistently negative, individual disease-longevity associations often included pleiotropic loci with opposite directional effects (**Supplementary Table 4**). Of the 1,142 pleiotropic loci identified, 942 (82%) showed negative pleiotropy, where the effects of the alleles on disease risk and longevity were in opposite directions. In contrast, 200 loci (18%) showed positive pleiotropy, with concordant effects on both disease risk and longevity (**Fig. 1c**). Overall, the circulatory system (n = 510) and the endocrine, nutritional and metabolic domain (n = 250) accounted for 67% of the pleiotropies with longevity, clearly enriched in negative pleiotropies (**Supplementary Fig. 11**).

Across the 62 diseases examined, 12 (19%) showed no pleiotropy with longevity. Among those displaying only one type of pleiotropy, 11 (17.7%) exhibited exclusively negative pleiotropies, while 3 (4.8%) manifested exclusively positive pleiotropies. No distinct pattern emerged regarding disease domains or age at disease onset, suggesting that these conditions form a heterogeneous subset of the studied diseases.

Several conditions exhibited notably high pleiotropy counts with longevity. Hypertension (3691) presented 170 pleiotropies with 97.6% being negative, myocardial infarction (GCST011364) had 162 pleiotropies with 98.7% negative, coronary artery disease (3693) had 111 pleiotropies with 97.3% negative, and lipoprotein metabolism disorders (3688) reached 106 pleiotropies with 98.1% negative. This pronounced negative relationship with lifespan was especially evident among circulatory disorders. By contrast, refractive error (GCST010003) showed the greatest number of positive pleiotropies at 26, followed by breast cancer (BCAC2020BC) and schizophrenia (PGC2022SCZ), each with 16. Collectively, these findings emphasize the diversity of pleiotropic profiles across diseases and illuminate the intricate genetic pathways of health span.

Notably, 25% (n = 191) of the 1,142 SNPs exhibited pleiotropy across multiple diseases (**Supplementary Table 4**). For additional details, please refer to Supplementary Section 3.2.3. For the functional analysis, we did not focus on disease-specific pleiotropies. Rather, we grouped the pleiotropic SNPs as either positively or negatively associated with longevity. Hence, even if a single SNP was linked to several diseases, we only counted it once to avoid redundancy. Therefore, we did the functional analysis with the set of 750 unique pleiotropic SNPs between longevity and disease (**Supplementary Tables 15-16**).

These loci were predominantly located in intronic regions (40.3%) and downstream (16.9%) and upstream (15%) of genes (**Supplementary Fig. 12**). Most variants were mapped to protein coding genes (64.6%) (**Supplementary Fig. 13**). The observed proportions were consistent with those in the background set of 2.2 million common SNPs shared across all analyzed traits, where the same functional categories and biotypes were also predominant (**Supplementary Fig. 32**). Moreover, these findings align with previous research on pleiotropic variants, which identified a preponderance of such variants in intronic regions and in proximity to genes (Watanabe et al. 2019). This distribution suggests that pleiotropic variants may function as regulatory elements, influencing gene expression and contributing to the observed pleiotropy, thereby reinforcing the importance of non-coding regions in mediating pleiotropic effects across multiple traits.

Chromosomes 1, 6, and 19 harbored the largest number of pleiotropic signals with longevity. Although 70–80% of these pleiotropies were negative, these chromosomes also contained the highest counts of positive pleiotropies. Notably, chromosome 2 displayed a relatively higher proportion of positive pleiotropies, accounting for 32% (**Supplementary Fig. 14**).

We conducted separate over-representation analyses for negative and positive pleiotropies with longevity, mapping SNPs to genes using the Ensembl Variant Effect Predictor (VEP). The primary aim was to characterize the distinct biological and molecular functions mediating how each variant or gene influences both longevity and disease risk.

To investigate tissue-specific expression, we used a differential expression analysis of these genes against GTEx data implemented in FUMA. Negative pleiotropies with longevity were upregulated in the colon when considering 30 general tissue classes (**Supplementary Fig. 15**). At a more granular level of 54 specific tissues, negative pleiotropies were downregulated in the cervical spinal cord and the substantia nigra of the brain (**Supplementary Fig. 16**). Research has demonstrated that genetic dysregulation in the colon and substantia nigra contributes to a range of disorders. In the colon, altered gene expression can predispose to conditions such as colon cancer and ulcerative colitis (Birkenkamp-Demtroder et al. 2005; Noble et al. 2008), whereas in the substantia nigra, deregulatory processes are implicated in schizophrenia, Parkinson's disease, and Alzheimer's Disease (M. R. Williams et al. 2014; Simunovic et al. 2009; López González et al. 2016).

Additionally, FUMA's gene set over-representation analysis (FDR < 0.05) highlighted several cytogenetic loci from the MsigDB c1 database with a high proportion of genes implicated in disease risk and associated with reduced longevity (**Supplementary Table 5**):

1. Within chr1p13, genes such as PRSC1 modulate LDL and other blood lipids, known risk factors for cardiovascular diseases (Keebler et al. 2010). PTPN22, another gene within this region, appears pleiotropic for cardiovascular disease and rheumatoid arthritis (Guo et al. 2023).

2. In the chr6p22 region, many genes lie near the human leukocyte antigen (HLA) system (Shiina et al. 2009). These loci are associated with cerebrospinal fluid regulation and

candidate genes for Alzheimer's disease, as well as immune-related pathways involving leukocyte-mediated immunity and antigen processing (Western et al. 2024).

3. The short arm of chromosome 7 at band 15 (chr7p15) encompasses the glycoprotein non-metastatic melanoma protein B (GPNMB) locus, which has a causal link to Parkinson's disease (Murthy et al. 2017). This region also contains INHBA, implicated in gastric cancer (Q. Wang et al. 2012), and members of the HOX gene family, essential to developmental processes. The HOXA locus exhibits prominent hypermethylation in glioblastoma tumors, which correlates with increased resistance to chemo-radiotherapy (Kurscheid et al. 2015).

4. On chromosome 6p25, risk elevations have been reported for all-stroke subtypes, particularly ischemic, non-cardioembolic, and cardioembolic stroke. The lead variant and those in strong linkage disequilibrium map either to the FOXQ1 or FOXF2 genes (Chauhan et al. 2016).

5. Farther along, the nicotinic receptor cluster at chr15p25, including CHRNA3 and CHRNA5, associates with nicotine dependence, smoking intensity, and elevated lung cancer risk (Caporaso et al. 2009).

6. Finally, the chr17p21 region harbors BRCA1, a seminal breast and ovarian cancer susceptibility gene central to genomic integrity and DNA repair (Hall et al. 1990; Narod and Foulkes 2004).

Pathway enrichment analysis using WikiPathways annotations in FUMA revealed 14 enriched pathways (**Supplementary Table 5**), six of which were directly related to lipoprotein metabolism. Key pathways included cholesterol metabolism (FDR=4.17e-04), processes involving LDL and HDL in cholesterol transport (FDR=1.30e-03), and the role of statins as competitive inhibitors in endogenous cholesterol synthesis (FDR=1.30e-03). Additional pathways of interest were associated with cardiomyocyte hypertrophy (FDR=2.97e-03), neuroblastoma metastasis (FDR=2.08e-02), the Type I interferon cytokine pathway (FDR=3.63e-02), and the insulin signaling pathway (FDR=4.46e-02), which is essential for glucose uptake from the blood.

In contrast to the genes exhibiting negative pleiotropy, our analysis of genes with positive pleiotropy did not reveal any significant tissue-specific differential expression. This lack of tissue specificity may be attributed to the relative scarcity of positive pleiotropies associated with longevity and our smaller sample size for this category. Consequently, the pathway enrichment analysis for positive pleiotropic genes yielded less informative results compared to those obtained for negative pleiotropic genes. Nonetheless, enrichment of the chr6p22 cytogenetic location and multiple cellular and molecular functions related to chromatin structure was observed, attributable to the large number of histone genes within the positively pleiotropic set (**Supplementary Tables 5**). A major histone gene cluster has been identified at position chr6p22 that might be implicated in chromatin structural alterations that affect gene expression in breast cancer (Fritz et al. 2018). Of the seven enriched GO cellular components, five were related to histones: DNA packing complex, nucleosome, chromosome, protein-DNA complex, and chromatin. Each of these terms encompasses structures composed of DNA and associated proteins that organize, package, and regulate genetic material within the cell. Chromatin structure specifically plays a critical role in regulating gene expression and in organizing chromosomes. It consists of DNA wrapped around histone proteins to form nucleosomes, which can be further compacted into higher-order configurations. These chromatin states enable dynamic control of DNA accessibility and transcriptional activity (Li, Carey, and Workman 2007; Bannister and Kouzarides 2011).

Overall, our results illustrated contrasting mechanisms by which pleiotropic genes influenced longevity and disease risk. Negative pleiotropies with longevity appear to act through diverse sets of genes that increase disease vulnerability and ultimately reduce lifespan. In contrast, positively pleiotropic genes were strongly enriched for histone genes and chromatin structural components. This enrichment suggested a potential trade-off, wherein elevated disease risk could coincide with enhanced longevity through mechanisms involving chromatin remodeling and gene regulation, highlighting the complex interplay between genetic risk factors and lifespan.

#### Pleiotropies with fertility

We identified 460 independent pleiotropic SNPs (conjFDR $< 0.05$) jointly associated with complex disease and fertility (**Fig. 1d**, **Supplementary Table 6**). Of these, 229 loci (49.8%) exhibited positive pleiotropy, whereas 231 loci (50.2%) showed negative pleiotropy.

Overall, no pleiotropy with fertility was observed for nine diseases (**Fig. 1d**). Among the remaining 53 diseases, 48 exhibited at least one positive pleiotropic SNP with fertility, and 40 had at least one negative pleiotropic SNP with fertility. Fourteen diseases showed no positive pleiotropies with fertility, while 22 showed no negative pleiotropies. Five diseases manifested only negative pleiotropies with fertility, and thirteen displayed only positive pleiotropies. Both groups constituted a heterogeneous set of diseases with no evident enrichment in a particular disease domain or onset age category.

Regarding individual diseases, refractive error (GCST010003) exhibited the highest number of pleiotropies (n = 54), of which 81% were negative. This disease also displayed the greatest number of negative pleiotropies with fertility, followed by alcohol use (PGC2019ALCUSE), which had 18 (**Fig. 1d**). The next two diseases with the largest number of pleiotropies were schizophrenia (PGC2022SCZ) and insomnia (4290), each with 29; for schizophrenia, 48% were positive, whereas 76% were positive for insomnia. Furthermore, insomnia showed the highest number of positive pleiotropies with fertility, followed by Major Depressive Disorder (PGC2019MDD) with 15. These findings highlight the heterogeneous nature of fertility-associated pleiotropies across diseases and underscore the positive genetic relationship between certain mental disorders and increased fertility.

Notably, 26% (n = 74) of the 460 SNPs exhibited pleiotropy across multiple diseases (**Supplementary Table 6**; see next section for details). As with the longevity analysis, we grouped pleiotropic SNPs by their association with fertility—either positive or negative. To avoid redundancy, each SNP was counted only once, even if linked to several diseases. Consequently, the functional analysis was performed on a set of 285 unique pleiotropic SNPs connecting fertility and disease (**Supplementary Table 17**).

Chromosomes 3, 6, 11, and 17 displayed the highest number of fertility-associated pleiotropic signals, with chromosome 6 harboring the largest proportion (25%). Meanwhile, chromosomes 10, 13, 14, 15, and 18 showed at least 67% positive pleiotropies, whereas chromosomes 9, 16, and 19 displayed 60% or more negative pleiotropies (**Supplementary Fig. 34**). Over-representation analysis of cytogenetic locations from the MsigDB c1 database revealed an enrichment of genes in the chr3p21 region (FDR=5.27e-03) for positive pleiotropies and in chr17q12 (FDR=4.86e-04) and chr2q34 (FDR=1.12e-02) for negative pleiotropies (**Supplementary Table 17**).

Notably, the chr3p21 region includes a well-documented susceptibility locus for severe COVID-19 inherited from Neanderthals (The COVID-19 Host Genetics Initiative 2020; Zeberg and Pääbo 2020). Our results also suggest that pleiotropic variants in this region may enhance reproductive outcomes while increasing disease risk, particularly in the circulatory, mental, and digestive domains (**Supplementary Table 6**). Although few studies link chr3p21 to reproductive outcomes, one report identified a copy-neutral loss of heterozygosity (cnLOH) at 3p21.31 that may be associated with male infertility (Singh et al. 2019). The significance of these combined findings remains unclear, and we did not investigate this further, leaving it an open question for future research. Lastly, we did not identify any additional enriched biological pathways for positive pleiotropies in the FUMA databases.

For negative pleiotropies, the chr17q12 region was associated with a copy number variation syndrome (FDR=2.36e-03, **Supplementary Table 17**). One study identified a deletion within this locus that increases the risk of autism and schizophrenia (Rasmussen et al. 2016), and a strong candidate locus for asthma also resides in this region (Stein et al. 2018). Additionally, genes within the chr17q11 amplicon are affected by copy number alterations in breast tumors (FDR=1.14e-03). Further mapping indicated that negative pleiotropies involve WNT4, ESR1, and MED1, which were enriched in a GO biological process (MsigDB c5) related to mammary gland duct branching during pregnancy (FDR=1.06e-02, **Supplementary Table 17**). These genes form part of an estrogen-responsive cascade that expands the ductal system in the first trimester (Alex, Bhandary, and McGuire 2020) and are also implicated in decidualization of the endometrium, creating a favorable environment for placenta formation (Matsuyama, Whiteside, and Li 2024). Our results suggest that pleiotropic variants regulating these genes could enhance reproductive success while simultaneously offering protection against certain diseases.

Overall, these findings emphasize the complex interplay between fertility and disease risk, highlighting the presence of both positive (concordant effects) and negative (opposed effects) genetic trade-offs. The enrichment of mental and digestive diseases, among others, underscores the diverse ways in which pleiotropy can shape health and reproductive outcomes. Further research with functional studies of specific loci will be essential to clarify the biological underpinnings of these pleiotropic effects and to determine how they might be leveraged for interventions aimed at improving both reproductive and general health.

#### Pleiotropic variants across multiple disease phenotypes

Approximately one quarter of the pleiotropic variants linking life-history traits to diseases exhibited pleiotropy for two or more diseases (**Supplementary Tables 4** and **6**). For both longevity and fertility, the maximum number of diseases associated with a single variant was 12. Here, we focused on the variants displaying high pleiotropic potential as they may inform an integrated approach spanning geroscience and reproductive health.

For longevity, rs17696736 showed pleiotropy with 12 diseases, and in 9 of these it acted as a negative pleiotropy—indicating that increased disease risk was associated with reduced longevity. This intronic variant maps to the N-alpha-acetyltransferase 25, NatB auxiliary subunit (NAA25) gene, which catalyzes a major post-translational modification by acetylating methionine residues followed by acidic or asparagine residues. NatB is essential for normal cell proliferation (Starheim et al. 2008), and NAA25 knockdown studies have demonstrated reduced breast cancer progression via increased apoptosis (Xu et al. 2022). Moreover, NatB is involved in influenza virus shutoff activity, suggesting a broader role in virus replication (Oishi et al. 2018).

Another highly pleiotropic variant associated with longevity was rs2476601, a missense variant in the lymphoid-specific intracellular phosphatase gene PTPN22. This gene is strongly linked to type 1 diabetes (T1D) (Bottini et al. 2004) and multiple autoimmune diseases such as rheumatoid arthritis (RA) (Begovich et al. 2004). rs2476601 showed genome-wide significant associations in T1D and RA GWASs, and our pleiotropy analysis extended its disease associations to additional disease domains as well as longevity.

For fertility, the variant rs2071293 exhibited pleiotropy with 12 different diseases. Notably, in 8 of these cases, the pleiotropic effect was negative, meaning that the allele conferring protection against these diseases was simultaneously associated with higher fertility. Of the remaining diseases, Crohn's Disease, Ulcerative Colitis, and Alzheimer's Disease are autoimmune-related or involve dysregulated immune responses. When queried in Ensembl, rs2071293 was found to map to multiple genomic locations, reflecting the structural complexity of the surrounding region. More specifically, this SNP lies in an intergenic segment within the major histocompatibility complex (MHC)—recognized as the most variable region in the human genome. Due to this complexity, the same SNP appears in four characterized MHC haplotypes in Europeans (COX, DBB, MANN, and MCF) (Horton et al. 2008). According to the 1000 Genomes Project Phase 3 data (European population), the fertility-increasing allele occurs at a frequency of approximately 67% and corresponds to the ancestral state. The interplay among the pleiotropic effects suggests a potential evolutionary trade-off that highlights both the genetic intricacy of the MHC and the multifaceted nature of fertility-related processes.

The second most pleiotropic variant linking complex diseases and fertility was rs3130976, which lies upstream of PSORS1C1 and C6orf15 and downstream of CDSN, all within the MHC region. Notably, PSORS1C1 has been implicated in several immune-related conditions, including systemic sclerosis and psoriasis (Allanore et al. 2011; Wiśniewski et al. 2018). Although no direct research has connected these specific genes to fertility outcomes, a study in the Hutterite population showed that HLA genes can influence both pregnancy outcomes and mate choice (Ober 1999), although the choice of the sample population may bias the results (Havlicek and Roberts 2009).

In conclusion, these pleiotropic variants may function as pivotal nodes in an omnigenic network linking disease with key life-history traits, highlighting the need for deeper investigation into their broader biological and clinical implications.

#### Three pleiotropic SNPs linking disease risk, longevity and fertility

We conducted an independent search for pleiotropic variants associated with disease, focusing separately on longevity and fertility. Among the identified variants, three stood out for their pleiotropic relationships, linking one or more diseases to both longevity and fertility. While this finding is not unexpected given the polygenic (or omnigenic) architecture of the studied phenotypes (Boyle, Li, and Pritchard 2017), it underscores potential genetic and evolutionary trade-offs between life-history traits and health outcomes.

The first variant, rs3811696, is located upstream of the GMPPB gene and downstream of the IP6K1 gene. This variant demonstrates antagonistic pleiotropy between heart failure, longevity, and fertility. Specifically, the allele associated with increased fertility correlates with a higher risk of heart failure while simultaneously being linked to reduced longevity. This finding suggests an intricate balance between reproductive fitness and long-term health outcomes.

In contrast, rs4458695 presents a more complex pleiotropic relationship. This variant influence fertility alongside two health conditions: diverticular disease and refractive error. It also exhibited a positive pleiotropic effect with longevity, linking increased risk of alcohol use to higher longevity. Remarkably, while the allele associated with increased fertility heightens the risk of diverticular disease, it appears to offer protection against refractive error and is linked to greater longevity. This duality illustrates an intriguing aspect of pleiotropy where the same genetic variant can confer both advantageous and disadvantageous effects depending on the context and thus their impact on fitness is challenging to determine.

The third variant, rs73077174, is an intronic variant of the IP6K1 gene that showcases a multifaceted pleiotropic landscape. It affects fertility in relation to osteoarthritis and insomnia while also influencing longevity in conjunction with hypertension and myocardial infarction. Like rs3811696, the allele associated with increased fertility in this case was correlated with an increased risk of diseases and reduced longevity. Interestingly, the IP6K1 gene encodes inositol hexakisphosphate kinase 1, a crucial enzyme in the inositol phosphate signaling pathway implicated in a wide range of biochemical processes in mammals. This versatile kinase plays pivotal roles in several fundamental cellular functions—from cytoskeletal remodeling to DNA repair—thereby supporting overall cellular health and function.

The pleiotropic landscape drawn by the circumstance of these three variants, encompassed eight diseases with diverse onset patterns and health impacts. Of these eight conditions, five demonstrated a late-onset profile, with 5% prevalence occurring after age 35. This timing is particularly significant from an evolutionary perspective, as it allows potentially detrimental alleles to persist in the population by manifesting their negative effects primarily after peaking reproductive years.

The Global Burden of Disease Study 2021 for Western Europe provides further context for the remaining three early-onset conditions: insomnia, refractive error, and alcohol use. Despite their earlier manifestation, these conditions contribute substantially less to the overall burden of disability-adjusted life years (DALYs) compared to cardiovascular diseases (CVDs), which accounted for 27.8% of total deaths in Western Europe and remained the leading cause of mortality worldwide (Mubarik et al. 2024).

The observed antagonistic genetic effects point to a broader evolutionary theme: alleles that enhance fertility may persist despite their association with increased disease risk later in life. This pattern aligns with the concept of Antagonistic Pleiotropy, where genes beneficial for reproduction and early survival may have deleterious effects in post-reproductive years. The late onset of the more severe conditions supports this hypothesis, as their detrimental effects emerge after individuals have likely already reproduced.

#### 3.3. Genetic trade-offs between longevity and fertility

We observed a negative genetic correlation between the two fitness components, longevity and fertility ($r_g$=-0.0963, P=0.0024). We also identified 8 pleiotropic loci jointly associated with both traits (at a conjFDR < 0.05, **Supplementary Table 7**). Most of the pleiotropies (7 out of 8) had antagonistic effects, where the allele increasing fertility was associated with reduced longevity, as predicted by the AP theory of aging (G. Williams 1957). According to this theory, alleles offering benefits earlier in life, when selection pressure is high, can persist in the population despite exerting deleterious effects in later life. Specifically, antagonistic pleiotropy arises when an allele enhances fitness (e.g., through reproductive success and survival) yet also contributes to aging at older ages.

Our analysis revealed that antagonistic pleiotropies predominantly manifest as intronic variants localized to protein-coding genes and lincRNAs (**Supplementary Table 7**). The implicated protein-coding genes span diverse functional roles, but none have been previously documented to influence fertility outcomes in humans. For instance, TRAIP is an E3 ubiquitin ligase that is a master regulator during the repair of DNA interstrand crosslinks (Wu et al. 2019) and has been implicated in brain sexual differentiation by regulating apoptosis (Krishnan et al. 2009). The Sphingosine-dependent kinase 1 (SDK1) is a member of the well conserved sphingolipid metabolic network, central modulators of cell growth, migration or senescence (Teixeira and Costa 2016). While sphingolipids serve as essential structural components of cellular membranes, they also function as bioactive signaling molecules. Mechanistically, the Target of Rapamycin (TOR) pathway has been shown to coordinate sphingolipid biosynthesis (Teixeira and Costa 2016).

Although the identified lincRNAs are expressed at low overall levels (TPM < 10), they exhibit marked tissue-specific patterns, with pronounced enrichment in testicular and brain tissues (**Supplementary Fig. 20**). In contrast to the antagonistic variants, the sole agonistic pleiotropy identified corresponds to an intronic variant within the WNT3 gene, a critical regulator of embryonic development and germ cell maturation (Basu et al. 2018). Strikingly, allele frequency analysis from the 1000 Genomes Project Phase 3 in Europeans revealed that the allele associated with increased fertility is rare (0.049).

When examining allele frequencies for all pleiotropic variants, we did not observe the expected trend of higher frequencies for fertility-enhancing alleles. Variants in TRAP, MON1A, and

SNX29 exhibited intermediate allele frequencies, while the fertility-enhancing allele for the lincRNA LINC00669 was very rare (0.019). In contrast, variants in SDK1 and the lincRNA RP11-436D23.1 met expectations, with fertility-enhancing allele frequencies of 0.76 and 0.61, respectively. This complex, incomplete picture underscores the intricate nature of these traits and suggests that either additional fitness-related phenotypes beyond fertility and longevity are influencing variant distributions or that evolutionary pressures have shifted over time.

#### 3.4. Evolutionary signals of pleiotropic SNPs

To examine evolutionary signatures in pleiotropic disease-risk alleles that influence longevity and fertility, we used two established indices of positive selection: the integrated haplotype score (iHS), which detects selection events over the past ~30,000 years (Voight et al. 2006), and the singleton density score (SDS), sensitive to more recent (~2,000 years) polygenic selection (Field et al. 2016). Although both indices ultimately compare derived and ancestral states at biallelic SNPs, we focused on the disease-risk allele to capture its specific contribution to fitness-related traits (**Supplementary Tables 4-6**).

We hypothesized that alleles conferring fitness benefits—enhanced longevity or fertility—would exhibit stronger positive selection than those imposing fitness costs (i.e., reduced longevity or fertility). In positive pleiotropies, the disease-risk allele also boosts longevity or fertility, whereas in negative pleiotropies, it diminishes these traits. By tracking the disease-risk allele itself, we could determine whether any observed selective differences stemmed from its influence on these life-history traits.

We visualized pleiotropic effects across the age at disease onset by plotting cumulative pleiotropies, distinguishing positive (blue) from negative (red) variants. At each age threshold, we calculated the mean effect of pleiotropies occurring before that age. This approach enabled us to pinpoint potential differences between early and late-onset diseases. We employed the Mann–Whitney U test at each age bin, and the final bin incorporated all pleiotropic variants—thus capturing the overall difference between positive and negative pleiotropies (**Fig. 2a**).

Using the SDS, we found a significant divergence between positive and negative pleiotropies associated with fertility (Mann–Whitney P=5.4e-04) (**Fig. 2**). Positive pleiotropies had more positive SDS values (mean = 0.23) than negative ones (mean = -0.32). Because we specifically targeted disease-risk alleles that increase fertility in positive pleiotropies and decrease it in negative ones, these higher SDS values suggest that alleles increasing reproductive success have been favored by natural selection—even at the cost of higher disease susceptibility. Notably, this difference remained apparent for both early- and late-onset diseases when removing all repeated pleiotropic SNPs (**Supplementary Table 6**), and a leave-one-out analysis confirmed that no single disease disproportionately drove the effect (**Supplementary Table 15**). A resampling approach further supported the robustness of these findings, placing the observed difference within the most extreme 5% of the resampled distribution (**Supplementary Fig. 21**).

The SDS was developed using whole-genome sequences from 3,781 individuals in the UK10K Project, a British population cohort. Hence, SDS is optimized to resolve selection signals within the genetic architecture of this population. This methodological alignment is critical for our analysis, as the UK Biobank represents the primary source of GWAS data for both life-history traits and diseases in this study (**Supplementary Table 14**).

In contrast, the iHS, which detects older selection signals (~30,000 years), showed no significant distinction (P=2.4e-01) between positive and negative pleiotropies for fertility, even though positive pleiotropies tended toward more positive values (**Supplementary Table 6)**. The discrepancy between the two measures suggests that the selective forces favoring fertility-increasing alleles have likely acted more recently.

For longevity, no significant difference emerged between positive and negative pleiotropies using SDS (P=5.07e-02) or iHS (P=1.07e-01) (**Supplementary Table 4**). Moreover, these patterns were strongly influenced by the diseases included in the analysis, as indicated by leave-one-out cross-validation (**Supplementary Table 15**).

#### 3.5. Mendelian randomization

Understanding causality is crucial when investigating genetic correlations and pleiotropy between complex traits. While genetic correlations indicate shared genetic architecture, they do not necessarily imply a causal relationship. Pleiotropy, where a single genetic variant influences multiple traits, is common in the human genome and plays a critical role in shaping complex traits (Watanabe 2019). However, distinguishing between causal and non-causal pleiotropy is vital for uncovering the underlying mechanisms driving the observed relationships.

Pleiotropy can be categorized into vertical, horizontal, and spurious types, each with distinct implications for Mendelian randomization (MR). Vertical pleiotropy occurs when one trait causally influences another in a cascade of events, aligning with MR assumptions (Hemani, Bowden, and Davey Smith 2018). In contrast, horizontal pleiotropy arises when a genetic variant independently affects multiple traits through separate pathways, potentially biasing MR results. Spurious pleiotropy, which reflects confounding rather than true biological links, underscores the need for robust analytical frameworks to disentangle these effects.

Our analyses provide compelling evidence for a directional link between complex disease risk and longevity (**Supplementary Figs. 22-23** and **Supplementary Tables 8-9**). Importantly, these findings remained robust after adjusting for socioeconomic factors in the multivariable MR analysis. From these results, we derive several key findings with significant theoretical implications:

1. Disease burden and longevity: genetic liability to 22 complex diseases displayed significant causal effects on reduced longevity, remaining robust after adjusting for socioeconomic confounders and multiple testing.

2. Circulatory diseases drove the strongest effects, consistent with their established role in aging-related mortality (Tsao et al. 2023).

3. Late-onset diseases like hypertension, diverticular disease, lipoprotein metabolism disorders, coronary artery disease, myocardial infarction and heart failure exhibited the strongest causal effects. This predominance aligns with the Mutation Accumulation theory where deleterious alleles evading purifying selection disproportionately impact post-reproductive longevity (Medawar 1952).

In contrast, our analyses revealed less pronounced bidirectional causal effects between fertility and disease risk (**Supplementary Tables 10-13**). Although fertility was causally linked to conditions such as schizophrenia, breast cancer, and osteoarthritis, none of these associations survived multiple testing correction (**Supplementary Figs. 24-25**). The absence of robust findings may be attributable to weak instrument bias, which limits our power to detect true associations. Reverse MR analyses examining the impact of genetic liability to disease on fertility yielded only modest effect sizes, and the overall number of significant causal estimates was low, making it difficult to extract broad trends. Notably, for early-onset conditions such as childhood asthma and ADHD, the causal estimates were among the highest observed, linking genetic liability for disease to increased fertility. This observation aligns with the theory of Antagonistic Pleiotropy, where alleles conferring early-life fitness benefits persist despite their detrimental effects, suggesting a trade-off between disease risk and fitness advantages. Moreover, multivariable MR analyses that included education as a mediator showed that, although socioeconomic factors contribute to these relationships, they do not fully explain the observed causal effects. However, their role as potential mediators seemed to be more pronounced for fertility than for longevity.

General conclusions:

1. Our multivariable Mendelian randomization analyses incorporated educational attainment as a mediator to adjust for potential socioeconomic confounding. Although this mediator exerted measurable effects on both disease phenotypes and life-history traits, the residual causal associations persisted, indicating that educational attainment does not fully explain the observed relationships.

2. Sensitivity analyses—including MR-Egger regression and the weighted median method—revealed instances of horizontal pleiotropy, highlighting inconsistencies in causal estimates across different MR approaches (**Supplementary Tables 8-13**).

3. Furthermore, heterogeneity assessments using Cochran's Q statistic exposed potential weak instrument bias, particularly for certain diseases and more prominently for fertility.

By integrating socioeconomic factors through multivariable MR, we demonstrate that genetic predisposition to disease risk significantly influences both longevity and fertility—independent of confounding influences.

At the same time, our results stress the inherent challenges in disentangling pleiotropic effects and maintaining robust instrument strength in causal inference studies. Consequently, while our findings provide valuable insights, they should be interpreted with caution given the study's methodological limitations.

### References

Abdellaoui, Abdel, Hilary C. Martin, Martin Kolk, Adam Rutherford, Michael Muthukrishna, Felix C. Tropf, Melinda C. Mills, Brendan P. Zietsch, Karin J. H. Verweij, and Peter M. Visscher. 2025. “Socio-Economic Status Is a Social Construct with Heritable Components and Genetic Consequences.” *Nature Human Behaviour*, March, 1–13. https://doi.org/10.1038/s41562-025-02150-4.

Abdellatif, Mahmoud, Peter P. Rainer, Simon Sedej, and Guido Kroemer. 2023. “Hallmarks of Cardiovascular Ageing.” *Nature Reviews Cardiology* 20 (11): 754–77. https://doi.org/10.1038/s41569-023-00881-3.

Alex, Ashley, Eva Bhandary, and Kandace P. McGuire. 2020. “Anatomy and Physiology of the Breast during Pregnancy and Lactation.” In *Diseases of the Breast during Pregnancy and Lactation*, edited by Sadaf Alipour and Ramesh Omranipour, 3–7. Cham: Springer International Publishing. https://doi.org/10.1007/978-3-030-41596-9_1.

Allanore, Yannick, Mohamad Saad, Philippe Dieudé, Jérôme Avouac, Jorg H. W. Distler, Philippe Amouyel, Marco Matucci-Cerinic, et al. 2011. “Genome-Wide Scan Identifies TNIP1, PSORS1C1, and RHOB as Novel Risk Loci for Systemic Sclerosis.” *PLOS Genetics* 7 (7): e1002091. https://doi.org/10.1371/journal.pgen.1002091.

Alvergne, Alexandra, Markus Jokela, and Virpi Lummaa. 2010. “Personality and Reproductive Success in a High-Fertility Human Population.” *Proceedings of the National Academy of Sciences* 107 (26): 11745–50. https://doi.org/10.1073/pnas.1001752107.

Andreassen, Ole A., Wesley K. Thompson, Andrew J. Schork, Stephan Ripke, Morten Mattingsdal, John R. Kelsoe, Kenneth S. Kendler, et al. 2013. “Improved Detection of Common Variants Associated with Schizophrenia and Bipolar Disorder Using Pleiotropy-Informed Conditional False Discovery Rate.” *PLOS Genetics* 9 (4): e1003455. https://doi.org/10.1371/journal.pgen.1003455.

Azzolino, Domenico, Giulia Carla Immacolata Spolidoro, Edoardo Saporiti, Costanza Luchetti, Carlo Agostoni, and Matteo Cesari. 2021. “Musculoskeletal Changes Across the Lifespan: Nutrition and the Life-Course Approach to Prevention.” *Frontiers in Medicine* 8 (August):697954. https://doi.org/10.3389/fmed.2021.697954.

Bannister, Andrew J., and Tony Kouzarides. 2011. “Regulation of Chromatin by Histone Modifications.” *Cell Research* 21 (3): 381–95. https://doi.org/10.1038/cr.2011.22.

Barban, Nicola, Rick Jansen, Ronald de Vlaming, Ahmad Vaez, Jornt J. Mandemakers, Felix C. Tropf, Xia Shen, et al. 2016. “Genome-Wide Analysis Identifies 12 Loci Influencing Human Reproductive Behavior.” *Nature Genetics* 48 (12): 1462–72. https://doi.org/10.1038/ng.3698.

Basu, Sayon, Satya Pal Arya, Abul Usmani, Bhola Shankar Pradhan, Rajesh Kumar Sarkar, Nirmalya Ganguli, Mansi Shukla, et al. 2018. “Defective Wnt3 Expression by Testicular Sertoli Cells Compromise Male Fertility.” *Cell and Tissue Research* 371 (2): 351–63. https://doi.org/10.1007/s00441-017-2698-5.

Begovich, Ann B., Victoria E. H. Carlton, Lee A. Honigberg, Steven J. Schrodi, Anand P. Chokkalingam, Heather C. Alexander, Kristin G. Ardlie, et al. 2004. “A Missense Single-Nucleotide Polymorphism in a Gene Encoding a Protein Tyrosine Phosphatase (*PTPN22*) Is Associated with Rheumatoid Arthritis.” *The American Journal of Human Genetics* 75 (2): 330–37. https://doi.org/10.1086/422827.

Benonisdottir, Stefania, Vincent J. Straub, Augustine Kong, and Melinda C. Mills. 2024.

“Genetics of Female and Male Reproductive Traits and Their Relationship with Health, Longevity and Consequences for Offspring.” *Nature Aging* 4 (12): 1745–59. https://doi.org/10.1038/s43587-024-00733-w.

Birkenkamp-Demtroder, K., S. H. Olesen, F. B. Sørensen, S. Laurberg, P. Laiho, L. A. Aaltonen, and T. F. Ørntoft. 2005. “Differential Gene Expression in Colon Cancer of the Caecum versus the Sigmoid and Rectosigmoid.” *Gut* 54 (3): 374–84. https://doi.org/10.1136/gut.2003.036848.

Bottini, Nunzio, Lucia Musumeci, Andres Alonso, Souad Rahmouni, Konstantina Nika, Masoud Rostamkhani, James MacMurray, et al. 2004. “A Functional Variant of Lymphoid Tyrosine Phosphatase Is Associated with Type I Diabetes.” *Nature Genetics* 36 (4): 337–38. https://doi.org/10.1038/ng1323.

Bowden, Jack, George Davey Smith, and Stephen Burgess. 2015. “Mendelian Randomization with Invalid Instruments: Effect Estimation and Bias Detection through Egger Regression.” *International Journal of Epidemiology* 44 (2): 512–25. https://doi.org/10.1093/ije/dyv080.

Boyle, Evan A., Yang I. Li, and Jonathan K. Pritchard. 2017. “An Expanded View of Complex Traits: From Polygenic to Omnigenic.” *Cell* 169 (7): 1177–86. https://doi.org/10.1016/j.cell.2017.05.038.

Brewster, Karin L., and Ronald R. Rindfuss. 2000. “Fertility and Women’s Employment in Industrialized Nations.” *Annual Review of Sociology* 26:271–96.

Briley, Daniel A., Felix C. Tropf, and Melinda C. Mills. 2017. “What Explains the Heritability of Completed Fertility? Evidence from Two Large Twin Studies.” *Behavior Genetics* 47 (1): 36–51. https://doi.org/10.1007/s10519-016-9805-3.

Bulik-Sullivan, Brendan, Hilary K. Finucane, Verneri Anttila, Alexander Gusev, Felix R. Day, Po-Ru Loh, Laramie Duncan, et al. 2015. “An Atlas of Genetic Correlations across Human Diseases and Traits.” *Nature Genetics* 47 (11): 1236–41. https://doi.org/10.1038/ng.3406.

Caporaso, Neil, Fangyi Gu, Nilanjan Chatterjee, Jin Sheng-Chih, Kai Yu, Meredith Yeager, Constance Chen, et al. 2009. “Genome-Wide and Candidate Gene Association Study of Cigarette Smoking Behaviors.” *PLOS ONE* 4 (2): e4653. https://doi.org/10.1371/journal.pone.0004653.

Chang, Christopher C, Carson C Chow, Laurent CAM Tellier, Shashaank Vattikuti, Shaun M Purcell, and James J Lee. 2015. “Second-Generation PLINK: Rising to the Challenge of Larger and Richer Datasets.” *GigaScience* 4 (1): s13742-015-0047–0048. https://doi.org/10.1186/s13742-015-0047-8.

Chauhan, Ganesh, Corey R. Arnold, Audrey Y. Chu, Myriam Fornage, Azadeh Reyahi, Joshua C. Bis, Aki S. Havulinna, et al. 2016. “Identification of Additional Risk Loci for Stroke and Small Vessel Disease: A Meta-Analysis of Genome-Wide Association Studies.” *The Lancet Neurology* 15 (7): 695–707. https://doi.org/10.1016/S1474-4422(16)00102-2.

Choi, Shing Wan, Timothy Shin-Heng Mak, and Paul F. O’Reilly. 2020. “Tutorial: A Guide to Performing Polygenic Risk Score Analyses.” *Nature Protocols* 15 (9): 2759–72. https://doi.org/10.1038/s41596-020-0353-1.

Christopher J L Murray. 2020. “Global Burden of 87 Risk Factors in 204 Countries and Territories, 1990-2019: A Systematic Analysis for the Global Burden of Disease Study 2019.” *Lancet (London, England)* 396 (10258): 1223–49. https://doi.org/10.1016/s0140-6736(20)30752-2.

Collister, Jennifer A., Xiaonan Liu, and Lei Clifton. 2022. “Calculating Polygenic Risk Scores (PRS) in UK Biobank: A Practical Guide for Epidemiologists.” *Frontiers in Genetics* 13 (February). https://doi.org/10.3389/fgene.2022.818574.

Das, Sayantan, Lukas Forer, Sebastian Schönherr, Carlo Sidore, Adam E. Locke, Alan Kwong, Scott I. Vrieze, et al. 2016. “Next-Generation Genotype Imputation Service and Methods.” *Nature Genetics* 48 (10): 1284–87. https://doi.org/10.1038/ng.3656.

Davey Smith, George, and Gibran Hemani. 2014. “Mendelian Randomization: Genetic Anchors for Causal Inference in Epidemiological Studies.” *Human Molecular Genetics* 23 (R1): R89–98. https://doi.org/10.1093/hmg/ddu328.

Eilbeck, Karen, Suzanna E. Lewis, Christopher J. Mungall, Mark Yandell, Lincoln Stein, Richard Durbin, and Michael Ashburner. 2005. “The Sequence Ontology: A Tool for the Unification of Genome Annotations.” *Genome Biology* 6 (5): R44. https://doi.org/10.1186/gb-2005-6-5-r44.

Field, Yair, Evan A Boyle, Natalie Telis, Ziyue Gao, Kyle J. Gaulton, David Golan, Loic Yengo, et al. 2016. “Detection of Human Adaptation during the Past 2000 Years.” *Science* 354 (6313): 760–64. https://doi.org/10.1126/science.aag0776.

Frejka, Tomas. 2016. “The Demographic Transition Revisited: A Cohort Perspective.” WP-2016-012. 0 ed. Rostock: Max Planck Institute for Demographic Research. https://doi.org/10.4054/MPIDR-WP-2016-012.

Fritz, Andrew J., Prachi N. Ghule, Joseph R. Boyd, Coralee E. Tye, Natalie A. Page, Deli Hong, David J. Shirley, et al. 2018. “Intranuclear and Higher-Order Chromatin Organization of the Major Histone Gene Cluster in Breast Cancer.” *Journal of Cellular Physiology* 233 (2): 1278–90. https://doi.org/10.1002/jcp.25996.

Galor, Oded. 2012. “The Demographic Transition: Causes and Consequences.” *Cliometrica* 6 (1): 1–28. https://doi.org/10.1007/s11698-011-0062-7.

Garg, Deepika, and Sarah L. Berga. 2020. “Chapter 1 - Neuroendocrine Mechanisms of Reproduction.” In *Handbook of Clinical Neurology*, edited by Eric A. P. Steegers, Marilyn J. Cipolla, and Eliza C. Miller, 171:3–23. Neurology and Pregnancy. Elsevier. https://doi.org/10.1016/B978-0-444-64239-4.00001-1.

Ge, Tian, Chia-Yen Chen, Yang Ni, Yen-Chen Anne Feng, and Jordan W. Smoller. 2019. “Polygenic Prediction via Bayesian Regression and Continuous Shrinkage Priors.” *Nature Communications* 10 (1): 1776. https://doi.org/10.1038/s41467-019-09718-5.

Guo, Yanjun, Wonil Chung, Zhilei Shan, Zhaozhong Zhu, Karen H. Costenbader, and Liming Liang. 2023. “Genome-Wide Assessment of Shared Genetic Architecture Between Rheumatoid Arthritis and Cardiovascular Diseases.” *Journal of the American Heart Association* 12 (22): e030211. https://doi.org/10.1161/JAHA.123.030211.

Guzzo, Karen Benjamin. 2014. “New Partners, More Kids: Multiple-Partner Fertility in the United States.” *The ANNALS of the American Academy of Political and Social Science* 654 (1): 66–86. https://doi.org/10.1177/0002716214525571.

Hall, Jeff M., Ming K. Lee, Beth Newman, Jan E. Morrow, Lee A. Anderson, Bing Huey, and Mary-Claire King. 1990. “Linkage of Early-Onset Familial Breast Cancer to Chromosome 17q21.” *Science* 250 (4988): 1684–89. https://doi.org/DOI: 10.1126/science.2270482.

Harrison, Peter W, M Ridwan Amode, Olanrewaju Austine-Orimoloye, Andrey G Azov, Matthieu Barba, If Barnes, Arne Becker, et al. 2024. “Ensembl 2024.” *Nucleic Acids Research* 52 (D1): D891–99. https://doi.org/10.1093/nar/gkad1049.

Havlicek, Jan, and S. Craig Roberts. 2009. “MHC-Correlated Mate Choice in Humans: A Review.” *Psychoneuroendocrinology* 34 (4): 497–512. https://doi.org/10.1016/j.psyneuen.2008.10.007.

Hayward, Laura Katharine, and Guy Sella. 2022. “Polygenic Adaptation after a Sudden Change in Environment.” Edited by Graham Coop, George H Perry, Peter L Ralph, and Guillaume Martin. *eLife* 11 (September):e66697.

https://doi.org/10.7554/eLife.66697.

Hemani, Gibran, Jack Bowden, and George Davey Smith. 2018. “Evaluating the Potential Role of Pleiotropy in Mendelian Randomization Studies.” *Human Molecular Genetics* 27 (R2): R195–208. https://doi.org/10.1093/hmg/ddy163.

Hill, W. David, Saskia P. Hagenaars, Riccardo E. Marioni, Sarah E. Harris, David C. M. Liewald, Gail Davies, Aysu Okbay, Andrew M. McIntosh, Catharine R. Gale, and Ian J. Deary. 2016. “Molecular Genetic Contributions to Social Deprivation and Household Income in UK Biobank.” *Current Biology: CB* 26 (22): 3083–89. https://doi.org/10.1016/j.cub.2016.09.035.

Hirsch, J.A., G. Nicola, G. McGinty, R.W. Liu, R.M. Barr, M.D. Chittle, and L. Manchikanti. 2016. “ICD-10: History and Context.” *AJNR: American Journal of Neuroradiology* 37 (4): 596–99. https://doi.org/10.3174/ajnr.A4696.

Horton, Roger, Richard Gibson, Penny Coggill, Marcos Miretti, Richard J. Allcock, Jeff Almeida, Simon Forbes, et al. 2008. “Variation Analysis and Gene Annotation of Eight MHC Haplotypes: The MHC Haplotype Project.” *Immunogenetics* 60 (1): 1–18. https://doi.org/10.1007/s00251-007-0262-2.

Jeselsohn, Rinath, Gilles Buchwalter, Carmine De Angelis, Myles Brown, and Rachel Schiff. 2015. “ESR1 Mutations—a Mechanism for Acquired Endocrine Resistance in Breast Cancer.” *Nature Reviews Clinical Oncology* 12 (10): 573–83. https://doi.org/10.1038/nrclinonc.2015.117.

Jia, Gengjie, Yu Li, Hanxin Zhang, Ishanu Chattopadhyay, Anders Boeck Jensen, David R. Blair, Lea Davis, et al. 2019. “Estimating Heritability and Genetic Correlations from Large Health Datasets in the Absence of Genetic Data.” *Nature Communications* 10 (1): 5508. https://doi.org/10.1038/s41467-019-13455-0.

Jiang, Xia, Paul F. O’Reilly, Hugues Aschard, Yi-Hsiang Hsu, J. Brent Richards, Josée Dupuis, Erik Ingelsson, et al. 2018. “Genome-Wide Association Study in 79,366 European-Ancestry Individuals Informs the Genetic Architecture of 25-Hydroxyvitamin D Levels.” *Nature Communications* 9 (1): 260. https://doi.org/10.1038/s41467-017-02662-2.

Jokela, Markus, Alexandra Alvergne, Thomas V. Pollet, and Virpi Lummaa. 2011. “Reproductive Behavior and Personality Traits of the Five Factor Model.” *European Journal of Personality* 25 (6): 487–500. https://doi.org/10.1002/per.822.

Junnila, Riia K., Edward O. List, Darlene E. Berryman, John W. Murrey, and John J. Kopchick. 2013. “The GH/IGF-1 Axis in Ageing and Longevity.” *Nature Reviews Endocrinology* 9 (6): 366–76. https://doi.org/10.1038/nrendo.2013.67.

Keebler, Mary E., Rahul C. Deo, Aarti Surti, David Konieczkowski, Candace Guiducci, Noel Burtt, Sarah G. Buxbaum, et al. 2010. “Fine-Mapping in African Americans of 8 Recently Discovered Genetic Loci for Plasma Lipids.” *Circulation: Cardiovascular Genetics* 3 (4): 358–64. https://doi.org/10.1161/CIRCGENETICS.109.914267.

Kirk, Dudley. 1996. “Demographic Transition Theory.” *Population Studies* 50 (3): 361–87. https://doi.org/10.1080/0032472031000149536.

Krishnan, Sudha, Karlie A. Intlekofer, Leah K. Aggison, and Sandra L. Petersen. 2009. “Central Role of TRAF-Interacting Protein in a New Model of Brain Sexual Differentiation.” *Proceedings of the National Academy of Sciences* 106 (39): 16692–97. https://doi.org/10.1073/pnas.0906293106.

Kurki, Mitja I., Juha Karjalainen, Priit Palta, Timo P. Sipilä, Kati Kristiansson, Kati M. Donner, Mary P. Reeve, et al. 2023. “FinnGen Provides Genetic Insights from a Well-Phenotyped Isolated Population.” *Nature* 613 (7944): 508–18. https://doi.org/10.1038/s41586-022-05473-8.

Kurscheid, Sebastian, Pierre Bady, Davide Sciuscio, Ivana Samarzija, Tal Shay, Irene

Vassallo, Wim V. Criekinge, et al. 2015. “Chromosome 7 Gain and DNA Hypermethylation at the HOXA10 Locus Are Associated with Expression of a Stem Cell Related HOX-Signature in Glioblastoma.” *Genome Biology* 16 (1): 16. https://doi.org/10.1186/s13059-015-0583-7.

Lamp, Merit, Maire Peters, Eva Reinmaa, Kadri Haller-Kikkatalo, Tanel Kaart, Ülle Kadastik, Helle Karro, Andres Metspalu, and Andres Salumets. 2011. “Polymorphisms in ESR1, ESR2 and HSD17B1 Genes Are Associated with Fertility Status in Endometriosis.” *Gynecological Endocrinology* 27 (6): 425–33. https://doi.org/10.3109/09513590.2010.495434.

Lee, James J., Robbee Wedow, Aysu Okbay, Edward Kong, Omeed Maghzian, Meghan Zacher, Tuan Anh Nguyen-Viet, et al. 2018. “Gene Discovery and Polygenic Prediction from a Genome-Wide Association Study of Educational Attainment in 1.1 Million Individuals.” *Nature Genetics* 50 (8): 1112–21. https://doi.org/10.1038/s41588-018-0147-3.

Lee, Sang Hong, Michael E Goddard, Naomi R Wray, and Peter M Visscher. 2012. “A Better Coefficient of Determination for Genetic Profile Analysis.” *Genetic Epidemiology* 36 (3): 214–24. https://doi.org/10.1002/gepi.21614.

Lerchbaum, Elisabeth, and Barbara Obermayer-Pietsch. 2012. “MECHANISMS IN ENDOCRINOLOGY: Vitamin D and Fertility: A Systematic Review.” *European Journal of Endocrinology* 166 (5): 765–78. https://doi.org/10.1530/EJE-11-0984.

Li, Bing, Michael Carey, and Jerry L. Workman. 2007. “The Role of Chromatin during Transcription.” *Cell* 128 (4): 707–19. https://doi.org/10.1016/j.cell.2007.01.015.

Long, Erping, and Jianzhi Zhang. 2021. “Natural Selection Contributes to the Myopia Epidemic.” *National Science Review* 8 (6): nwaa175. https://doi.org/10.1093/nsr/nwaa175.

López González, Irene, Paula Garcia-Esparcia, Franc Llorens, and Isidre Ferrer. 2016. “Genetic and Transcriptomic Profiles of Inflammation in Neurodegenerative Diseases: Alzheimer, Parkinson, Creutzfeldt-Jakob and Tauopathies.” *International Journal of Molecular Sciences* 17 (2): 206. https://doi.org/10.3390/ijms17020206.

López-Otín, Carlos, Lorenzo Galluzzi, José M. P. Freije, Frank Madeo, and Guido Kroemer. 2016. “Metabolic Control of Longevity.” *Cell* 166 (4): 802–21. https://doi.org/10.1016/j.cell.2016.07.031.

Manolio, Teri A., Francis S. Collins, Nancy J. Cox, David B. Goldstein, Lucia A. Hindorff, David J. Hunter, Mark I. McCarthy, et al. 2009. “Finding the Missing Heritability of Complex Diseases.” *Nature* 461 (7265): 747–53. https://doi.org/10.1038/nature08494.

Marees, Andries T., Hilde de Kluiver, Sven Stringer, Florence Vorspan, Emmanuel Curis, Cynthia Marie-Claire, and Eske M. Derks. 2018. “A Tutorial on Conducting Genome-Wide Association Studies: Quality Control and Statistical Analysis.” *International Journal of Methods in Psychiatric Research* 27 (2): e1608. https://doi.org/10.1002/mpr.1608.

Martín, Teresa Castro. 1995. “Women’s Education and Fertility: Results from 26 Demographic and Health Surveys.” *Studies in Family Planning* 26 (4): 187–202. https://doi.org/10.2307/2137845.

Mathieson, Iain, Felix R. Day, Nicola Barban, Felix C. Tropf, David M. Brazel, Diana van Heemst, Ahmad Vaez, et al. 2023. “Genome-Wide Analysis Identifies Genetic Effects on Reproductive Success and Ongoing Natural Selection at the FADS Locus.” *Nature Human Behaviour* 7 (5): 790–801. https://doi.org/10.1038/s41562-023-01528-6.

Matsuyama, Satoko, Sarah Whiteside, and Shu-Yun Li. 2024. “Implantation and Decidualization in PCOS: Unraveling the Complexities of Pregnancy.” *International Journal of Molecular Sciences* 25 (2): 1203. https://doi.org/10.3390/ijms25021203.

Mattos, Clarissa Santiago de, Camila Martins Trevisan, Carla Peluso, Fernando Adami, Emerson Barchi Cordts, Denise Maria Christofolini, Caio Parente Barbosa, and Bianca Bianco. 2014. "ESR1 and ESR2 Gene Polymorphisms Are Associated with Human Reproduction Outcomes in Brazilian Women." *Journal of Ovarian Research* 7 (1): 114. https://doi.org/10.1186/s13048-014-0114-2.

Maynard Smith, John. 1998. *Evolutionary Genetics*. 2nd ed. Oxford University Press. https://cir.nii.ac.jp/crid/1130282271506505088.

McLaren, William, Laurent Gil, Sarah E. Hunt, Harpreet Singh Riat, Graham R. S. Ritchie, Anja Thormann, Paul Flicek, and Fiona Cunningham. 2016. "The Ensembl Variant Effect Predictor." *Genome Biology* 17 (1): 122. https://doi.org/10.1186/s13059-016-0974-4.

Medawar, P.B. 1952. *An Unsolved Problem of Biology: An Inaugural Lecture Delivered at University College, London, 6 December, 1951*. London: H.K. Lewis and Company.

Mills, Melinda C., and Felix C. Tropf. 2015. "The Biodemography of Fertility: A Review and Future Research Frontiers." *Kolner Zeitschrift Fur Soziologie Und Sozialpsychologie* 67 (Suppl 1): 397–424. https://doi.org/10.1007/s11577-015-0319-4.

Mills, Melinda, Ronald R. Rindfuss, Peter McDonald, Egbert te Velde, and on behalf of the ESHRE Reproduction and Society Task Force. 2011. "Why Do People Postpone Parenthood? Reasons and Social Policy Incentives." *Human Reproduction Update* 17 (6): 848–60. https://doi.org/10.1093/humupd/dmr026.

Mubarik, Sumaira, Shafaq Naeem, Hui Shen, Rabia Mubarak, Lisha Luo, Syeda Rija Hussain, Eelko Hak, Chuanhua Yu, and Xiaoxue Liu. 2024. "Population-Level Distribution, Risk Factors, and Burden of Mortality and Disability-Adjusted Life Years Attributable to Major Noncommunicable Diseases in Western Europe (1990-2021): Ecological Analysis." *JMIR Public Health and Surveillance* 10 (October):e57840. https://doi.org/10.2196/57840.

Murga-Moreno, Jesús, Marta Coronado-Zamora, Alejandra Bodelón, Antonio Barbadilla, and Sònia Casillas. 2019. "PopHumanScan: The Online Catalog of Human Genome Adaptation." *Nucleic Acids Research* 47 (D1): D1080–89. https://doi.org/10.1093/nar/gky959.

Murthy, Megha N., Cornelis Blauwendraat, Sebastian Guelfi, John Hardy, Patrick A. Lewis, Daniah Trabzuni, UKBEC, and IPDGC. 2017. "Increased Brain Expression of GPNMB Is Associated with Genome Wide Significant Risk for Parkinson's Disease on Chromosome 7p15.3." *Neurogenetics* 18 (3): 121–33. https://doi.org/10.1007/s10048-017-0514-8.

Narod, Steven A., and William D. Foulkes. 2004. "BRCA1 and BRCA2: 1994 and Beyond." *Nature Reviews Cancer* 4 (9): 665–76. https://doi.org/10.1038/nrc1431.

Ni, Guiyan, Azmeraw T. Amare, Xuan Zhou, Natalie Mills, Jacob Gratten, and S. Hong Lee. 2019. "The Genetic Relationship between Female Reproductive Traits and Six Psychiatric Disorders." *Scientific Reports* 9 (1): 12041. https://doi.org/10.1038/s41598-019-48403-x.

Nielsen, Rasmus. 2005. "Molecular Signatures of Natural Selection." *Annual Review of Genetics* 39 (Volume 39, 2005): 197–218. https://doi.org/10.1146/annurev.genet.39.073003.112420.

Noble, C. L., A. R. Abbas, J. Cornelius, C. W. Lees, G.-T. Ho, K. Toy, Z. Modrusan, et al. 2008. "Regional Variation in Gene Expression in the Healthy Colon Is Dysregulated in Ulcerative Colitis." *Gut* 57 (10): 1398–1405. https://doi.org/10.1136/gut.2008.148395.

Ober, C. 1999. "Studies of HLA, Fertility and Mate Choice in a Human Isolate." *Human Reproduction Update* 5 (2): 103–7. https://doi.org/10.1093/humupd/5.2.103.

Oishi, Kohei, Seiya Yamayoshi, Hiroko Kozuka-Hata, Masaaki Oyama, and Yoshihiro Kawaoka. 2018. “N-Terminal Acetylation by NatB Is Required for the Shutoff Activity of Influenza A Virus PA-X.” *Cell Reports* 24 (4): 851–60. https://doi.org/10.1016/j.celrep.2018.06.078.

Parhar, Ishwar S., Satoshi Ogawa, and Takayoshi Ubuka. 2016. “Reproductive Neuroendocrine Pathways of Social Behavior.” *Frontiers in Endocrinology* 7 (March). https://doi.org/10.3389/fendo.2016.00028.

Purcell, Shaun, Benjamin Neale, Kathe Todd-Brown, Lori Thomas, Manuel A. R. Ferreira, David Bender, Julian Maller, et al. 2007. “PLINK: A Tool Set for Whole-Genome Association and Population-Based Linkage Analyses.” *The American Journal of Human Genetics* 81 (3): 559–75. https://doi.org/10.1086/519795.

Rasmussen, Maria, Else Marie Vestergaard, Jesper Graakjaer, Yanko Petkov, Iben Bache, Christina Fagerberg, Maria Kibæk, et al. 2016. “17q12 Deletion and Duplication Syndrome in Denmark—A Clinical Cohort of 38 Patients and Review of the Literature.” *American Journal of Medical Genetics Part A* 170 (11): 2934–42. https://doi.org/10.1002/ajmg.a.37848.

Sanchez-Roige, Sandra, Mariela V. Jennings, Hayley H. A. Thorpe, Jazlene E. Mallari, Lieke C. van der Werf, Sevim B. Bianchi, Yuye Huang, et al. 2023. “CADM2 Is Implicated in Impulsive Personality and Numerous Other Traits by Genome- and Phenome-Wide Association Studies in Humans and Mice.” *Translational Psychiatry* 13 (1): 1–11. https://doi.org/10.1038/s41398-023-02453-y.

Shiina, Takashi, Kazuyoshi Hosomichi, Hidetoshi Inoko, and Jerzy K. Kulski. 2009. “The HLA Genomic Loci Map: Expression, Interaction, Diversity and Disease.” *Journal of Human Genetics* 54 (1): 15–39. https://doi.org/10.1038/jhg.2008.5.

Simunovic, Filip, Ming Yi, Yulei Wang, Laurel Macey, Lauren T. Brown, Anna M. Krichevsky, Susan L. Andersen, Robert M. Stephens, Francine M. Benes, and Kai C. Sonntag. 2009. “Gene Expression Profiling of Substantia Nigra Dopamine Neurons: Further Insights into Parkinson’s Disease Pathology.” *Brain* 132 (7): 1795–1809. https://doi.org/10.1093/brain/awn323.

Singh, Vertika, Renu Bala, Arijit Chakraborty, Singh Rajender, Sameer Trivedi, and Kiran Singh. 2019. “Duplications in 19p13.3 Are Associated with Male Infertility.” *Journal of Assisted Reproduction and Genetics* 36 (10): 2171–79. https://doi.org/10.1007/s10815-019-01547-1.

Skirbekk, Vegard. 2008. “Fertility Trends by Social Status.” *Demographic Research* 18:145–80.

Smeland, Olav B., Oleksandr Frei, Alexey Shadrin, Kevin O’Connell, Chun-Chieh Fan, Shahram Bahrami, Dominic Holland, et al. 2020. “Discovery of Shared Genomic Loci Using the Conditional False Discovery Rate Approach.” *Human Genetics* 139 (1): 85–94. https://doi.org/10.1007/s00439-019-02060-2.

Sobotka, Tomáš. 2017. “Post-Transitional Fertility: The Role Of Childbearing Postponement In Fuelling The Shift To Low And Unstable Fertility Levels.” *Journal of Biosocial Science* 49 (S1): S20–45. https://doi.org/10.1017/S0021932017000323.

Starheim, Kristian K., Thomas Arnesen, Darina Gromyko, Anita Ryningen, Jan Erik Varhaug, and Johan R. Lillehaug. 2008. “Identification of the Human N(Alpha)-Acetyltransferase Complex B (hNatB): A Complex Important for Cell-Cycle Progression.” *The Biochemical Journal* 415 (2): 325–31. https://doi.org/10.1042/BJ20080658.

Stein, Michelle M., Emma E. Thompson, Nathan Schoettler, Britney A. Helling, Kevin M. Magnaye, Catherine Stanhope, Catherine Igartua, et al. 2018. “A Decade of Research on the 17q12-21 Asthma Locus: Piecing Together the Puzzle.” *Journal of Allergy and*

*Clinical Immunology* 142 (3): 749-764.e3. https://doi.org/10.1016/j.jaci.2017.12.974.

Sudlow, Cathie, John Gallacher, Naomi Allen, Valerie Beral, Paul Burton, John Danesh, Paul Downey, et al. 2015. “UK Biobank: An Open Access Resource for Identifying the Causes of a Wide Range of Complex Diseases of Middle and Old Age.” *PLOS Medicine* 12 (3): e1001779. https://doi.org/10.1371/journal.pmed.1001779.

Tanturri, Maria Letizia, and Letizia Mencarini. 2008. “Childless or Childfree? Paths to Voluntary Childlessness in Italy.” *Population and Development Review* 34 (1): 51–77. https://doi.org/10.1111/j.1728-4457.2008.00205.x.

Teixeira, Vitor, and Vítor Costa. 2016. “Unraveling the Role of the Target of Rapamycin Signaling in Sphingolipid Metabolism.” *Progress in Lipid Research* 61 (January):109–33. https://doi.org/10.1016/j.plipres.2015.11.001.

The COVID-19 Host Genetics Initiative. 2020. “The COVID-19 Host Genetics Initiative, a Global Initiative to Elucidate the Role of Host Genetic Factors in Susceptibility and Severity of the SARS-CoV-2 Virus Pandemic.” *European Journal of Human Genetics* 28 (6): 715–18. https://doi.org/10.1038/s41431-020-0636-6.

THE GTEX CONSORTIUM. 2020. “The GTEx Consortium Atlas of Genetic Regulatory Effects across Human Tissues.” *Science* 369 (6509): 1318–30. https://doi.org/10.1126/science.aaz1776.

Tropf, Felix C., Gert Stulp, Nicola Barban, Peter M. Visscher, Jian Yang, Harold Snieder, and Melinda C. Mills. 2015. “Human Fertility, Molecular Genetics, and Natural Selection in Modern Societies.” *PLOS ONE* 10 (6): e0126821. https://doi.org/10.1371/journal.pone.0126821.

Tsao, Connie W., Aaron W. Aday, Zaid I. Almarzooq, Cheryl A.M. Anderson, Pankaj Arora, Christy L. Avery, Carissa M. Baker-Smith, et al. 2023. “Heart Disease and Stroke Statistics—2023 Update: A Report From the American Heart Association.” *Circulation* 147 (8): e93–621. https://doi.org/10.1161/CIR.0000000000001123.

Uffelmann, Emil, Qin Qin Huang, Nchangwi Syntia Munung, Jantina de Vries, Yukinori Okada, Alicia R. Martin, Hilary C. Martin, Tuuli Lappalainen, and Danielle Posthuma. 2021. “Genome-Wide Association Studies.” *Nature Reviews Methods Primers* 1 (1): 1–21. https://doi.org/10.1038/s43586-021-00056-9.

Verbanck, Marie, Chia-Yen Chen, Benjamin Neale, and Ron Do. 2018. “Detection of Widespread Horizontal Pleiotropy in Causal Relationships Inferred from Mendelian Randomization between Complex Traits and Diseases.” *Nature Genetics* 50 (5): 693–98. https://doi.org/10.1038/s41588-018-0099-7.

Viippola, Essi, Sara Kuitunen, Rodosthenis S Rodosthenous, Andrius Vabalas, Tuomo Hartonen, Pekka Vartiainen, Joanne Demmler, et al. 2023. “Data Resource Profile: Nationwide Registry Data for High-Throughput Epidemiology and Machine Learning (FinRegistry).” *International Journal of Epidemiology* 52 (4): e195–200. https://doi.org/10.1093/ije/dyad091.

Voight, Benjamin F., Sridhar Kudaravalli, Xiaoquan Wen, and Jonathan K. Pritchard. 2006. “A Map of Recent Positive Selection in the Human Genome.” *PLOS Biology* 4 (3): e72. https://doi.org/10.1371/journal.pbio.0040072.

Wang, Quan, Yu-Gang Wen, Da-Peng Li, Jun Xia, Chong-Zhi Zhou, Dong-Wang Yan, Hua-Mei Tang, and Zhi-Hai Peng. 2012. “Upregulated INHBA Expression Is Associated with Poor Survival in Gastric Cancer.” *Medical Oncology* 29 (1): 77–83. https://doi.org/10.1007/s12032-010-9766-y.

Wang, Zheng, Qiqi Zhang, Chen Zhang, Jun Yan, Tingting Yang, and Aifang Jiang. 2024. “CADM2 Participates in Endometriosis Development by Influencing the Epithelial-Mesenchymal Transition.” *Reproductive Sciences* 31 (10): 3049–57. https://doi.org/10.1007/s43032-024-01592-x.

Watanabe, Kyoko, Sven Stringer, Oleksandr Frei, Maša Umićević Mirkov, Christiaan de Leeuw, Tinca J. C. Polderman, Sophie van der Sluis, Ole A. Andreassen, Benjamin M. Neale, and Danielle Posthuma. 2019. "A Global Overview of Pleiotropy and Genetic Architecture in Complex Traits." *Nature Genetics* 51 (9): 1339–48. https://doi.org/10.1038/s41588-019-0481-0.

Watanabe, Kyoko, Erdogan Taskesen, Arjen van Bochoven, and Danielle Posthuma. 2017. "Functional Mapping and Annotation of Genetic Associations with FUMA." *Nature Communications* 8 (1): 1826. https://doi.org/10.1038/s41467-017-01261-5.

Western, Daniel, Jigyasha Timsina, Lihua Wang, Ciyang Wang, Chengran Yang, Bridget Phillips, Yueyao Wang, et al. 2024. "Proteogenomic Analysis of Human Cerebrospinal Fluid Identifies Neurologically Relevant Regulation and Implicates Causal Proteins for Alzheimer's Disease." *Nature Genetics* 56 (12): 2672–84. https://doi.org/10.1038/s41588-024-01972-8.

Willer, Cristen J., Yun Li, and Gonçalo R. Abecasis. 2010. "METAL: Fast and Efficient Meta-Analysis of Genomewide Association Scans." *Bioinformatics* 26 (17): 2190–91. https://doi.org/10.1093/bioinformatics/btq340.

Williams, GC. 1957. "Pleiotropy, Natural Selection, and the Evolution of Senescence." *Evolution (NY)* 11:398–411. https://doi.org/10.2307/2406060.

Williams, M. R., K. Galvin, B. O'Sullivan, C. D. MacDonald, E. W. K. Ching, F. Turkheimer, O. D. Howes, R. K. B. Pearce, S. R. Hirsch, and M. Maier. 2014. "Neuropathological Changes in the Substantia Nigra in Schizophrenia but Not Depression." *European Archives of Psychiatry and Clinical Neuroscience* 264 (4): 285–96. https://doi.org/10.1007/s00406-013-0479-z.

Wiśniewski, Andrzej, Łukasz Matusiak, Aneta Szczerkowska-Dobosz, Izabela Nowak, and Piotr Kuśnierczyk. 2018. "HLA-C*06:02-Independent, Gender-Related Association of PSORS1C3 and PSORS1C1/CDSN Single-Nucleotide Polymorphisms with Risk and Severity of Psoriasis." *Molecular Genetics and Genomics* 293 (4): 957–66. https://doi.org/10.1007/s00438-018-1435-4.

World Health Assembly, 43. 1990. "Report of the International Conference for the Tenth Revision of the International Classification of Diseases." https://iris.who.int/handle/10665/173188.

Wu, R. Alex, Daniel R. Semlow, Ashley N. Kamimae-Lanning, Olga V. Kochenova, Gheorghe Chistol, Michael R. Hodskinson, Ravindra Amunugama, et al. 2019. "TRAIP Is a Master Regulator of DNA Interstrand Crosslink Repair." *Nature* 567 (7747): 267–72. https://doi.org/10.1038/s41586-019-1002-0.

Xu, Jingkai, Zhi Li, Xianbo Zuo, Guozheng Li, Xuejun Zhang, Bo Zhang, and Yong Cui. 2022. "Knockdown of NAA25 Suppresses Breast Cancer Progression by Regulating Apoptosis and Cell Cycle." *Frontiers in Oncology* 11 (January). https://doi.org/10.3389/fonc.2021.755267.

Young, Alexander I. 2019. "Solving the Missing Heritability Problem." *PLOS Genetics* 15 (6): e1008222. https://doi.org/10.1371/journal.pgen.1008222.

Zeberg, Hugo, and Svante Pääbo. 2020. "The Major Genetic Risk Factor for Severe COVID-19 Is Inherited from Neanderthals." *Nature* 587 (7835): 610–12. https://doi.org/10.1038/s41586-020-2818-3.

Zietsch, Brendan P., Ralf Kuja-Halkola, Hasse Walum, and Karin J. H. Verweij. 2014. "Perfect Genetic Correlation between Number of Offspring and Grandoffspring in an Industrialized Human Population." *Proceedings of the National Academy of Sciences* 111 (3): 1032–36. https://doi.org/10.1073/pnas.1310058111.
